# Gene and metabolite expression dependence on body mass index in human myocardium

**DOI:** 10.1101/2021.06.29.21259697

**Authors:** Adewale S Adebayo, Marius Roman, Syabira Yusoff, Melanie Gulston, Lathishia Joel-David, Bony Anthony, Florence Y Lai, Antonio Murgia, Bryony Eagle-Hemming, Sophia Sheikh, Tracy Kumar, Hardeep Aujla, Will Dott, Julian L Griffin, Gavin J Murphy, Marcin J Woźniak

**Affiliations:** Department of Cardiovascular Sciences and NIHR Cardiovascular Biomedical Research Unit, University of Leicester, Glenfield Hospital, Leicester, LE3 9QP, UK; Department of Biochemistry and Cambridge Systems Biology Centre, The Sanger Building, 80 Tennis Court Road, Cambridge, CB2 1GA; Cardiovascular Sciences, King’s College London, London, UKGSE102088; Biomolecular Medicine, Department of Metabolism, Digestion and Reproduction, The Sir Alexander Fleming Building, Imperial College London, Exhibition Road, South Kensington, London, SW7 2AZ, UK

**Keywords:** Obesity, Cardiac Surgical Procedures, Gene Expression Profiling, Metabolome

## Abstract

We hypothesized that body mass index (BMI) dependent changes in myocardial gene expression and energy-related metabolites underlie the biphasic association between BMI and mortality (the obesity paradox) in cardiac surgery. We performed transcriptome profiling and measured a panel of 144 metabolites in 53 and 55, respectively, myocardial biopsies from a cohort of sixty-seven adult patients undergoing coronary artery bypass grafting (registration: NCT02908009). The initial analysis identified 239 transcripts with biphasic BMI dependence. 120 displayed u-shape and 119 n-shape expression patterns. The identified local minima or maxima peaked at BMI 28-29. Based on these results and to best fit the WHO classification, we grouped the patients into three groups: BMI<25, 25≤BMI≤32, and BMI>32. The group analysis indicated that protein translation-related pathways were downregulated in 25≤BMI≥32 compared with BMI<25 patients. Muscle contraction transcripts were upregulated in 25≤BMI≥32 patients, and cholesterol synthesis and innate immunity transcripts were upregulated in the BMI>32 group. Transcripts involved in translation, muscle contraction and lipid metabolism also formed distinct correlation networks with biphasic dependence on BMI. Metabolite analysis identified acylcarnitines and ribose-5-phosphate increasing in the BMI>32 group and α-ketoglutarate increasing in the BMI<25 group. Molecular differences in the myocardium mirror the biphasic relationship between BMI and mortality.

## Background

Elevated Body Mass Index (BMI) is an important risk factor for heart failure and cardiovascular death. ^1^ However, recent studies have reported a biphasic u-shaped relationship between increasing BMI and mortality in clinical settings characterized by acute metabolic stress such as cardiac surgery, ^2,3^ acute coronary syndromes, ^4^ heart failure, ^5^ and in patients requiring dialysis.^6^. Here people with BMI between 25 and 35 have paradoxically better survival than those with low or normal BMI, or very high BMI (>35). These observations may be attributable to reverse epidemiology where people who are underweight or who have severe obesity have worse outcomes attributable to frailty or sarcopenia, or to unmeasured confounding such as fitness, or the presence or absence of metabolic syndrome. Both severe obesity and frailty lead to mitochondrial dysfunction ^7^ and dysregulated bioenergetics ^8^. We hypothesized that these may contribute to the biphasic association between BMI and adverse events following cardiac surgery. We tested this in human myocardial biopsies subjected to untargeted next generation sequencing and targeted metabolomics acquired as part of an ongoing observational study. The aim was to identify genes and energy metabolites whose expression in the biopsies show a biphasic response to increasing BMI and to evaluate how these differences could translate into differences in clinical outcomes.

## Methods

### Study design

Ob-Card – a Case-Control Study to Identify the Role of Epigenetic Regulation of Genes Responsible for Energy Metabolism and Mitochondrial Function in the Obesity Paradox in Cardiac Surgery was a prospective observational study approved by The East Midlands – Nottingham 1 Research Ethics Committee. The study protocol was registered at https://clinicaltrials.gov/ct2/show/NCT02908009. The study is reported as per the STrengthening the Reporting of Observational Studies in Epidemiology (STROBE) statement.

### Study cohort

Adult cardiac surgery patients (>16 years) undergoing coronary artery bypass grafting with or without valve surgery. Patients with pre-existing paroxysmal, persistent or chronic atrial fibrillation, pre-existing inflammatory state (sepsis undergoing treatment, acute kidney injury within five days, chronic inflammatory disease, congestive heart failure), ejection fraction <30 %, pregnancy and in a critical preoperative state (Kidney Disease: Improving Global Outcomes (KDIGO) Stage 3 AKI ^9^ or requiring inotropes, ventilation or intra-aortic balloon pump) were excluded. Emergency or salvage procedures were also excluded.

### Sampling

Atrial biopsies (30 – 100 mg) were collected prior to cardiopulmonary bypass from the right atrium auricle from patients that fasted at least eight hours before the surgery. Samples were immediately snap-frozen in liquid nitrogen, split for RNA isolation and metabolomics analysis and stored at −80°C.

### Outcomes

Levels of metabolites and transcripts in atrial biopsies.

### RNA isolation and sequencing

RNA was isolated from 20mg of tissue using ISOLATE II RNA Mini Kit (bioline, London, UK). Sample quality was assessed using the RNA ScreenTape assay on the Agilent Tapestation 4200. Only samples with RNA integrity number equal to or greater than eight were sequenced.

Library preparation and sequencing was carried out in two batches by Source BioScience (Nottingham, UK). The Stranded total RNA libraries were prepared in accordance with the Illumina TruSeq Stranded Total RNA Sample Preparation Guide with Ribo-Zero Human/Mouse/Rat for Illumina Paired-End Multiplexed Sequencing. The libraries were validated on the Agilent BioAnalyzer 2100 to check the size distribution of the libraries and on the Qubit High Sensitivity to check the concentration of the libraries. Sequencing was performed using 75bp paired-end chemistry on HiSeq 4000 with the TruSeq Stranded Total RNA Human kit.

### Metabolomics

A panel of 144 metabolites involved in mitochondrial function and energy metabolism were analyzed using a targeted assay on a Thermo Quantiva interfaced with a Vanquish Liquid Chromatography System as previously described in. ^10,11^ In brief, tissue was extracted using a modified Folch extraction into chloroform/methanol (2:1 600 ul per 50 mg of tissue, followed by 200 ul of water, 200 ul of chloroform, repeated once). For nucleotides and acyl-CoA derivatives one half of the aqueous extract was dissolved in 150 µl of 70:30 acetonitrile:water containing 20 µM deoxy-glucose 6 phosphate and 20 µM [U–13C, 15N] glutamate. The resulting solution was vortexed, sonicated and centrifuged. Chromatography consisted of a strong mobile phase (A) was 100 mM ammonium acetate, and weak mobile phase was acetonitrile (B) and the LC column used was the ZIC-HILIC column from SeQuant (100 mm × 2.1 mm, 5 µm).

For amino acids and TCA cycle intermediates aqueous extracts were reconstituted in 50 μl of 10 mmol/l ammonium acetate in water before TCA cycle intermediates were separated using reversed-phase liquid chromatography on a C18-PFP column (150 mm × 2.1 mm, 2.0 μm; ACE). For chromatography on the UHPLC system, mobile phase A was 0.1% formic acid in water, and mobile phase B was 0.1% formic acid in acetonitrile. Mass transitions of each species were as follows (precursor > product): D5-L-proline 121.2 > 74.2; D8-L-valine 126.1 > 80.2; D10-L-leucine 142.0 > 96.2; L-glutamate [M] 148.0 > 84.2; L-glutamate [M+1] 149.0 > 85.2; L-glutamate [M+6] 154.1 > 89.1; citrate 191.0 > 111.0; citrate [M+1] 192.0 > 112.0; citrate [M+2] 193.0 > 113.0; citrate [M+3] 194.0 > 114.0; citrate [M+4] 195.0 > 114.0; citrate [M+5] 196.0 > 115.0; citrate [M+6] 197.0 > 116.0. Collision energies and radio frequency (RF) lens voltages were generated for each species using the TSQ Quantiva optimization function.

### Data processing and statistical analysis

#### Transcriptomics

Sequencing data were quality-checked with Fastqc v0.11.5, ^12^ quantified with Salmon v1.21 ^13^ after indexing and annotating with Gencode34 (Ensembl v100) reference genome and transcriptome files. Transcript quantities were normalized to length-scaled transcripts per million and filtered to retain only high quantities (filterByExpr function in edgeR) before downstream analysis using limma-voom model ^14^ with empirical Bayes moderation ^15^ with batch accounted for. The false discovery rate was set at 5%. Pathway enrichment in the dataset was tested with *camera* function (*limma*) ^16^ using Reactome ^17^ (protein-coding transcripts) and Gene Ontology ^18,19^ (lncRNA) annotations.

The biphasic BMI relationship of gene and metabolite expression was tested with the Two Line algorithm, ^20^. Weighted gene correlation networks analysis (WGCNA) was performed with the WGCNA R package. ^21^ These analyses were performed on log-transformed and quantile normalised expression values with batch effect removed as they are not multivariate models.

#### Metabolomics

Peak area ratios of metabolites were obtained by integration within vendor software (Xcalibur QuanBrowser, Thermo Scientific, Hemel Hempstead, UK) and compared with isotopically labelled standards for quantification. Data for metabolites where no-expression values were greater than 20% were removed from the analysis. Further pre-processing included filtering features of constant value and extreme relative standard deviation or coefficient of variation, as well as missing value replacement using MetaboAnalystRv2. ^22^ The processed data was log-normalized and Pareto-scaled. Pairwise comparisons of sample groups were carried out using cross-validated PLSDA and t-test. Metabolites were considered differential where variable importance in projection scores (VIP) >1 and t-test p-value < 0.05.

Sample separation was visualized using principal component analysis plot of normalized transcriptome and metabolite data with R base and ggplot2. ^23^

#### Multiomics analyses

RNA and metabolite were combined using block sparse PLSDA models using mixOmics v0.6 ^24^. Canonical correlation patterns and association networks derived from the model components were then used to infer relationship among genes and metabolites. Network visualization was carried out with Cytoscape. ^25^

### Data Availability

Sequencing and samples group data are available via NCBI Gene Expression Omnibus (GSE159612).

## Results

### Study cohort and group definition

The ObCard study (NCT02908009) screened 558 participants between August 2017 and May 2019, out of which 221 were approached. Sixty-eight consecutive patients were recruited to the study, with one protocol violation recorded and one participant withdrew from the study (**Figure 1A**). The distribution of BMI in the sample is shown in **Figure 1B**.

**Figure 1.**
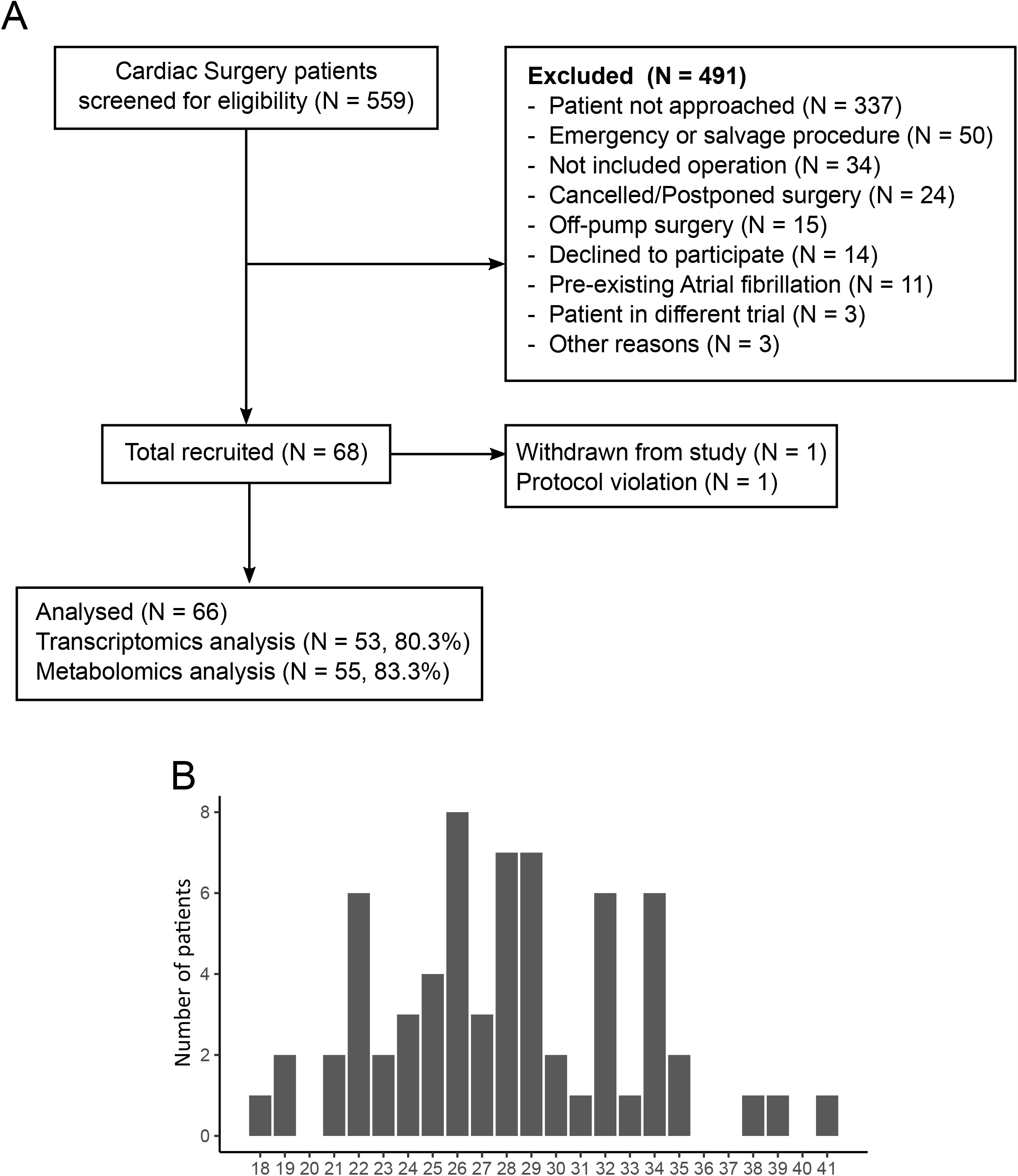
(**A**) Consort diagram, (**B**) BMI distribution in the cohort.

Out of 66 collected samples, we analyzed the transcriptome in 53 and metabolites in 56 myocardial biopsies. (**Figure 1A** and **Table S1**). Metabolomics data for one sample was not included in the analysis because we were not able to detect majority of the metabolites.

After data processing, 14,967 transcripts were identified as protein-coding, 2,102 were long non-coding RNAs (lncRNA), 662 were pseudogenes, and the remaining 239 transcripts included unknown transcripts, micro-RNAs, rRNA and other non-coding RNAs (**Figure 2A**). All samples were very similar in transcript biotype composition (**Figure 2B**).

**Figure 2.**
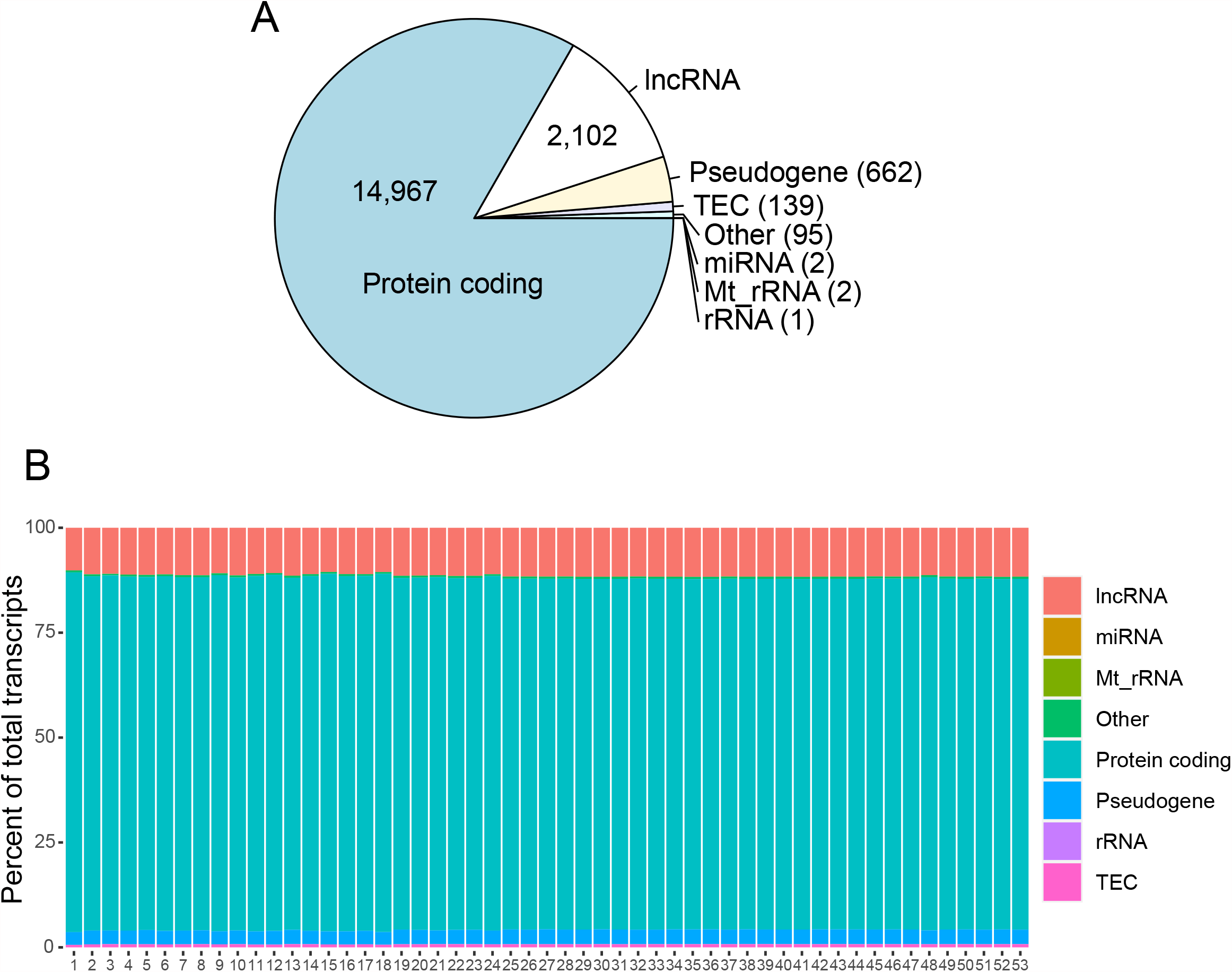
(**A**) Summary of transcript biotypes in the whole transcriptomics dataset. (**B**) Biotypes percentage by sample

To identify transcripts and metabolites that display biphasic u- or n-shape BMI relationships, we used the Two Lines method developed by Simonsohn. ^20^ The algorithm does not impose an assumption of form on the data, which overcomes a limitation of quadratic regression known to lead to false positives. The Two Lines method tests two regression lines and detects a breakpoint between them. The analysis identified 239 transcripts with a biphasic relationship, out of which 107 had their local minimum or maximum at BMI 28 or 29 (**Figure 3A** and **Table S2**). None of the analyzed metabolites displayed a biphasic BMI dependence.

**Figure 3.**
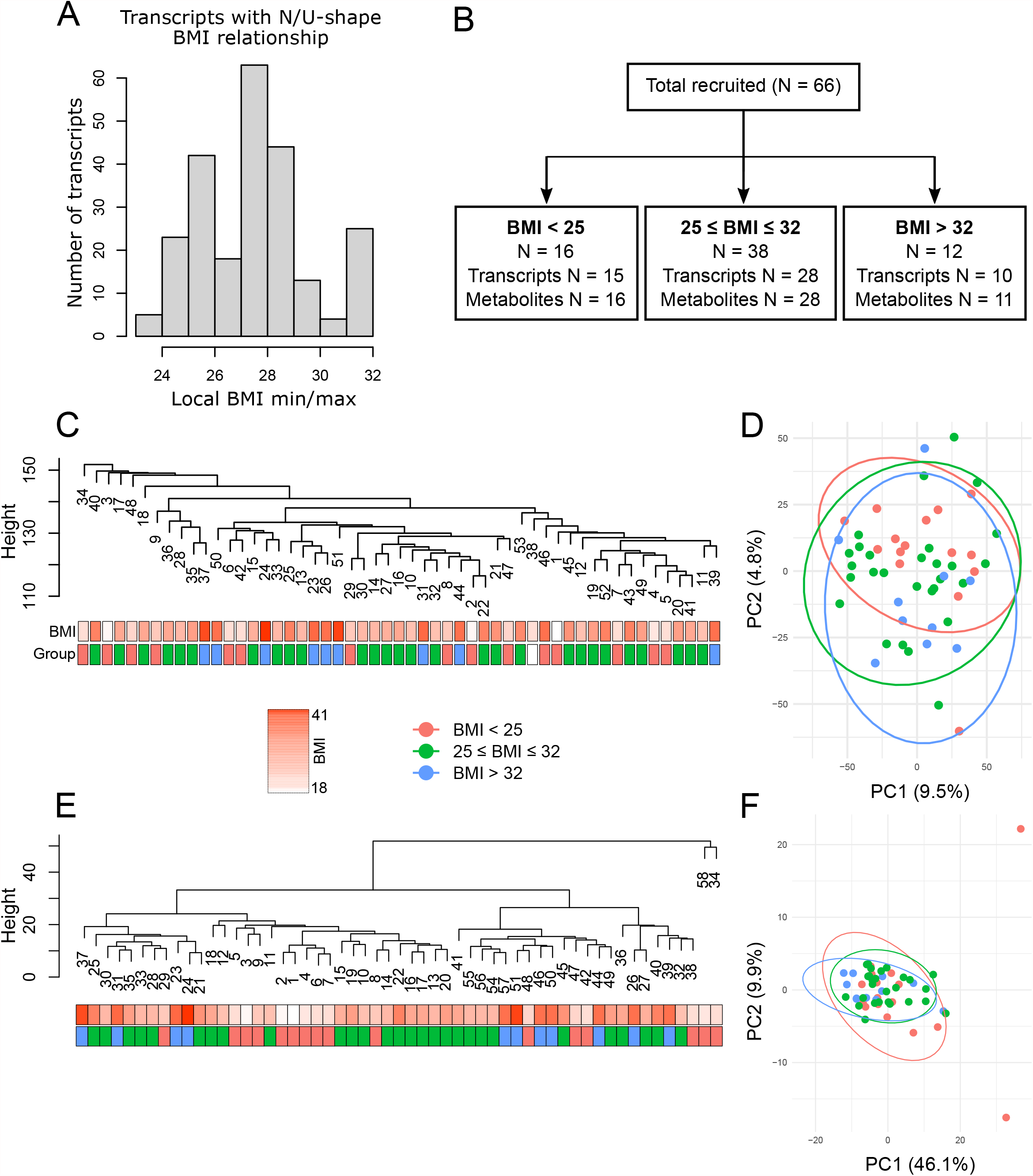
(**A**) Distribution of local BMI minima and maxima for transcripts whose expression display U or N-shape BMI dependence. (**B**) BMI groups definition with numbers of samples. (**C**) Hierarchical clustering of samples using transcriptomics data. (**D**) Principal component analysis of transcriptomics data: the first two components were plotted with 95% confidence interval for each BMI group. (**E**) Hierarchical clustering of samples using metabolomics data. (**F**) Principal component analysis of metabolomics data: the first two components were plotted with 95% confidence intervals for each BMI group.

Given the limitations of BMI as a measure of obesity and the *ad hoc* nature of the WHO BMI obesity classifications we adopted a data-driven approach to group participants. Based on the analysis of transcripts, we defined the group that included local minima or maxima of gene expression as BMI 28/29 ± 3, range 25 – 32 (**Figure 3B**). The other two groups were BMI<25 and BMI>32. The three patient groups were well matched with respect to baseline demographics, clinical characteristics, medications, and operative procedures (**Table 1**) apart from post-surgery worst multiple organ dysfunction score (MODS, higher if BMI ≥25) and non-red blood cells transfusion (highest if BMI <25) that were different between groups. Levels of missing data were low (**Table 1**). The analysis cohort included 53 participants (80.3%) with complete transcriptome data and 55 participants (83.3%) with complete metabolomics data (**Table S1**). Baseline characteristics in the complete case cohort were comparable to the analyzed cohort (**Table S3**).

**Table 1.** Pre- and Post-operative characteristics. (*) - Tests among BMI groups were conducted by exact test for categorical variables, and ANOVA or non-parametric Kruskal-Wallis test for continuous variables. Data are presented as n (%) for categorical variables and mean (standard deviation, STD) or median (interquartile range, IQR) for continuous variables. Abbreviations: ACE – Angiotensin Converting Enzyme; AKI – Acute Kidney Injury; BMI – Body Mass Index; CABG – Coronary artery Bypass Grafting; CCS – Canadian Cardiovascular Society; Hct – Haematocrit; FiO2 – Fraction of Inspired Oxygen; KDIGO - The Kidney Disease Improving Global Outcomes; MABP – Mean Arterial Blood Pressure; MODS – Multiorgan Dysfunction Syndrome; NYHA – New York Heart Association; PO2 – Partial Pressure of Oxygen; RBC – Red Blood Cells; VD – Vessel Disease.

| **All (n=66)** | **BMI<25**<br>**(n=16)** | **25≥BMI≤32**<br>**(n=33)** | **BMI>32**<br>**(n=11)** | **p value** | **Missing**<br>**data (n)** |
| --- | --- | --- | --- | --- | --- |
| Age (years) - Median (IQR) | 68 (52 - 82) | 68 (50 - 79) | 62 (51 - 72) | 0.277 | 0 |
| Sex (male) - n (%) | 14 (88%) | 32 (84%) | 11 (92%) | 0.999 | 0 |
| Ethnic (White) - n (%) | 14 (88%) | 32 (86%) | 12 (92%) | 0.999 | 0 |
| BMI | 21.8 (1.8) | 28.2 (2.5) | 35.4 (2.3) | <0.001 | 0 |
| *Smoking History* |  |  |  |  |  |
| Never smoker - n (%) | 6 (38%) | 15 (39%) | 4 (33%) |  |  |
| Ex-smoker - n (%) | 8 (50%) | 20 (53%) | 6 (50%) | 0.908 | 0 |
| Current smoker - n (%) | 2 (13%) | 3 (8%) | 2 (17%) |  |  |
| *Medical history* |  |  |  |  |  |
| Diabetes - n (%) | 2 (13%) | 11 (29%) | 4 (33%) | 0.403 | 0 |
| Permanent Pacemaker - n (%) | 1 (6%) | 2 (5%) | 0 (0%) | 0.999 | 0 |
| Stroke/Transient Ischaemic Attack - n (%) | 2 (13%) | 3 (8%) | 1 (8%) | 0.844 | 0 |
| Chronic pulmonary disease - n (%) | 3 (19%) | 4 (11%) | 3 (25%) | 0.366 | 0 |
| Neurological disease - n (%) | 0 (0%) | 0 (0%) | 0 (0%) | N/A | 0 |
| Renal disease - n (%) | 0 (0%) | 1 (3%) | 2 (17%) | 0.126 | 0 |
| Myocardial infarction - n (%) | 5 (31%) | 11 (29%) | 1 (8%) | 0.318 | 0 |
| Extracardiac arteriopathy - n (%) | 2 (13%) | 4 (11%) | 1 (8%) | 1.000 | 0 |
| Liver disease - n (%) | 0 (0%) | 0 (0%) | 0 (0%) | N/A | 0 |
| Pulmonary hypertension - n (%) | 0 (0%) | 1 (3%) | 0 (0%) | 1.000 | 0 |
| *Pre-operative Medication History* |  |  |  |  |  |
| Statin - n (%) | 12 (75%) | 28 (74%) | 11 (92%) | 0.477 | 0 |
| Anti-platelet agents - n (%) | 11 (69%) | 32 (84%) | 11 (92%) | 0.255 | 0 |
| ACE inhibitors - n (%) | 9 (56%) | 14 (37%) | 5 (42%) | 0.487 | 0 |
| *Clinical characteristics* |  |  |  |  |  |
| *Surgery type* |  |  |  |  |  |
| CABG only - n (%) | 13 (81%) | 32 (84%) | 11 (92%) |  |  |
| CABG & Valve - n (%) | 3 (19%) | 5 (13%) | 1 (8%) | 0.878 | 0 |
| Others - n (%) | 0 (0%) | 1 (3%) | 0 (0%) |  |  |
| NYHA |  |  |  |  |  |
| Class I - n (%) | 4 (25%) | 12 (32%) | 4 (33%) |  |  |
| Class II - n (%) | 11 (69%) | 23 (61%) | 6 (50%) | 0.813 | 0 |
| Class III, IV - n (%) | 1 (6%) | 2 (5%) | 3 (25%) |  |  |
| CCS |  |  |  |  |  |
| Asymptomatic - n (%) | 5 (31%) | 2 (5%) | 2 (17%) |  |  |
| Class I - n (%) | 6 (38%) | 12 (32%) | 4 (33%) | 0.228 | 0 |
| Class II - n (%) | 4 (25%) | 19 (50%) | 5 (42%) |  |  |
| Class III, IV - n (%) | 1 (6%) | 5 (13%) | 1 (8%) |  |  |
| Left Ventricular Ejection Fraction |  |  |  |  |  |
| Good (>49%) - n (%) | 13 (81%) | 31 (82%) | 9 (75%) | 0.912 | 0 |
| Fair (30-49%) - n (%) | 3 (19%) | 7 (18%) | 3 (25%) |  |  |
| Left main stem disease - n (%) | 2 (13%) | 9 (24%) | 2 (17%) | 0.761 | 0 |
| Extent of coronary disease |  |  |  |  |  |
| Normal/ 1VD - n (%) | 1 (6%) | 2 (5%) | 3 (25%) |  |  |
| 2VD - n (%) | 7 (44%) | 7 (18%) | 2 (17%) | 0.066 | 0 |
| 3VD - n (%) | 8 (50%) | 29 (76%) | 7 (58%) |  |  |
| Pre-operative PaO2/FiO2 ratio - Median (IQR) | 533 (445 - 691) | 457 (410 - 533) | 457 (410 - 495) | 0.399 | 8 |
| Pre-operative Platelets count (x10<sup>9</sup>/L) - Mean (STD) | 222.3 (55.7) | 229.3 (63.3) | 227.5 (62.0) | 0.928 | 1 |
| Pre-operative Serum Creatinine (umol/L) - Median (IQR) | 78.5 (70.6 - 102.5) | 79.5 (69 - 89.3) | 86.0 (77.5 - 96.5) | 0.483 | 0 |
| Pre-operative Bilirubin (umol/L) - Median (IQR) | 10.0 (7.5 - 12.5) | 11.0 (8 - 13) | 8.5 (8 - 13.5) | 0.754 | 5 |
| *Postoperative* |  |  |  |  |  |
| Hct (%) - Mean (STD) | 31.9 (3.9) | 34.2 (4.0) | 35.5 (4.9) | 0.069 | 0 |
| MABP (mm Hg) - Median (IQR) | 76 (68.8 - 85.0) | 72.5 (64.3 - 75.8) | 67.5 (63.3 - 73.5) | 0.739 | 0 |
| Lactate (mmol/L) - Median (IQR) | 1.5 (1.1 - 2.0) | 1.9 (1.4 - 2.3) | 1.5 (1.1 - 1.9) | 0.985 | 1 |
| Inotropic score at 24h - Median (IQR) | 0 (0 - 2) | 1.5 (0 - 3) | 0 (0 - 3) | 0.282 | 7 |
| Vasoactive score at 24h - Median (IQR) | 5 (2.5 - 9) | 6 (3 - 8) | 3 (1 - 7) | 0.426 | 4 |
| MODS ICU - Median (1st - 3rd quartile) | 1 (1 - 3) | 2 (2 - 3) | 3 (1.8 - 3) | 0.122 | 2 |
| Worst postoperative MODS - Median (IQR) | 2.5 (1 - 5) | 3 (2 - 4) | 3 (2.5 - 4) | 0.048 | 1 |
| PaO2/FiO2 ratio at 48h - Median (IQR) | 410 (342 - 573) | 358 (307 - 410) | 433 (359 - 460) | 0.144 | 1 |
| Serum creatinine 48h (umol/L) - Median (IQR) | 73 (67.5 - 90.0) | 75.5 (60.3 - 84.0) | 74.5 (72.3 - 78.3) | 0.390 | 2 |
| RBC transfused postoperative - n (%) | 8 (50%) | 15 (39%) | 2 (17%) | 0.186 | 0 |
| nonRBC transfusion at more than 48h - n (%) | 2 (13%) | 4 (11%) | 1 (8%) | 0.999 | 0 |
| nonRBC transfusion within 48h -<br>n (%) | 6 (40%) | 7 (18%) | 0 (0%) | 0.031 | 1 |
| PaO2/FiO2 ratio at 48hr <=300 -<br>n (%) | 3 (19%) | 7 (18%) | 2 (17%) | 0.999 | 1 |
| AKI according to kdigo criteria -<br>n (%) | 1 (6%) | 1 (3%) | 0 (0%) | 0.807 | 1 |

### Transcriptomics and metabolomics data analysis

Hierarchical clustering and principal component analyses were performed on log-transformed and quantile normalized transcriptomics data with batch effect removed. Neither analysis revealed any clear differences between groups when all transcriptomics data were included (**Figure 3C, D**). However, only 15.3% of sample variability could be explained indicating that the majority of the heterogeneity is not captured by the first two components (**Figure 3D**). Clustering of the metabolomics data also did not show any clear differences between groups (**Figure 3E, F**). Instead, it identified samples 34 and 58 as clear outliers. These samples were removed from further analysis. Differential expression (DE) analysis identified only one significantly different transcript (AC006059.2, logFC=-3.91, adjusted p-value=0.008) between BMI>32 and 25≥ BMI ≤ 32 groups and also across all groups (adjusted p-value=0.018). AC006059.2 encodes an uncharacterized protein within the hypoxia-induced gene 1 domain.

To determine differences between groups of transcripts, we performed a differential pathway analysis using competitive gene set testing as described by Wu and Smyth ^16^ to identify groups of transcripts annotated to a specific pathway (Reactome and Gene Ontology) that behave differently between the BMI groups. Out of 63 identified pathways (adjusted p-value<0.05), 29 were related to translation and mRNA processing (over 300 transcripts, **Figure 4A** and **Table S4**). These pathways were downregulated in both the 25≥ BMI ≤ 32 and BMI >32 groups as compared with the BMI <25 group. Transcripts involved in striated muscle contraction (29 transcripts; **Figure 4A** and **Table S4**) behaved in an opposite manner. They were expressed at higher levels in both 25≥ BMI ≤ 32 and BMI >32 groups as compared with the BMI<25 group. In addition, cholesterol biosynthesis (24 transcripts), and innate immunity pathways like FCGR activation (58 transcripts) or creation of C2 and C4 activators (56 transcripts) were upregulated in the BMI >32 group as compared with the 25≥ BMI ≤ 32 (**Figure 4A** and **Table S4**). Ribosomal proteins L3, L4 and L6, signal peptidase complex catalytic subunit (SEC11C), SMG5 and proline-rich nuclear receptor coactivator 2 (PNRC2) were the most variable between the groups (p-value < 0.05, **Table S5**) and among translation-related transcripts. These transcripts also displayed a biphasic u-shape (or n-shape, SMG5) BMI relationship (**Figure 4B**). Other transcripts that had membership in significant pathways and displaying biphasic BMI dependence are plotted in **Figure S1**. These include CARS1 (translation), UPF3A, PPP2R1A (nonsense-mediated decay), NHP2, THUMPD1 (rRNA processing), IGKV2-28, FCN3 (creation of C4 and C2 activators), OAS3 (interferon signalling), KYAT3, RSAD2 (metabolism of amino acids) and TNNT2 (striated muscle contraction).

**Figure 4.**
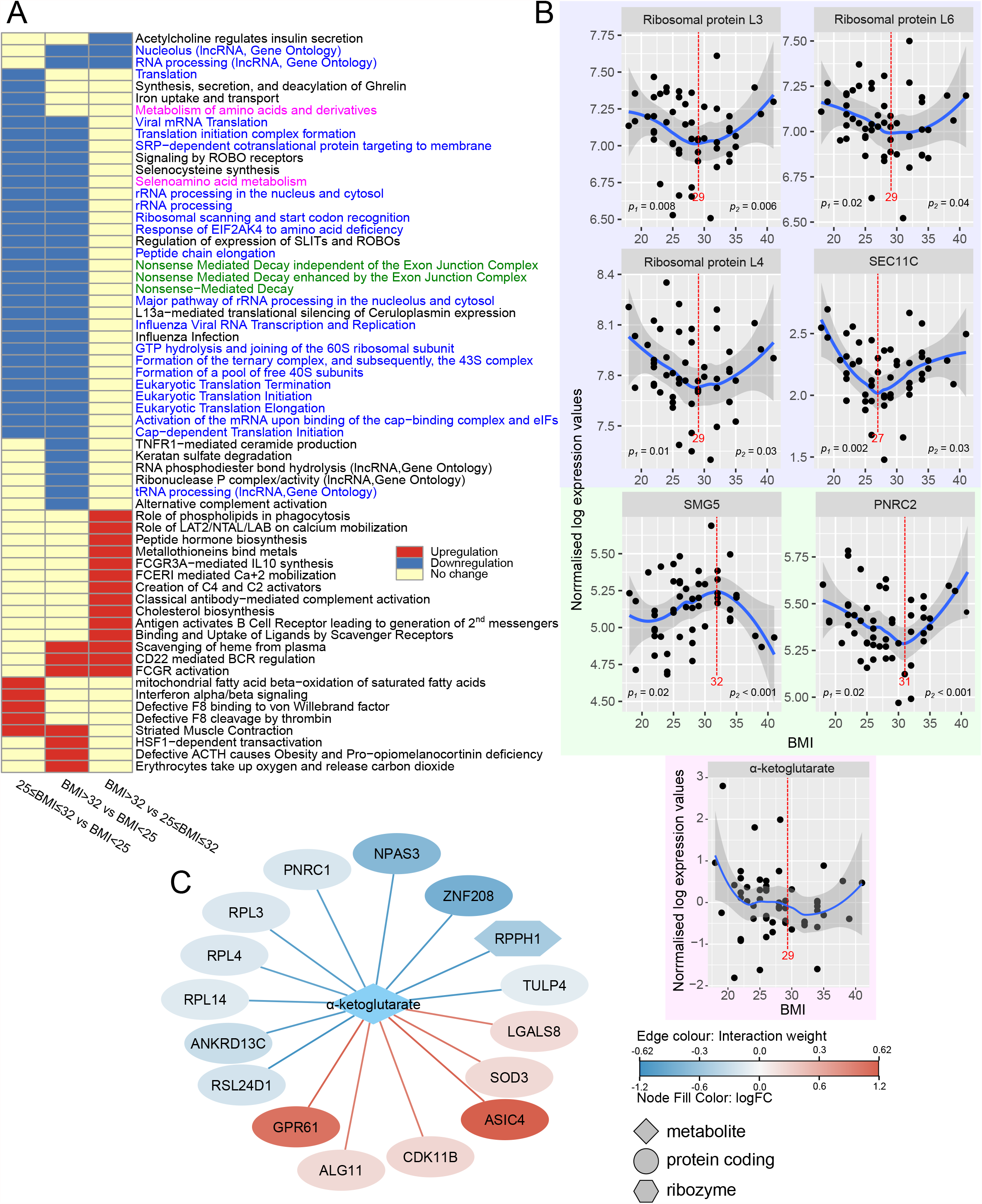
(**A**) Significant pathways plotted as a heatmap. Green and blue color show pathways related to translation and RNA processing. Magenta indicates amino acids metabolism pathways that include - ketoglutarate. (**B**) Highly variable transcripts between the BMI groups with the membership in significant pathways and showing biphasic BMI relationship are plotted against BMI. ✓-ketoglutarate that is significantly different between the 25≥ BMI ≤ 32 and BMI<25 groups was also plotted. The red line and number indicate local minimum or maximum; p1 and p2 specify regression p-values before and after the local minimum/maximum. (**C**) ✓-ketoglutarate – transcripts similarity network. The colour of edges indicates the interaction weight and node colour indicates log fold change in the 25≥ BMI ≤ 32 vs BMI<25 comparison. Node shape indicates transcripts’ biotype.

Metabolite analysis identified α-ketoglutarate as significantly upregulated in the 25≥ BMI ≤ 32 group when compared with the BMI <25 group (**Table 2**). α-ketoglutarate also has membership in one of the significant pathways, i.e., selenoamino acid metabolism (**Figure 4A**). Two Lines analysis of α-ketoglutarate expression did not identify significant u or n-shape BMI dependence (**Figure 4B**). However, metabolite-transcripts association analysis found its expression patterns similar to some of the transcripts with annotations in translation (RPL3, RPL4, RPL14, RSL24D1) and RNA processing (PNRC1 and RPPH1, **Figure 4C**). Long and middle chain acylcarnitines expressed at highest levels in the BMI >32 group as compared with both the 25≥ BMI ≤ 32 and BMI <25 groups. There was no significant difference between the BMI <25 and 25≥ BMI ≤ 32 groups. Two line analysis indicated that acylcarnitines significantly increased in patients with BMI greater than 25 (**Figure S2** and **Table S2**). Four out of eight acylcarnitines (C16-OH, C18-OH, C18:1-OH, C18:2-OH) were hydroxylated. Their unhydroxylated counterparts (apart from carnitine C18) did not show any significant BMI dependence (**Table S2**). Ribose-5-phosphate was also significantly upregulated in the BMI >32 group as compared with the BMI <25 group and its expression increased significantly with BMI (**Figure S2** and **Table S2**).

**Table 2.**
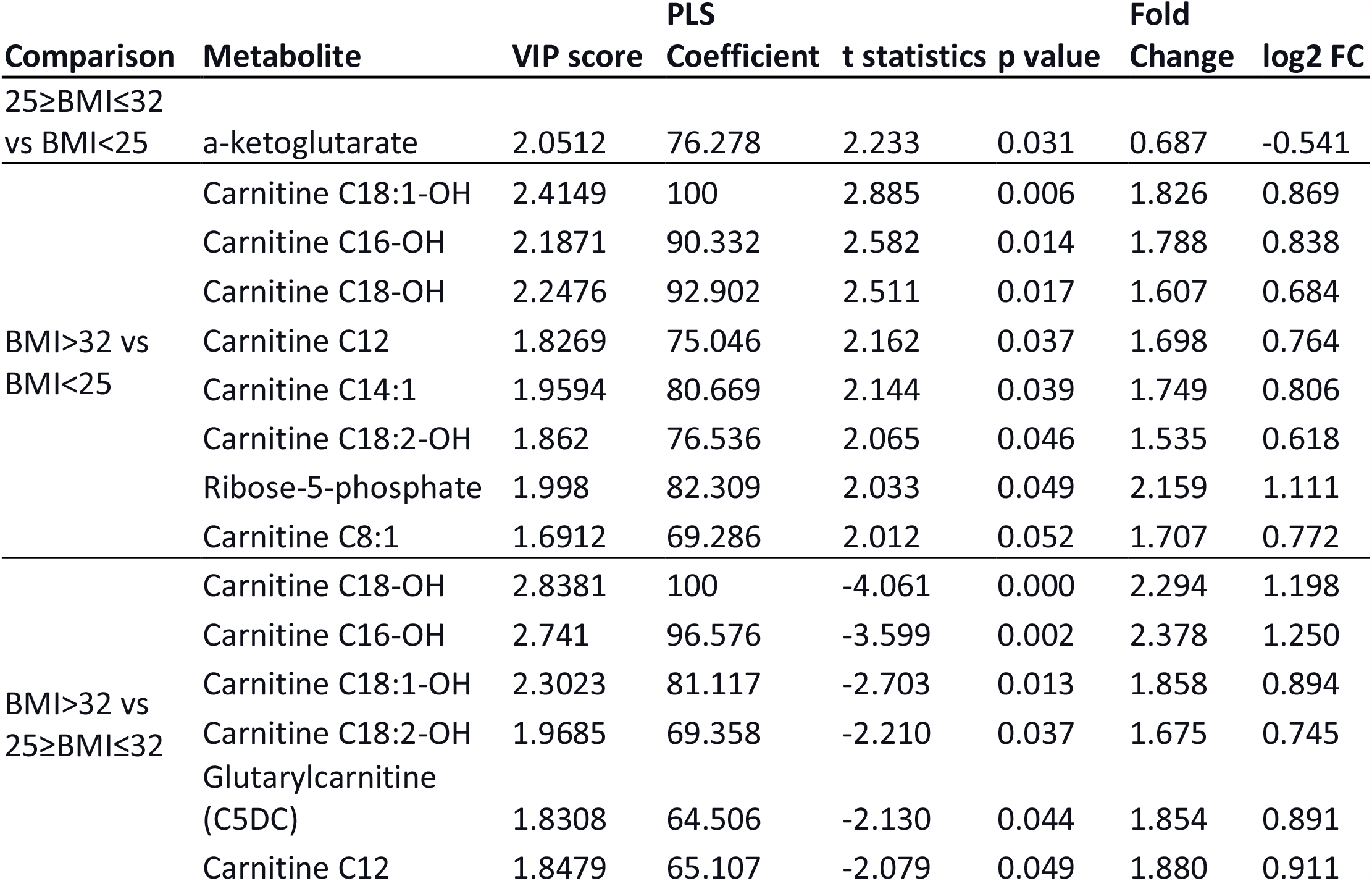
Differentially expressed metabolites between analyzed groups. VIP score - Variable Importance in Projection in the Partial Least Square Discriminant Analysis, PLS Coefficient - regression parameters in the Partial Least Square model. Positive log2 fold change (log2 FC) indicates higher expression in the group with the higher BMI.

Next, we examined whether groups of genes without any form of pathway information were differential, and whether such groups show biphasic BMI relationship. We explored correlations between expression patterns of 429 transcripts that were most variable between the three BMI groups (p-value < 0.05, **Table S5**) using weighted gene correlation networks analysis. The analysis grouped the transcripts into twelve networks. Eigengene values of five showed either u or n shape BMI relationship (Turquoise, Purple, Grey, Green and Yellow in **Figure 5A**) and eigengene values of one network significantly increased with BMI across the whole range (Black in **Figure 5A**). The networks’ eigengene values for each of the six networks were different between the three BMI groups as indicated by one-way ANOVA (**Figure 5B**).

**Figure 5.**
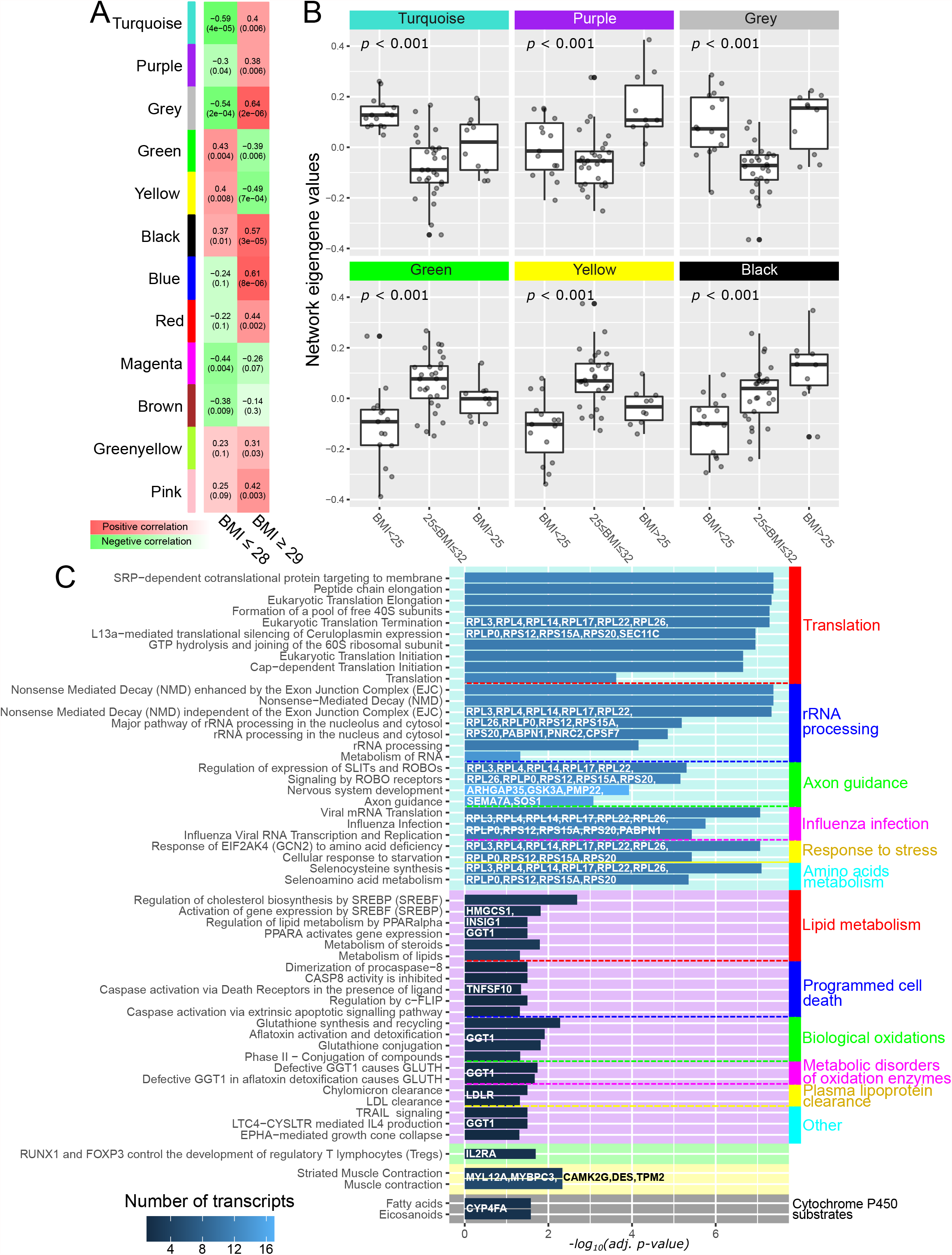
(**A**) Correlations between transcript networks’ eigengene values and BMI. Values is brackets are false discovery rate adjusted p-values. (**B**) Boxplots of networks’ eigengene values in the BMI groups with one-way ANOVA p-values. (**C**) Reactome pathway enrichments with network-specific genes. Color intensity of the bars indicate number of genes from each network that enriched particular pathways. The length of the bars indicates –log10(p-value). Only pathways with false discovery rate adjusted p-values less than 0.05 are shown.

To identify the biological function of the six significant networks, we submitted the specific transcripts to Reactome pathway enrichment tool. ^17^ Risosomal protein transcripts from the Turquoise network significantly (adjusted p-value < 0.05) enriched translation and RNA processing pathways; transcripts from the Purple network (Hydroxymethylglutaryl-CoA synthase, Insulin-induced gene 1, Glutathione hydrolase 1, Tumor necrosis factor ligand superfamily member 10 and Low-density lipoprotein receptor) enriched lipids and steroid metabolism, biological oxidations, and apoptotic pathways. Transcripts from the Yellow network enriched muscle contraction pathways with tropomyosin β-chain, desmin, myosin regulatory light chain (MYL12A), myosin-binding protein C and Ca^2+^/calmodulin-dependent protein kinase type II subunit gamma (CAMK2G). Interleukin-2 receptor alpha chain from the Green network enriched ‘RUNX1 and FOXP3 control the development of Treg’. Cytochrome P450 (CYP4FA) present in the black network enriched ‘Cytochrome P450 substrates’ (**Figure 5C**). The Grey network included ten transcripts that were not clustered in any other network and, as expected, did not significantly enrich any pathway.

## Discussion

### Main findings

The obesity paradox refers to a u-shaped biphasic relationship between BMI and mortality in people exposed to acute metabolic stress including cardiac surgery. ^2^ In the current analysis we show that groups of myocardial transcripts involved in translation and muscle contraction show similar biphasic BMI dependence; specifically the expression of genes involved in muscle contraction were greatest, and the expression of genes associated with translation were lowest in the 25≥ BMI ≤ 32 group. In addition, BMI >32 was associated with upregulated innate immunity and cholesterol synthesis. These results were consistent across both competitive gene set analyses, and weighted gene correlation networks analysis. Contrary to our original hypothesis, metabolites involved in energy production did not demonstrate biphasic BMI dependence. Instead, acylcarnitines and ribose-5-phosphate increased together with BMI. Our findings are summarized in **Figure 6**.

**Figure 6.**
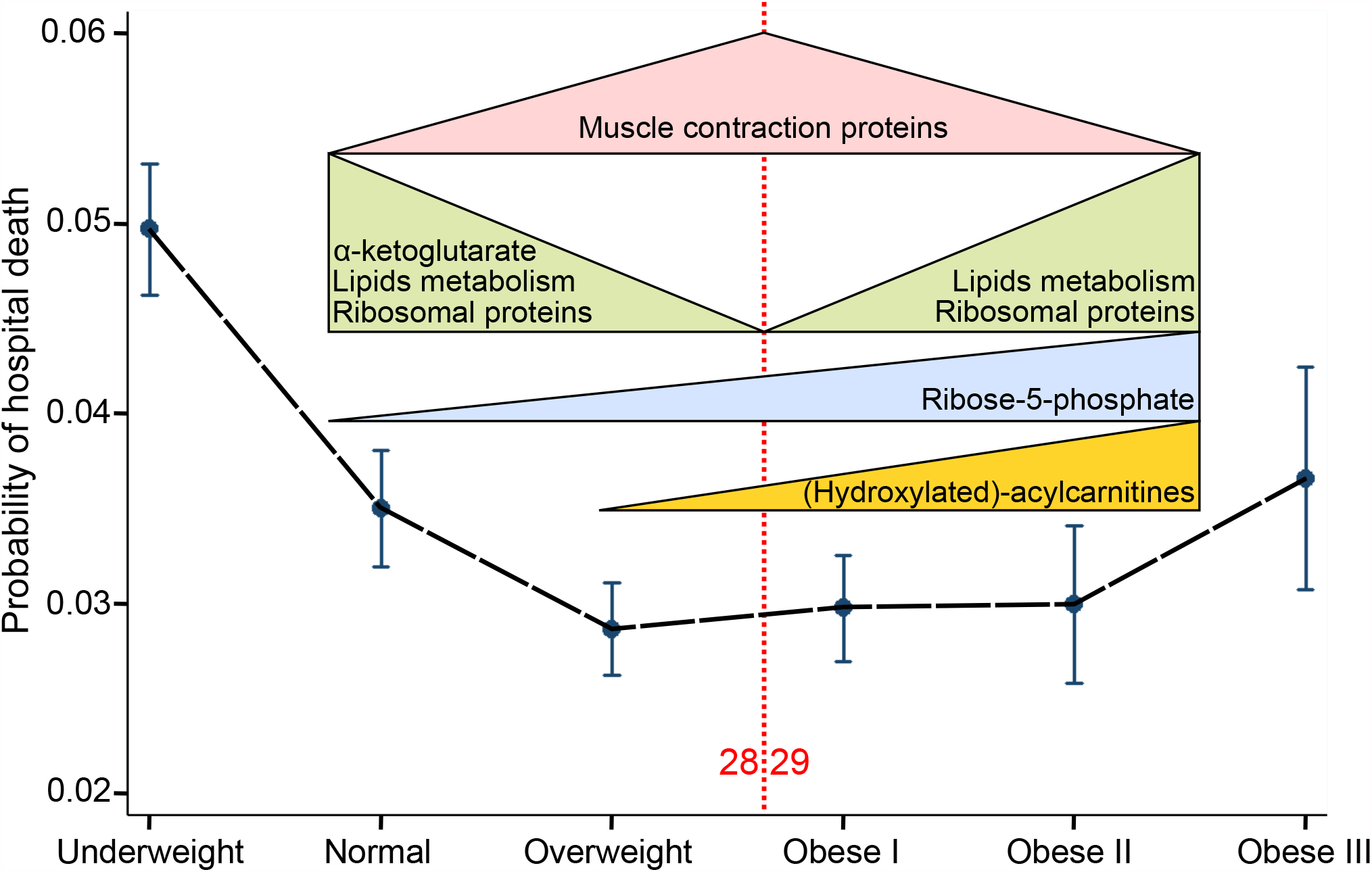
Summary of the results overlaid on probability of hospital death in WHO BMI groups. ^2^

### Clinical Importance

These findings provide new evidence that BMI is associated with molecular differences in human myocardium that mirror the clinical biphasic relationship between BMI and mortality in the obesity paradox. These findings do not prove causality between obesity and improved outcomes, neither do they disprove unmeasured confounding or reverse epidemiology as explanations of the obesity paradox. Rather they support the hypothesis that the obesity paradox is attributable to an underlying molecular mechanisms, and suggest that further explorations of the therapeutic potential of these findings are warranted.

A novel finding of this study is that the expression of translation-related genes showed biphasic BMI dependence. However, it is not clear why this would provide an advantage to the 25≥ BMI ≤ 32 participants. Sarcopenia and frailty commonly observed in people who are under- and normal weight, or who have severe obesity, are more often associated with reductions in protein synthesis. Importantly, levels of protein synthesis were not measured in this analysis, so the results do not allow us to speculate on whether this was influenced by the observed global differences in translation. Changes in protein translation may have multiple other effects on cell metabolism. For example changes in ribosomal proteins influence processes like regulation of gene expression, cell cycle control, apoptosis, cell migration in angiogenesis, and intimal thickening (reviewed in ^26^). Dysregulation of ribosomal proteins expression is also a characteristic of cancer progression. ^27^ In the current study, changes in the expression of ribosomal proteins in 25≥ BMI ≤ 32 patients appear to regulate the expression of specific transcripts. Out of 104 transcripts in the Turquoise network (**Figure 5)**, at least thirteen have a clear function in translation-related processes. These results warrant further investigation of translation-related and associated transcripts and proteins to determine whether they could explain the apparent survival benefits of overweight patients in cardiac surgery.

The expression of genes involved in muscle contraction were increased 1.05 – 1.6-fold in the 25≥ BMI ≤ 32 group compared with BMI<25 and BMI >32 groups. These included tropomyosin β-chain, desmin, myosin regulatory light chain (MYL12A), myosin-binding protein C and Ca^2+^/calmodulin-dependent protein kinase type II subunit gamma (CAMK2G). Evidence indicates that changes in the expression of proteins responsible for muscle contraction lead to cardiomyopathies. Mutations in myosin regulatory light chains associate with disruptions of contractility and hypertrophy. Some of the mutations disrupt phosphorylation sites of the myosin regulatory light chains that is proposed to be mediated by Ca^2+^/calmodulin-dependent protein kinase and essential for muscle contraction. ^28^ Mutations in myosin-binding protein C also leads to hypertrophy ^29^ and decreased desmin expression levels associated with the progression of heart failure. ^30^ Imaging studies demonstrate reduced contractility in the atria of people with severe obesity, with remodeling and improved contractility following weight loss interventions. ^31^ These observations support the further evaluation of pre-surgery weight loss as an organ protection strategy in people with severe obesity.

The muscle contraction transcripts were linked with 26 other transcripts that included tRNA methyltransferase O (TRMO) and SMG5 nonsense mediated mRNA decay factor that are involved in RNA processing. These transcripts can potentially form a regulatory network controlling expression of muscle contraction. A question appears whether there is a functional relationship between expressions of ribosomal and muscle contraction genes.

The metabolite analysis identified several long and middle-length acylcarnitines that were significantly increased in the BMI>32 group. The longest-chain acylcarnitines were hydroxylated (C16-OH,C18-OH, C18:1-OH and C18:2-OH). This type of modification of acylcarnitines was recently identified as a biomarker of mitochondrial myopathy. ^32^ Their unhydroxylated counterparts did not change significantly between the groups, however, the significance of this finding can be biased by the fact that increasing BMI positively correlates with circulating lipid levels. In this study, we did not measure plasma metabolites to adjust for that. Also ratios of hydroxylated/unhydroxylated acylcarnitines were not statistically different. Apart from acylcarnitines, we did not detect any significant difference in expression of glycolytic and oxidative phosphorylation metabolites. The reason can be the fact that there was no significant difference between the groups in the incidence of diabetes and heart failure between the groups, which have clear associations with BMI and where the role of mitochondria is clearly established ^33^. Alternatively, it is possible that the differences become apparent after surgery. Overall, the results do not allow us to speculate on the role of mitochondria in the obesity paradox. Nevertheless, the levels of α-ketoglutarate, involved in branched amino acids catabolism in mitochondria, were increased in the BMI<25 group and its expression patterns were similar to ribosomal proteins. The significance of this finding needs further investigation.

The study has several limitations. First, the sample size was small. As a result we could not detect significant differences in single gene expression. Second, the samples collected during surgery also potentially differed in the percentage of cardiomyocytes and could include fragments of blood vessels or fat tissue. Due to small biopsy sizes (30-100mg), it was impossible to assess the cell-type heterogeneity of the samples. Third, we recruited a very low risk cohort to the analysis; rates of acute kidney injury, lung injury, and low cardiac output were very low. This, along with the small sample size reduced our ability to demonstrate differences in clinical outcomes between the three groups.

In summary, our results indicate that the expression of genes involved in translation and related processes are downregulated in the myocardium of 25≥ BMI ≤ 32 patients. In contrast genes involved in muscle contraction are overexpressed. To establish whether the two groups of genes are functionally related requires further research. Future research should also assess whether interventions targeting these processes may have translational potential for clinical myocardial protection.

## Supporting information

Adebayo et al - Supplemental Material

Adebayo et al - STROBE checklist

## Data Availability

Sequencing and samples group data are available via NCBI Gene Expression Omnibus (GSE159612)

https://www.ncbi.nlm.nih.gov/geo/query/acc.cgi?acc=GSE159612

## Funding Sources

Van Geest Foundation, Leicester NIHR Biomedical Research Centre, British Heart Foundation CH/12/1/29419, AA18/3/34220.

## Disclosures

Mrs Kumar, Prof. Murphy and Dr Woźniak received a grant from Zimmer Biomet. Dr Murphy also received grants from Terumo and Baxter. The remaining authors have disclosed that they do not have any potential conflicts of interest.

## Notes

### Clinical Trial

NCT02908009

### Author Declarations

The East Midlands Nottingham 1 Research Ethics Committee

