## Supplementary material for "Gene and metabolite expression dependence on body mass index in human myocardium": Adebayo et al - Supplemental Material

**Table S1** – Breakdown of samples with transcriptomics and metabolomics analyses.

| ID | Transcriptomics | Metabolomics | BMI | Group |
|---|---|---|---|---|
| 1 | Yes | Yes | 18 | BMI<25 |
| 2 | Yes | Yes | 19 | BMI<25 |
| 3 | Yes | Yes | 19 | BMI<25 |
| 4 | Yes | Yes | 21 | BMI<25 |
| 58 | RIN<8 | Yes | 21 | BMI<25 |
| 5 | Yes | Yes | 22 | BMI<25 |
| 6 | Yes | Yes | 22 | BMI<25 |
| 7 | Yes | Yes | 22 | BMI<25 |
| 34 | Yes | Yes | 22 | BMI<25 |
| 38 | Yes | Yes | 22 | BMI<25 |
| 42 | Yes | Yes | 22 | BMI<25 |
| 47 | Yes | Yes | 23 | BMI<25 |
| 48 | Yes | Yes | 23 | BMI<25 |
| 8 | Yes | Yes | 24 | BMI<25 |
| 9 | Yes | Yes | 24 | BMI<25 |
| 29 | Yes | Yes | 24 | BMI<25 |
| 10 | Yes | Yes | 25 | 25≥BMI≤32 |
| 11 | Yes | Yes | 25 | 25≥BMI≤32 |
| 12 | Yes | Yes | 25 | 25≥BMI≤32 |
| 32 | Yes | Yes | 25 | 25≥BMI≤32 |
| 13 | Yes | Yes | 26 | 25≥BMI≤32 |
| 14 | Yes | Yes | 26 | 25≥BMI≤32 |
| 28 | Yes | Yes | 26 | 25≥BMI≤32 |
| 30 | Yes | Yes | 26 | 25≥BMI≤32 |
| 33 | Yes | Yes | 26 | 25≥BMI≤32 |
| 54 | RIN<8 | Yes | 26 | 25≥BMI≤32 |
| 55 | RIN<8 | Yes | 26 | 25≥BMI≤32 |
| 56 | RIN<8 | Yes | 26 | 25≥BMI≤32 |
| 15 | Yes | Yes | 27 | 25≥BMI≤32 |
| 27 | Yes | Yes | 27 | 25≥BMI≤32 |
| 16 | Yes | Yes | 28 | 25≥BMI≤32 |
| 17 | Yes | Yes | 28 | 25≥BMI≤32 |
| 18 | Yes | Yes | 28 | 25≥BMI≤32 |
| 36 | Yes | Yes | 28 | 25≥BMI≤32 |
| 41 | Yes | Yes | 28 | 25≥BMI≤32 |
| 19 | Yes | Yes | 29 | 25≥BMI≤32 |
| 20 | Yes | Yes | 29 | 25≥BMI≤32 |
| 35 | Yes | Yes | 29 | 25≥BMI≤32 |
| 49 | Yes | Yes | 29 | 25≥BMI≤32 |
| 21 | Yes | Yes | 30 | 25≥BMI≤32 |
| 45 | Yes | Yes | 30 | 25≥BMI≤32 |
| 53 | Yes | NA | 31 | 25≥BMI≤32 |
| 22 | Yes | Yes | 32 | 25≥BMI≤32 |
| 43 | Yes | removed | 32 | 25≥BMI≤32 |
| 25 | Yes | Yes | 32 | 25≥BMI≤32 |
| 40 | Yes | Yes | 32 | 25≥BMI≤32 |
| 52 | Yes | NA | 32 | 25≥BMI≤32 |
| 26 | Yes | Yes | 34 | BMI>32 |
| 39 | Yes | Yes | 34 | BMI>32 |
| 44 | Yes | Yes | 34 | BMI>32 |
| 57 | RIN<8 | Yes | 34 | BMI>32 |
| 46 | Yes | Yes | 34 | BMI>32 |
| 50 | Yes | Yes | 34 | BMI>32 |
| 23 | Yes | Yes | 35 | BMI>32 |
| 31 | Yes | Yes | 35 | BMI>32 |
| 37 | Yes | Yes | 38 | BMI>32 |
| 51 | Yes | Yes | 39 | BMI>32 |
| 24 | Yes | Yes | 41 | BMI>32 |
| 59 | NA | NA | 29 | 25≥BMI≤32 |
| 60 | NA | NA | 27 | 25≥BMI≤32 |
| 61 | NA | NA | 33 | BMI>32 |
| 62 | NA | NA | 29 | 25≥BMI≤32 |
| 63 | NA | NA | 29 | 25≥BMI≤32 |
| 64 | NA | NA | 28 | 25≥BMI≤32 |
| 65 | NA | NA | 32 | 25≥BMI≤32 |
| 66 | NA | NA | 28 | 25≥BMI≤32 |

Table S2 – Results of the Two Lines analysis for all transcripts significant for both regression lines and for metabolites significant between the analyzed group. z1, z2 – test statistics for both regression lines; b1, b2 – regression coefficients, p1, p2 – p-values for each regression line.

| Ensembl ID | Gene description | Gene/metabolite name | Type | b1 | z1 | p1 | b2 | z2 | p2 | breakpoint |
|---|---|---|---|---|---|---|---|---|---|---|
| ENSG00000272768 | novel transcript, antisense to PURB | AC004854.2 | lncRNA | -0.446 | -3.137 | 0.002 | 0.090 | 2.637 | 0.008 | 24 |
| ENSG00000269696 | novel transcript, antisense to ZFP28 | AC005498.3 | lncRNA | -0.209 | -2.230 | 0.026 | 0.081 | 2.182 | 0.029 | 25 |
| ENSG00000224063 | novel transcript, antisense to TFPI and CALCRL | AC007319.1 | lncRNA | 0.163 | 2.556 | 0.011 | -0.113 | -2.839 | 0.005 | 28 |
| ENSG00000279249 | novel transcript, antisense to CBLN1 | AC007614.1 | lncRNA | 0.189 | 2.883 | 0.004 | -0.180 | -2.176 | 0.030 | 28 |
| ENSG00000267179 | novel protein | AC008770.2 | protein_coding | -0.184 | -2.105 | 0.035 | 0.257 | 3.266 | 0.001 | 29 |
| ENSG00000279598 | novel transcript, antisense to titin | AC009948.3 | TEC | 0.131 | 2.658 | 0.008 | -0.121 | -2.618 | 0.009 | 26 |
| ENSG00000272622 | novel transcript | AC010735.2 | lncRNA | -0.138 | -2.170 | 0.030 | 0.042 | 2.067 | 0.039 | 26 |
| ENSG00000285596 | novel transcript | AC017116.2 | lncRNA | 0.151 | 2.428 | 0.015 | -0.116 | -2.319 | 0.020 | 28 |
| ENSG00000186019 | novel transcript, antisense to ZNF225 and ZNF224 | AC021092.1 | lncRNA | 0.096 | 2.072 | 0.038 | -0.089 | -2.863 | 0.004 | 30 |
| ENSG00000248115 | novel transcript | AC023154.1 | lncRNA | -0.083 | -2.171 | 0.030 | 0.059 | 2.625 | 0.009 | 28 |
| ENSG00000273771 | novel transcript | AC024337.2 | lncRNA | -0.102 | -2.679 | 0.007 | 0.087 | 3.059 | 0.002 | 27 |
| ENSG00000273329 | novel transcript | AC078846.1 | lncRNA | 0.038 | 2.365 | 0.018 | -0.112 | -2.986 | 0.003 | 32 |
| ENSG00000253190 | novel transcript | AC084082.1 | lncRNA | -0.183 | -2.264 | 0.024 | 0.113 | 3.199 | 0.001 | 26 |
| ENSG00000267127 | novel protein | AC090360.1 | protein_coding | 0.100 | 2.769 | 0.006 | -0.186 | -3.917 | 0.000 | 32 |
| ENSG00000279283 | novel transcript | AC131009.4 | TEC | 0.115 | 2.272 | 0.023 | -0.096 | -2.424 | 0.015 | 28 |
| ENSG00000279204 | TEC | AC134043.3 | TEC | -0.167 | -2.674 | 0.008 | 0.065 | 2.122 | 0.034 | 26 |
| ENSG00000182827 | acyl-CoA binding domain containing 3 [Source:HGNC Symbol;Acc:HGNC:15453] | ACBD3 | protein_coding | -0.041 | -2.533 | 0.011 | 0.018 | 2.151 | 0.032 | 26 |
| ENSG00000118507 | A-kinase anchoring protein 7 [Source:HGNC Symbol;Acc:HGNC:377] | AKAP7 | protein_coding | -0.076 | -2.601 | 0.009 | 0.083 | 3.676 | 0.000 | 28 |
| ENSG00000285723 | novel protein | AL034430.2 | protein_coding | 0.070 | 1.997 | 0.046 | -0.073 | -2.264 | 0.024 | 29 |

| | | | | | | | | | | |
|---|---|---|---|---|---|---|---|---|---|---|
| ENSG00000276809 | novel transcript, sense intronic to DNAJC3 | AL138955.1 | lncRNA | -0.279 | -4.095 | 0.000 | 0.088 | 2.432 | 0.015 | 26 |
| ENSG00000187186 | uncharacterized LOC730098 [Source:NCBI gene (formerly Entrezgene);Acc:730098] | AL162231.1 | protein_coding | -0.108 | -1.983 | 0.047 | 0.078 | 1.972 | 0.049 | 26 |
| ENSG00000269896 | small nuclear ribonucleoprotein N (SNRPN) pseudogene | AL513477.1 | transcribed_processed_pseudogene | 0.095 | 2.243 | 0.025 | -0.154 | -2.569 | 0.010 | 28 |
| ENSG00000253710 | ALG11 alpha-1,2-mannosyltransferase [Source:HGNC Symbol;Acc:HGNC:32456] | ALG11 | protein_coding | 0.034 | 2.115 | 0.034 | -0.025 | -2.198 | 0.028 | 29 |
| ENSG00000139645 | ankyrin repeat domain 52 [Source:HGNC Symbol;Acc:HGNC:26614] | ANKRD52 | protein_coding | 0.047 | 2.300 | 0.021 | -0.045 | -2.323 | 0.020 | 28 |
| ENSG00000151572 | anoctamin 4 [Source:HGNC Symbol;Acc:HGNC:23837] | ANO4 | protein_coding | 0.100 | 2.381 | 0.017 | -0.198 | -3.228 | 0.001 | 29 |
| ENSG00000106367 | adaptor related protein complex 1 subunit sigma 1 [Source:HGNC Symbol;Acc:HGNC:559] | AP1S1 | protein_coding | -0.040 | -2.176 | 0.030 | 0.048 | 2.137 | 0.033 | 30 |
| ENSG00000136044 | adaptor protein, phosphotyrosine interacting with PH domain and leucine zipper 2 [Source:HGNC Symbol;Acc:HGNC:18242] | APPL2 | protein_coding | 0.053 | 2.519 | 0.012 | -0.040 | -2.633 | 0.008 | 28 |
| ENSG00000177479 | ariadne RBR E3 ubiquitin protein ligase 2 [Source:HGNC Symbol;Acc:HGNC:690] | ARIH2 | protein_coding | -0.072 | -2.156 | 0.031 | 0.011 | 1.983 | 0.047 | 24 |
| ENSG00000177917 | ADP ribosylation factor like GTPase 6 interacting protein 6 [Source:HGNC Symbol;Acc:HGNC:24048] | ARL6IP6 | protein_coding | -0.106 | -2.635 | 0.008 | 0.054 | 2.031 | 0.042 | 27 |
| ENSG00000224470 | ataxin 1 like [Source:HGNC Symbol;Acc:HGNC:33279] | ATXN1L | protein_coding | -0.024 | -3.174 | 0.002 | 0.021 | 2.745 | 0.006 | 28 |
| ENSG00000105778 | AVL9 cell migration associated [Source:HGNC Symbol;Acc:HGNC:28994] | AVL9 | protein_coding | 0.048 | 2.834 | 0.005 | -0.024 | -1.971 | 0.049 | 26 |
| ENSG00000162630 | beta-1,3-galactosyltransferase 2 [Source:HGNC Symbol;Acc:HGNC:917] | B3GALT2 | protein_coding | -0.110 | -2.674 | 0.007 | 0.038 | 2.138 | 0.032 | 25 |
| ENSG00000123810 | B9 domain containing 2 [Source:HGNC Symbol;Acc:HGNC:28636] | B9D2 | protein_coding | 0.143 | 2.186 | 0.029 | -0.082 | -2.050 | 0.040 | 28 |

| | | | | | | | | | | |
|---|---|---|---|---|---|---|---|---|---|---|
| ENSG00000169925 | bromodomain containing 3 [Source:HGNC Symbol;Acc:HGNC:1104] | BRD3 | protein_coding | 0.037 | 2.912 | 0.004 | -0.039 | -2.805 | 0.005 | 29 |
| ENSG00000164061 | bassoon presynaptic cytomatrix protein [Source:HGNC Symbol;Acc:HGNC:1117] | BSN | protein_coding | 0.219 | 3.092 | 0.002 | -0.066 | -2.848 | 0.004 | 25 |
| ENSG00000148925 | BTB domain containing 10 [Source:HGNC Symbol;Acc:HGNC:21445] | BTBD10 | protein_coding | -0.057 | -3.007 | 0.003 | 0.024 | 2.199 | 0.028 | 27 |
| ENSG00000240809 | CAP1 pseudogene 1 [Source:HGNC Symbol;Acc:HGNC:31134] | CAP1P1 | processed_pseudogene | -0.084 | -2.217 | 0.027 | 0.090 | 2.214 | 0.027 | 29 |
| ENSG00000110619 | cysteinyl-tRNA synthetase 1 [Source:HGNC Symbol;Acc:HGNC:1493] | CARS1 | protein_coding | 0.041 | 2.210 | 0.027 | -0.044 | -2.616 | 0.009 | 29 |
| ENSG00000165813 | coiled-coil domain containing 186 [Source:HGNC Symbol;Acc:HGNC:24349] | CCDC186 | protein_coding | -0.037 | -2.090 | 0.037 | 0.023 | 2.444 | 0.015 | 25 |
| ENSG00000166510 | coiled-coil domain containing 68 [Source:HGNC Symbol;Acc:HGNC:24350] | CCDC68 | protein_coding | 0.066 | 2.633 | 0.008 | -0.063 | -3.113 | 0.002 | 28 |
| ENSG00000275385 | C-C motif chemokine ligand 18 [Source:HGNC Symbol;Acc:HGNC:10616] | CCL18 | protein_coding | -0.186 | -2.677 | 0.007 | 0.414 | 3.641 | 0.000 | 32 |
| ENSG00000149654 | cadherin 22 [Source:HGNC Symbol;Acc:HGNC:13251] | CDH22 | protein_coding | 0.111 | 2.179 | 0.029 | -0.162 | -2.528 | 0.011 | 28 |
| ENSG00000213753 | CENPB DNA-binding domains containing 1 pseudogene 1 [Source:HGNC Symbol;Acc:HGNC:28421] | CENPBD1P1 | transcribed_processed_pseudogene | -0.069 | -2.235 | 0.025 | 0.078 | 3.088 | 0.002 | 27 |
| ENSG00000172831 | carboxylesterase 2 [Source:HGNC Symbol;Acc:HGNC:1864] | CES2 | protein_coding | 0.027 | 2.195 | 0.028 | -0.024 | -2.540 | 0.011 | 28 |
| ENSG00000116785 | complement factor H related 3 [Source:HGNC Symbol;Acc:HGNC:16980] | CFHR3 | protein_coding | -0.343 | -2.763 | 0.006 | 0.106 | 2.179 | 0.029 | 25 |
| ENSG00000125611 | coiled-coil-helix-coiled-coil-helix domain containing 5 [Source:HGNC Symbol;Acc:HGNC:17840] | CHCHD5 | protein_coding | -0.050 | -2.142 | 0.032 | 0.051 | 2.135 | 0.033 | 29 |
| ENSG00000090539 | chordin [Source:HGNC Symbol;Acc:HGNC:1949] | CHRD | protein_coding | 0.075 | 2.358 | 0.018 | -0.107 | -4.389 | 0.000 | 29 |
| ENSG00000247572 | CKMT2 antisense RNA 1 [Source:HGNC Symbol;Acc:HGNC:48997] | CKMT2-AS1 | lncRNA | 0.114 | 2.156 | 0.031 | -0.054 | -2.442 | 0.015 | 28 |

| | | | | | | | | | | |
|---|---|---|---|---|---|---|---|---|---|---|
| ENSG00000175505 | cardiotrophin like cytokine factor 1 [Source:HGNC Symbol;Acc:HGNC:17412] | CLCF1 | protein_coding | -0.070 | -2.651 | 0.008 | 0.114 | 2.587 | 0.010 | 32 |
| ENSG00000111729 | C-type lectin domain family 4 member A [Source:HGNC Symbol;Acc:HGNC:13257] | CLEC4A | protein_coding | -0.204 | -2.578 | 0.010 | 0.285 | 3.403 | 0.001 | 30 |
| ENSG00000198791 | CCR4-NOT transcription complex subunit 7 [Source:HGNC Symbol;Acc:HGNC:14101] | CNOT7 | protein_coding | -0.018 | -2.191 | 0.028 | 0.018 | 2.200 | 0.028 | 28 |
| ENSG00000136943 | cathepsin V [Source:HGNC Symbol;Acc:HGNC:2538] | CTSV | protein_coding | -0.098 | -1.962 | 0.050 | 0.248 | 2.919 | 0.004 | 29 |
| ENSG00000132716 | DDB1 and CUL4 associated factor 8 [Source:HGNC Symbol;Acc:HGNC:24891] | DCAF8 | protein_coding | 0.039 | 2.759 | 0.006 | -0.033 | -2.178 | 0.029 | 26 |
| ENSG00000145214 | diacylglycerol kinase theta [Source:HGNC Symbol;Acc:HGNC:2856] | DGKQ | protein_coding | 0.223 | 3.617 | 0.000 | -0.056 | -2.940 | 0.003 | 25 |
| ENSG00000138246 | DnaJ heat shock protein family (Hsp40) member C13 [Source:HGNC Symbol;Acc:HGNC:30343] | DNAJC13 | protein_coding | 0.028 | 3.625 | 0.000 | -0.038 | -2.974 | 0.003 | 29 |
| ENSG00000178498 | deltex E3 ubiquitin ligase 3 [Source:HGNC Symbol;Acc:HGNC:24457] | DTX3 | protein_coding | 0.018 | 2.124 | 0.034 | -0.040 | -2.110 | 0.035 | 32 |
| ENSG00000243701 | DPPA2 upstream binding RNA [Source:HGNC Symbol;Acc:HGNC:48569] | DUBR | lncRNA | 0.058 | 2.567 | 0.010 | -0.047 | -2.271 | 0.023 | 29 |
| ENSG00000093144 | ethylmalonyl-CoA decarboxylase 1 [Source:HGNC Symbol;Acc:HGNC:21489] | ECHDC1 | protein_coding | 0.064 | 3.604 | 0.000 | -0.022 | -2.117 | 0.034 | 26 |
| ENSG00000203734 | epithelial cell transforming 2 like [Source:HGNC Symbol;Acc:HGNC:21118] | ECT2L | protein_coding | -0.387 | -2.423 | 0.015 | 0.094 | 2.022 | 0.043 | 25 |
| ENSG00000186976 | EF-hand calcium binding domain 6 [Source:HGNC Symbol;Acc:HGNC:24204] | EFCAB6 | protein_coding | -0.151 | -2.813 | 0.005 | 0.051 | 2.123 | 0.034 | 26 |
| ENSG00000145242 | EPH receptor A5 [Source:HGNC Symbol;Acc:HGNC:3389] | EPHA5 | protein_coding | 0.109 | 2.648 | 0.008 | -0.132 | -2.722 | 0.006 | 29 |
| ENSG00000089248 | endoplasmic reticulum protein 29 [Source:HGNC Symbol;Acc:HGNC:13799] | ERP29 | protein_coding | 0.035 | 2.037 | 0.042 | -0.018 | -2.599 | 0.009 | 26 |
| ENSG00000157557 | ETS proto-oncogene 2, transcription factor | ETS2 | protein_coding | -0.037 | -2.199 | 0.028 | 0.045 | 2.913 | 0.004 | 28 |

| | | | | | | | | | | |
|---|---|---|---|---|---|---|---|---|---|---|
| | [Source:HGNC Symbol;Acc:HGNC:3489] | | | | | | | | | |
| ENSG00000184083 | family with sequence similarity 120C [Source:HGNC Symbol;Acc:HGNC:16949] | FAM120C | protein_coding | -0.073 | -2.155 | 0.031 | 0.043 | 2.208 | 0.027 | 28 |
| ENSG00000221909 | family with sequence similarity 200 member A [Source:HGNC Symbol;Acc:HGNC:25401] | FAM200A | protein_coding | -0.104 | -2.723 | 0.006 | 0.045 | 2.194 | 0.028 | 26 |
| ENSG00000071859 | family with sequence similarity 50 member A [Source:HGNC Symbol;Acc:HGNC:18786] | FAM50A | protein_coding | -0.066 | -5.312 | 0.000 | 0.022 | 2.051 | 0.040 | 26 |
| ENSG00000167196 | F-box protein 22 [Source:HGNC Symbol;Acc:HGNC:13593] | FBXO22 | protein_coding | -0.114 | -2.683 | 0.007 | 0.029 | 2.077 | 0.038 | 25 |
| ENSG00000142748 | ficolin 3 [Source:HGNC Symbol;Acc:HGNC:3625] | FCN3 | protein_coding | 0.182 | 3.313 | 0.001 | -0.067 | -2.052 | 0.040 | 28 |
| ENSG00000109158 | gamma-aminobutyric acid type A receptor subunit alpha4 [Source:HGNC Symbol;Acc:HGNC:4078] | GABRA4 | protein_coding | -0.391 | -3.287 | 0.001 | 0.143 | 2.749 | 0.006 | 26 |
| ENSG00000136542 | polypeptide N-acetylgalactosaminyltransferase 5 [Source:HGNC Symbol;Acc:HGNC:4127] | GALNT5 | protein_coding | 0.076 | 2.960 | 0.003 | -0.061 | -2.015 | 0.044 | 29 |
| ENSG00000005436 | GC-rich sequence DNA-binding factor 2 [Source:HGNC Symbol;Acc:HGNC:1317] | GCFC2 | protein_coding | -0.128 | -4.328 | 0.000 | 0.041 | 2.975 | 0.003 | 25 |
| ENSG00000162676 | growth factor independent 1 transcriptional repressor [Source:HGNC Symbol;Acc:HGNC:4237] | GFI1 | protein_coding | 0.176 | 2.324 | 0.020 | -0.163 | -2.387 | 0.017 | 29 |
| ENSG00000163655 | guanine monophosphate synthase [Source:HGNC Symbol;Acc:HGNC:4378] | GMPS | protein_coding | -0.027 | -2.266 | 0.023 | 0.035 | 2.255 | 0.024 | 29 |
| ENSG00000087258 | G protein subunit alpha o1 [Source:HGNC Symbol;Acc:HGNC:4389] | GNAO1 | protein_coding | 0.040 | 1.996 | 0.046 | -0.033 | -2.264 | 0.024 | 29 |
| ENSG00000108433 | golgi SNAP receptor complex member 2 [Source:HGNC Symbol;Acc:HGNC:4431] | GOSR2 | protein_coding | -0.038 | -2.022 | 0.043 | 0.050 | 3.793 | 0.000 | 29 |
| ENSG00000159592 | GC-rich promoter binding protein 1 like 1 [Source:HGNC Symbol;Acc:HGNC:28843] | GPBP1L1 | protein_coding | -0.037 | -2.382 | 0.017 | 0.013 | 2.529 | 0.011 | 25 |
| ENSG00000156097 | G protein-coupled receptor 61 [Source:HGNC Symbol;Acc:HGNC:13300] | GPR61 | protein_coding | 0.172 | 2.217 | 0.027 | -0.183 | -2.579 | 0.010 | 28 |

| | | | | | | | | | | |
|---|---|---|---|---|---|---|---|---|---|---|
| ENSG00000123901 | G protein-coupled receptor 83 [Source:HGNC Symbol;Acc:HGNC:4523] | GPR83 | protein_coding | 0.137 | 2.406 | 0.016 | -0.099 | -1.990 | 0.047 | 28 |
| ENSG00000125675 | glutamate ionotropic receptor AMPA type subunit 3 [Source:HGNC Symbol;Acc:HGNC:4573] | GRIA3 | protein_coding | 0.072 | 2.179 | 0.029 | -0.108 | -2.497 | 0.013 | 32 |
| ENSG00000228315 | GUSB pseudogene 11 [Source:HGNC Symbol;Acc:HGNC:42325] | GUSBP11 | lncRNA | 0.105 | 2.404 | 0.016 | -0.065 | -2.843 | 0.004 | 26 |
| ENSG00000092036 | HAUS augmin like complex subunit 4 [Source:HGNC Symbol;Acc:HGNC:20163] | HAUS4 | protein_coding | -0.041 | -2.124 | 0.034 | 0.038 | 2.480 | 0.013 | 29 |
| ENSG00000249115 | HAUS augmin like complex subunit 5 [Source:HGNC Symbol;Acc:HGNC:29130] | HAUS5 | protein_coding | 0.112 | 2.836 | 0.005 | -0.070 | -2.964 | 0.003 | 28 |
| ENSG00000162639 | HEN methyltransferase 1 [Source:HGNC Symbol;Acc:HGNC:26400] | HENMT1 | protein_coding | -0.083 | -2.742 | 0.006 | 0.104 | 2.909 | 0.004 | 29 |
| ENSG00000169660 | hexosaminidase D [Source:HGNC Symbol;Acc:HGNC:26307] | HEXD | protein_coding | 0.076 | 2.328 | 0.020 | -0.035 | -2.193 | 0.028 | 25 |
| ENSG00000164120 | 15-hydroxyprostaglandin dehydrogenase [Source:HGNC Symbol;Acc:HGNC:5154] | HPGD | protein_coding | -0.097 | -2.205 | 0.027 | 0.162 | 3.025 | 0.002 | 30 |
| ENSG00000178922 | hydroxypyruvate isomerase (putative) [Source:HGNC Symbol;Acc:HGNC:26948] | HYI | protein_coding | -0.058 | -2.057 | 0.040 | 0.057 | 3.382 | 0.001 | 28 |
| ENSG00000244242 | interferon induced transmembrane protein 10 [Source:HGNC Symbol;Acc:HGNC:40022] | IFITM10 | protein_coding | 0.140 | 2.381 | 0.017 | -0.162 | -2.105 | 0.035 | 28 |
| ENSG00000128581 | intraflagellar transport 22 [Source:HGNC Symbol;Acc:HGNC:21895] | IFT22 | protein_coding | -0.039 | -2.937 | 0.003 | 0.030 | 2.505 | 0.012 | 29 |
| ENSG00000211970 | immunoglobulin heavy variable 4-61 [Source:HGNC Symbol;Acc:HGNC:5655] | IGHV4-61 | IG_V_gene | -0.148 | -2.152 | 0.031 | 0.206 | 2.787 | 0.005 | 30 |
| ENSG00000244116 | immunoglobulin kappa variable 2-28 [Source:HGNC Symbol;Acc:HGNC:5783] | IGKV2-28 | IG_V_gene | -0.228 | -2.054 | 0.040 | 0.398 | 2.484 | 0.013 | 32 |
| ENSG00000134470 | interleukin 15 receptor subunit alpha [Source:HGNC Symbol;Acc:HGNC:5978] | IL15RA | protein_coding | 0.072 | 2.361 | 0.018 | -0.040 | -2.321 | 0.020 | 25 |

| | | | | | | | | | | |
|---|---|---|---|---|---|---|---|---|---|---|
| ENSG00000081148 | interphotoreceptor matrix proteoglycan 2 [Source:HGNC Symbol;Acc:HGNC:18362] | IMPG2 | protein_coding | -0.093 | -2.069 | 0.039 | 0.060 | 2.908 | 0.004 | 26 |
| ENSG00000151689 | inositol polyphosphate-1-phosphatase [Source:HGNC Symbol;Acc:HGNC:6071] | INPP1 | protein_coding | -0.037 | -2.037 | 0.042 | 0.029 | 2.260 | 0.024 | 29 |
| ENSG00000169896 | integrin subunit alpha M [Source:HGNC Symbol;Acc:HGNC:6149] | ITGAM | protein_coding | 0.149 | 3.388 | 0.001 | -0.057 | -3.093 | 0.002 | 26 |
| ENSG00000123700 | potassium inwardly rectifying channel subfamily J member 2 [Source:HGNC Symbol;Acc:HGNC:6263] | KCNJ2 | protein_coding | -0.142 | -2.409 | 0.016 | 0.060 | 2.625 | 0.009 | 25 |
| ENSG00000267365 | KCNJ2 antisense RNA 1 [Source:HGNC Symbol;Acc:HGNC:43720] | KCNJ2-AS1 | lncRNA | -0.117 | -2.057 | 0.040 | 0.073 | 3.476 | 0.001 | 27 |
| ENSG00000155666 | lysine demethylase 8 [Source:HGNC Symbol;Acc:HGNC:25840] | KDM8 | protein_coding | -0.085 | -2.103 | 0.035 | 0.078 | 2.934 | 0.003 | 28 |
| ENSG00000110427 | KIAA1549 like [Source:HGNC Symbol;Acc:HGNC:24836] | KIAA1549L | protein_coding | 0.272 | 3.271 | 0.001 | -0.132 | -2.755 | 0.006 | 26 |
| ENSG00000165185 | KIAA1958 [Source:HGNC Symbol;Acc:HGNC:23427] | KIAA1958 | protein_coding | -0.090 | -2.829 | 0.005 | 0.084 | 3.857 | 0.000 | 28 |
| ENSG00000174010 | kelch like family member 15 [Source:HGNC Symbol;Acc:HGNC:29347] | KLHL15 | protein_coding | -0.099 | -3.813 | 0.000 | 0.057 | 2.844 | 0.004 | 26 |
| ENSG00000114796 | kelch like family member 24 [Source:HGNC Symbol;Acc:HGNC:25947] | KLHL24 | protein_coding | -0.037 | -2.774 | 0.006 | 0.024 | 2.077 | 0.038 | 27 |
| ENSG00000230658 | KLHL7 divergent transcript [Source:HGNC Symbol;Acc:HGNC:43431] | KLHL7-DT | lncRNA | -0.279 | -3.088 | 0.002 | 0.078 | 2.141 | 0.032 | 26 |
| ENSG00000170484 | keratin 74 [Source:HGNC Symbol;Acc:HGNC:28929] | KRT74 | protein_coding | -0.150 | -2.101 | 0.036 | 0.573 | 2.787 | 0.005 | 32 |
| ENSG00000137944 | kynurenine aminotransferase 3 [Source:HGNC Symbol;Acc:HGNC:33238] | KYAT3 | protein_coding | -0.050 | -2.300 | 0.021 | 0.052 | 2.433 | 0.015 | 30 |
| ENSG00000188186 | late endosomal/lysosomal adaptor, MAPK and MTOR activator 4 [Source:HGNC Symbol;Acc:HGNC:33772] | LAMTOR4 | protein_coding | -0.022 | -2.436 | 0.015 | 0.053 | 4.332 | 0.000 | 32 |
| ENSG00000236090 | lactate dehydrogenase A pseudogene 3 [Source:HGNC Symbol;Acc:HGNC:6538] | LDHAP3 | processed_pseudogene | -0.175 | -2.095 | 0.036 | 0.096 | 2.166 | 0.030 | 26 |

| | | | | | | | | | | |
|---|---|---|---|---|---|---|---|---|---|---|
| ENSG00000223935 | LGALSL divergent transcript [Source:HGNC Symbol;Acc:HGNC:53951] | LGALSL-DT | lncRNA | 0.105 | 2.596 | 0.009 | -0.409 | -3.414 | 0.001 | 32 |
| ENSG00000050405 | LIM domain and actin binding 1 [Source:HGNC Symbol;Acc:HGNC:24636] | LIMA1 | protein_coding | 0.080 | 2.246 | 0.025 | -0.016 | -2.159 | 0.031 | 24 |
| ENSG00000148943 | lin-7 homolog C, crumbs cell polarity complex component [Source:HGNC Symbol;Acc:HGNC:17789] | LIN7C | protein_coding | -0.029 | -2.115 | 0.034 | 0.026 | 2.026 | 0.043 | 28 |
| ENSG00000223685 | long intergenic non-protein coding RNA 571 [Source:HGNC Symbol;Acc:HGNC:43721] | LINC00571 | lncRNA | -0.137 | -2.796 | 0.005 | 0.107 | 3.486 | 0.000 | 28 |
| ENSG00000223546 | long intergenic non-protein coding RNA 630 [Source:HGNC Symbol;Acc:HGNC:44263] | LINC00630 | lncRNA | 0.248 | 5.960 | 0.000 | -0.021 | -2.093 | 0.036 | 24 |
| ENSG00000281852 | long intergenic non-protein coding RNA 891 [Source:HGNC Symbol;Acc:HGNC:48577] | LINC00891 | lncRNA | -0.087 | -2.084 | 0.037 | 0.030 | 1.999 | 0.046 | 25 |
| ENSG00000235314 | long intergenic non-protein coding RNA 957 [Source:HGNC Symbol;Acc:HGNC:22332] | LINC00957 | lncRNA | -0.083 | -2.562 | 0.010 | 0.021 | 2.039 | 0.041 | 24 |
| ENSG00000261617 | long intergenic non-protein coding RNA 2177 [Source:HGNC Symbol;Acc:HGNC:53039] | LINC02177 | lncRNA | -0.194 | -1.966 | 0.049 | 0.102 | 2.123 | 0.034 | 26 |
| ENSG00000235160 | long intergenic non-protein coding RNA 2248 [Source:HGNC Symbol;Acc:HGNC:53147] | LINC02248 | lncRNA | -0.073 | -2.045 | 0.041 | 0.027 | 2.296 | 0.022 | 27 |
| ENSG00000251432 | long intergenic non-protein coding RNA 2615 [Source:HGNC Symbol;Acc:HGNC:53402] | LINC02615 | lncRNA | 0.106 | 2.214 | 0.027 | -0.134 | -2.049 | 0.040 | 30 |
| ENSG00000198121 | lysophosphatidic acid receptor 1 [Source:HGNC Symbol;Acc:HGNC:3166] | LPAR1 | protein_coding | -0.035 | -2.648 | 0.008 | 0.059 | 5.651 | 0.000 | 32 |
| ENSG00000148356 | leucine rich repeat and sterile alpha motif containing 1 [Source:HGNC Symbol;Acc:HGNC:25135] | LRSAM1 | protein_coding | 0.036 | 2.159 | 0.031 | -0.032 | -2.162 | 0.031 | 29 |
| ENSG00000180660 | mab-21 like 1 [Source:HGNC Symbol;Acc:HGNC:6757] | MAB21L1 | protein_coding | 0.062 | 2.085 | 0.037 | -0.221 | -2.657 | 0.008 | 32 |
| ENSG00000139625 | mitogen-activated protein kinase kinase kinase 12 | MAP3K12 | protein_coding | 0.055 | 2.577 | 0.010 | -0.073 | -3.675 | 0.000 | 29 |

| | | | | | | | | | | |
|---|---|---|---|---|---|---|---|---|---|---|
| | [Source:HGNC Symbol;Acc:HGNC:6851] | | | | | | | | | |
| ENSG00000156875 | major facilitator superfamily domain containing 14A [Source:HGNC Symbol;Acc:HGNC:23363] | MFSD14A | protein_coding | -0.034 | -2.177 | 0.029 | 0.022 | 2.253 | 0.024 | 28 |
| ENSG00000154889 | metallophosphoesterase 1 [Source:HGNC Symbol;Acc:HGNC:15988] | MPPE1 | protein_coding | 0.046 | 2.046 | 0.041 | -0.025 | -2.504 | 0.012 | 28 |
| ENSG00000198727 | mitochondrially encoded cytochrome b [Source:HGNC Symbol;Acc:HGNC:7427] | MT-CYB | protein_coding | 0.056 | 2.485 | 0.013 | -0.030 | -2.170 | 0.030 | 26 |
| ENSG00000132938 | microtubule associated scaffold protein 2 [Source:HGNC Symbol;Acc:HGNC:20595] | MTUS2 | protein_coding | 0.034 | 2.135 | 0.033 | -0.025 | -1.961 | 0.050 | 29 |
| ENSG00000205277 | mucin 12, cell surface associated [Source:HGNC Symbol;Acc:HGNC:7510] | MUC12 | protein_coding | -0.240 | -2.255 | 0.024 | 0.388 | 3.648 | 0.000 | 30 |
| ENSG00000013364 | major vault protein [Source:HGNC Symbol;Acc:HGNC:7531] | MVP | protein_coding | 0.042 | 2.057 | 0.040 | -0.029 | -1.962 | 0.050 | 27 |
| ENSG00000151503 | non-SMC condensin II complex subunit D3 [Source:HGNC Symbol;Acc:HGNC:28952] | NCAPD3 | protein_coding | -0.080 | -2.090 | 0.037 | 0.041 | 2.749 | 0.006 | 28 |
| ENSG00000116701 | neutrophil cytosolic factor 2 [Source:HGNC Symbol;Acc:HGNC:7661] | NCF2 | protein_coding | -0.069 | -2.871 | 0.004 | 0.109 | 2.438 | 0.015 | 31 |
| ENSG00000145912 | NHP2 ribonucleoprotein [Source:HGNC Symbol;Acc:HGNC:14377] | NHP2 | protein_coding | -0.047 | -3.112 | 0.002 | 0.030 | 2.838 | 0.005 | 28 |
| ENSG00000048162 | NOP16 nucleolar protein [Source:HGNC Symbol;Acc:HGNC:26934] | NOP16 | protein_coding | 0.061 | 3.355 | 0.001 | -0.037 | -2.866 | 0.004 | 29 |
| ENSG00000185551 | nuclear receptor subfamily 2 group F member 2 [Source:HGNC Symbol;Acc:HGNC:7976] | NR2F2 | protein_coding | 0.018 | 2.178 | 0.029 | -0.038 | -2.568 | 0.010 | 32 |
| ENSG00000179915 | neurexin 1 [Source:HGNC Symbol;Acc:HGNC:8008] | NRXN1 | protein_coding | -0.075 | -2.125 | 0.034 | 0.039 | 2.334 | 0.020 | 25 |
| ENSG00000173598 | nudix hydrolase 4 [Source:HGNC Symbol;Acc:HGNC:8051] | NUDT4 | protein_coding | -0.057 | -2.542 | 0.011 | 0.021 | 2.317 | 0.021 | 25 |
| ENSG00000111331 | 2'-5'-oligoadenylate synthetase 3 [Source:HGNC Symbol;Acc:HGNC:8088] | OAS3 | protein_coding | 0.062 | 2.164 | 0.030 | -0.074 | -2.216 | 0.027 | 28 |

| | | | | | | | | | | |
|---|---|---|---|---|---|---|---|---|---|---|
| ENSG00000116774 | olfactomedin like 3 [Source:HGNC Symbol;Acc:HGNC:24956] | OLFML3 | protein_coding | -0.040 | -2.043 | 0.041 | 0.027 | 2.016 | 0.044 | 29 |
| ENSG00000083093 | partner and localizer of BRCA2 [Source:HGNC Symbol;Acc:HGNC:26144] | PALB2 | protein_coding | -0.061 | -2.462 | 0.014 | 0.023 | 2.090 | 0.037 | 26 |
| ENSG00000100105 | POZ/BTB and AT hook containing zinc finger 1 [Source:HGNC Symbol;Acc:HGNC:13071] | PATZ1 | protein_coding | 0.020 | 2.096 | 0.036 | -0.039 | -2.541 | 0.011 | 32 |
| ENSG00000204304 | PBX homeobox 2 [Source:HGNC Symbol;Acc:HGNC:8633] | PBX2 | protein_coding | 0.021 | 2.182 | 0.029 | -0.020 | -2.286 | 0.022 | 32 |
| ENSG00000224729 | PCOLCE antisense RNA 1 [Source:HGNC Symbol;Acc:HGNC:40430] | PCOLCE-AS1 | lncRNA | 0.135 | 2.145 | 0.032 | -0.106 | -2.189 | 0.029 | 29 |
| ENSG00000115257 | proprotein convertase subtilisin/kexin type 4 [Source:HGNC Symbol;Acc:HGNC:8746] | PCSK4 | protein_coding | 0.209 | 2.298 | 0.022 | -0.081 | -2.282 | 0.022 | 25 |
| ENSG00000184588 | phosphodiesterase 4B [Source:HGNC Symbol;Acc:HGNC:8781] | PDE4B | protein_coding | -0.040 | -2.055 | 0.040 | 0.093 | 3.034 | 0.002 | 30 |
| ENSG00000229828 | PDE4DIP pseudogene 1 [Source:HGNC Symbol;Acc:HGNC:50867] | PDE4DIPP1 | unprocessed_pseudogene | 0.247 | 2.339 | 0.019 | -0.305 | -2.827 | 0.005 | 28 |
| ENSG00000088356 | p53 and DNA damage regulated 1 [Source:HGNC Symbol;Acc:HGNC:16119] | PDRG1 | protein_coding | 0.032 | 2.167 | 0.030 | -0.082 | -2.017 | 0.044 | 32 |
| ENSG00000172367 | PDZ domain containing 3 [Source:HGNC Symbol;Acc:HGNC:19891] | PDZD3 | protein_coding | 0.382 | 2.184 | 0.029 | -0.198 | -2.034 | 0.042 | 26 |
| ENSG00000142655 | peroxisomal biogenesis factor 14 [Source:HGNC Symbol;Acc:HGNC:8856] | PEX14 | protein_coding | 0.056 | 1.970 | 0.049 | -0.022 | -2.763 | 0.006 | 26 |
| ENSG00000204220 | prefoldin subunit 6 [Source:HGNC Symbol;Acc:HGNC:4926] | PFDN6 | protein_coding | -0.074 | -2.811 | 0.005 | 0.050 | 2.726 | 0.006 | 27 |
| ENSG00000103066 | phospholipase A2 group XV [Source:HGNC Symbol;Acc:HGNC:17163] | PLA2G15 | protein_coding | 0.033 | 2.294 | 0.022 | -0.046 | -2.650 | 0.008 | 27 |
| ENSG00000189266 | proline rich nuclear receptor coactivator 2 [Source:HGNC Symbol;Acc:HGNC:23158] | PNRC2 | protein_coding | -0.020 | -2.365 | 0.018 | 0.042 | 3.369 | 0.001 | 30 |

| | | | | | | | | | | |
|---|---|---|---|---|---|---|---|---|---|---|
| ENSG00000105568 | protein phosphatase 2 scaffold subunit Aalpha [Source:HGNC Symbol;Acc:HGNC:9302] | PPP2R1A | protein_coding | -0.034 | -2.310 | 0.021 | 0.018 | 2.034 | 0.042 | 26 |
| ENSG00000138738 | PR/SET domain 5 [Source:HGNC Symbol;Acc:HGNC:9349] | PRDM5 | protein_coding | -0.084 | -2.881 | 0.004 | 0.031 | 2.072 | 0.038 | 27 |
| ENSG00000204540 | psoriasis susceptibility 1 candidate 1 [Source:HGNC Symbol;Acc:HGNC:17202] | PSORS1C1 | protein_coding | 0.183 | 2.085 | 0.037 | -0.075 | -2.113 | 0.035 | 28 |
| ENSG00000134222 | proline and serine rich coiled-coil 1 [Source:HGNC Symbol;Acc:HGNC:24472] | PSRC1 | protein_coding | 0.069 | 2.553 | 0.011 | -0.118 | -4.530 | 0.000 | 31 |
| ENSG00000185920 | patched 1 [Source:HGNC Symbol;Acc:HGNC:9585] | PTCH1 | protein_coding | 0.058 | 2.134 | 0.033 | -0.102 | -2.557 | 0.011 | 31 |
| ENSG00000237984 | phosphatase and tensin homolog pseudogene 1 [Source:HGNC Symbol;Acc:HGNC:9589] | PTENP1 | transcribed_processed_pseudogene | -0.106 | -2.322 | 0.020 | 0.114 | 3.504 | 0.000 | 28 |
| ENSG00000099246 | RAB18, member RAS oncogene family [Source:HGNC Symbol;Acc:HGNC:14244] | RAB18 | protein_coding | -0.034 | -2.695 | 0.007 | 0.014 | 2.310 | 0.021 | 26 |
| ENSG00000172007 | RAB33B, member RAS oncogene family [Source:HGNC Symbol;Acc:HGNC:16075] | RAB33B | protein_coding | -0.055 | -2.425 | 0.015 | 0.034 | 5.120 | 0.000 | 26 |
| ENSG00000136144 | RCC1 and BTB domain containing protein 1 [Source:HGNC Symbol;Acc:HGNC:18243] | RCBTB1 | protein_coding | 0.026 | 2.043 | 0.041 | -0.024 | -2.551 | 0.011 | 29 |
| ENSG00000159788 | regulator of G protein signaling 12 [Source:HGNC Symbol;Acc:HGNC:9994] | RGS12 | protein_coding | 0.086 | 3.538 | 0.000 | -0.049 | -2.477 | 0.013 | 28 |
| ENSG00000091844 | regulator of G protein signaling 17 [Source:HGNC Symbol;Acc:HGNC:14088] | RGS17 | protein_coding | -0.122 | -2.325 | 0.020 | 0.142 | 2.405 | 0.016 | 29 |
| ENSG00000229927 | RHEB pseudogene 1 [Source:HGNC Symbol;Acc:HGNC:10010] | RHEBP1 | processed_pseudogene | 0.244 | 2.712 | 0.007 | -0.284 | -3.427 | 0.001 | 28 |
| ENSG00000140983 | ras homolog family member T2 [Source:HGNC Symbol;Acc:HGNC:21169] | RHOT2 | protein_coding | 0.046 | 2.651 | 0.008 | -0.071 | -2.912 | 0.004 | 29 |
| ENSG00000255794 | rhabdomyosarcoma 2 associated transcript [Source:HGNC Symbol;Acc:HGNC:29893] | RMST | lncRNA | -0.107 | -2.226 | 0.026 | 0.055 | 2.594 | 0.009 | 28 |

| | | | | | | | | | | |
|---|---|---|---|---|---|---|---|---|---|---|
| ENSG00000265727 | RNA, 7SL, cytoplasmic 648, pseudogene [Source:HGNC Symbol;Acc:HGNC:46664] | RN7SL648P | misc_RNA | 0.123 | 1.979 | 0.048 | -0.082 | -2.357 | 0.018 | 26 |
| ENSG00000133874 | ring finger protein 122 [Source:HGNC Symbol;Acc:HGNC:21147] | RNF122 | protein_coding | -0.038 | -1.973 | 0.049 | 0.067 | 1.990 | 0.047 | 28 |
| ENSG00000154133 | roundabout guidance receptor 4 [Source:HGNC Symbol;Acc:HGNC:17985] | ROBO4 | protein_coding | 0.061 | 2.200 | 0.028 | -0.061 | -2.235 | 0.025 | 28 |
| ENSG00000100316 | ribosomal protein L3 [Source:HGNC Symbol;Acc:HGNC:10332] | RPL3 | protein_coding | -0.025 | -2.644 | 0.008 | 0.025 | 2.764 | 0.006 | 29 |
| ENSG00000174444 | ribosomal protein L4 [Source:HGNC Symbol;Acc:HGNC:10353] | RPL4 | protein_coding | -0.028 | -2.526 | 0.012 | 0.019 | 2.127 | 0.033 | 29 |
| ENSG00000089009 | ribosomal protein L6 [Source:HGNC Symbol;Acc:HGNC:10362] | RPL6 | protein_coding | -0.016 | -2.321 | 0.020 | 0.016 | 2.021 | 0.043 | 29 |
| ENSG00000160208 | ribosomal RNA processing 1B [Source:HGNC Symbol;Acc:HGNC:23818] | RRP1B | protein_coding | -0.048 | -2.913 | 0.004 | 0.017 | 2.667 | 0.008 | 26 |
| ENSG00000134321 | radical S-adenosyl methionine domain containing 2 [Source:HGNC Symbol;Acc:HGNC:30908] | RSAD2 | protein_coding | 0.103 | 2.971 | 0.003 | -0.079 | -2.171 | 0.030 | 28 |
| ENSG00000167524 | ribosomal protein S6 kinase related [Source:HGNC Symbol;Acc:HGNC:26314] | RSKR | protein_coding | 0.085 | 2.049 | 0.041 | -0.052 | -2.328 | 0.020 | 29 |
| ENSG00000182552 | RWD domain containing 4 [Source:HGNC Symbol;Acc:HGNC:23750] | RWDD4 | protein_coding | 0.035 | 2.823 | 0.005 | -0.024 | -2.384 | 0.017 | 28 |
| ENSG00000163785 | receptor like tyrosine kinase [Source:HGNC Symbol;Acc:HGNC:10481] | RYK | protein_coding | -0.072 | -2.830 | 0.005 | 0.018 | 2.079 | 0.038 | 25 |
| ENSG00000246273 | SBF2 antisense RNA 1 [Source:HGNC Symbol;Acc:HGNC:27438] | SBF2-AS1 | lncRNA | -0.096 | -2.650 | 0.008 | 0.111 | 2.543 | 0.011 | 28 |
| ENSG00000166562 | SEC11 homolog C, signal peptidase complex subunit [Source:HGNC Symbol;Acc:HGNC:23400] | SEC11C | protein_coding | -0.054 | -3.153 | 0.002 | 0.025 | 2.145 | 0.032 | 27 |
| ENSG00000162430 | selenoprotein N [Source:HGNC Symbol;Acc:HGNC:15999] | SELENON | protein_coding | 0.047 | 2.210 | 0.027 | -0.020 | -2.512 | 0.012 | 25 |

| | | | | | | | | | | |
|---|---|---|---|---|---|---|---|---|---|---|
| ENSG00000197417 | sedoheptulokinase [Source:HGNC Symbol;Acc:HGNC:1492] | SHPK | protein_coding | 0.084 | 2.976 | 0.003 | -0.030 | -2.312 | 0.021 | 25 |
| ENSG00000204351 | Ski2 like RNA helicase [Source:HGNC Symbol;Acc:HGNC:10898] | SKIV2L | protein_coding | 0.042 | 2.619 | 0.009 | -0.061 | -2.458 | 0.014 | 30 |
| ENSG00000117090 | signaling lymphocytic activation molecule family member 1 [Source:HGNC Symbol;Acc:HGNC:10903] | SLAMF1 | protein_coding | 0.346 | 3.717 | 0.000 | -0.135 | -2.122 | 0.034 | 26 |
| ENSG00000151012 | solute carrier family 7 member 11 [Source:HGNC Symbol;Acc:HGNC:11059] | SLC7A11 | protein_coding | -0.139 | -2.700 | 0.007 | 0.109 | 2.683 | 0.007 | 28 |
| ENSG00000137571 | solute carrier organic anion transporter family member 5A1 [Source:HGNC Symbol;Acc:HGNC:19046] | SLCO5A1 | protein_coding | 0.040 | 2.048 | 0.041 | -0.053 | -2.367 | 0.018 | 29 |
| ENSG00000213599 | SLX1A-SULT1A3 readthrough (NMD candidate) [Source:HGNC Symbol;Acc:HGNC:44437] | SLX1A-SULT1A3 | lncRNA | 0.248 | 2.067 | 0.039 | -0.210 | -1.960 | 0.050 | 26 |
| ENSG00000170365 | SMAD family member 1 [Source:HGNC Symbol;Acc:HGNC:6767] | SMAD1 | protein_coding | 0.088 | 2.263 | 0.024 | -0.071 | -2.621 | 0.009 | 28 |
| ENSG00000198952 | SMG5 nonsense mediated mRNA decay factor [Source:HGNC Symbol;Acc:HGNC:24644] | SMG5 | protein_coding | 0.021 | 2.334 | 0.020 | -0.044 | -4.206 | 0.000 | 32 |
| ENSG00000172594 | sphingomyelin phosphodiesterase acid like 3A [Source:HGNC Symbol;Acc:HGNC:17389] | SMPDL3A | protein_coding | -0.034 | -2.577 | 0.010 | 0.048 | 2.574 | 0.010 | 32 |
| ENSG00000102172 | spermine synthase [Source:HGNC Symbol;Acc:HGNC:11123] | SMS | protein_coding | -0.064 | -3.330 | 0.001 | 0.019 | 2.091 | 0.037 | 26 |
| ENSG00000100028 | small nuclear ribonucleoprotein D3 polypeptide [Source:HGNC Symbol;Acc:HGNC:11160] | SNRPD3 | protein_coding | -0.043 | -2.314 | 0.021 | 0.030 | 2.169 | 0.030 | 27 |
| ENSG00000110025 | sorting nexin 15 [Source:HGNC Symbol;Acc:HGNC:14978] | SNX15 | protein_coding | -0.045 | -2.048 | 0.041 | 0.056 | 2.514 | 0.012 | 29 |
| ENSG00000109610 | superoxide dismutase 3 [Source:HGNC Symbol;Acc:HGNC:11181] | SOD3 | protein_coding | 0.045 | 2.078 | 0.038 | -0.042 | -2.947 | 0.003 | 28 |

| | | | | | | | | | | |
|---|---|---|---|---|---|---|---|---|---|---|
| ENSG00000067066 | SP100 nuclear antigen [Source:HGNC Symbol;Acc:HGNC:11206] | SP100 | protein_coding | 0.031 | 2.612 | 0.009 | -0.016 | -2.406 | 0.016 | 28 |
| ENSG00000122432 | spermatogenesis associated 1 [Source:HGNC Symbol;Acc:HGNC:14682] | SPATA1 | protein_coding | -0.177 | -3.427 | 0.001 | 0.132 | 2.790 | 0.005 | 28 |
| ENSG00000189419 | spermatogenesis associated 41 [Source:HGNC Symbol;Acc:HGNC:48613] | SPATA41 | lncRNA | 0.130 | 2.321 | 0.020 | -0.216 | -2.248 | 0.025 | 28 |
| ENSG00000116096 | sepiapterin reductase [Source:HGNC Symbol;Acc:HGNC:11257] | SPR | protein_coding | 0.033 | 1.979 | 0.048 | -0.030 | -2.199 | 0.028 | 28 |
| ENSG00000277363 | SRC kinase signaling inhibitor 1 [Source:HGNC Symbol;Acc:HGNC:29506] | SRCIN1 | protein_coding | -0.115 | -2.177 | 0.029 | 0.123 | 3.402 | 0.001 | 28 |
| ENSG00000008513 | ST3 beta-galactoside alpha-2,3-sialyltransferase 1 [Source:HGNC Symbol;Acc:HGNC:10862] | ST3GAL1 | protein_coding | 0.018 | 2.240 | 0.025 | -0.068 | -5.242 | 0.000 | 32 |
| ENSG00000144057 | ST6 beta-galactoside alpha-2,6-sialyltransferase 2 [Source:HGNC Symbol;Acc:HGNC:10861] | ST6GAL2 | protein_coding | 0.183 | 2.772 | 0.006 | -0.213 | -2.549 | 0.011 | 29 |
| ENSG00000160408 | ST6 N-acetylgalactosaminide alpha-2,6-sialyltransferase 6 [Source:HGNC Symbol;Acc:HGNC:23364] | ST6GALNAC6 | protein_coding | -0.046 | -3.403 | 0.001 | 0.020 | 2.314 | 0.021 | 28 |
| ENSG00000213533 | STIM activating enhancer [Source:HGNC Symbol;Acc:HGNC:30526] | STIMATE | protein_coding | 0.156 | 3.778 | 0.000 | -0.048 | -2.353 | 0.019 | 26 |
| ENSG00000162520 | syncoilin, intermediate filament protein [Source:HGNC Symbol;Acc:HGNC:28897] | SYNC | protein_coding | 0.058 | 2.365 | 0.018 | -0.039 | -2.381 | 0.017 | 29 |
| ENSG00000177156 | transaldolase 1 [Source:HGNC Symbol;Acc:HGNC:11559] | TALDO1 | protein_coding | -0.023 | -2.168 | 0.030 | 0.042 | 3.088 | 0.002 | 29 |
| ENSG00000138336 | tet methylcytosine dioxygenase 1 [Source:HGNC Symbol;Acc:HGNC:29484] | TET1 | protein_coding | -0.031 | -2.086 | 0.037 | 0.025 | 2.089 | 0.037 | 28 |
| ENSG00000174796 | THAP domain containing 6 [Source:HGNC Symbol;Acc:HGNC:23189] | THAP6 | protein_coding | 0.043 | 2.154 | 0.031 | -0.021 | -1.993 | 0.046 | 28 |
| ENSG00000144229 | thrombospondin type 1 domain containing 7B [Source:HGNC Symbol;Acc:HGNC:29348] | THSD7B | protein_coding | 0.099 | 2.389 | 0.017 | -0.142 | -2.700 | 0.007 | 32 |

| | | | | | | | | | | |
|---|---|---|---|---|---|---|---|---|---|---|
| ENSG00000066654 | THUMP domain containing 1 [Source:HGNC Symbol;Acc:HGNC:23807] | THUMPD1 | protein_coding | -0.044 | -2.314 | 0.021 | 0.032 | 2.156 | 0.031 | 28 |
| ENSG00000163659 | TCDD inducible poly(ADP-ribose) polymerase [Source:HGNC Symbol;Acc:HGNC:23696] | TIPARP | protein_coding | -0.027 | -2.789 | 0.005 | 0.097 | 2.822 | 0.005 | 32 |
| ENSG00000228828 | tousled like kinase 2 pseudogene 2 [Source:HGNC Symbol;Acc:HGNC:22227] | TLK2P2 | processed_pseudogene | 0.108 | 2.520 | 0.012 | -0.209 | -3.340 | 0.001 | 32 |
| ENSG00000164124 | transmembrane protein 144 [Source:HGNC Symbol;Acc:HGNC:25633] | TMEM144 | protein_coding | -0.042 | -2.752 | 0.006 | 0.032 | 2.808 | 0.005 | 27 |
| ENSG00000163444 | transmembrane protein 183A [Source:HGNC Symbol;Acc:HGNC:20173] | TMEM183A | protein_coding | 0.066 | 2.038 | 0.042 | -0.020 | -2.656 | 0.008 | 26 |
| ENSG00000187049 | transmembrane protein 216 [Source:HGNC Symbol;Acc:HGNC:25018] | TMEM216 | protein_coding | -0.063 | -2.281 | 0.023 | 0.063 | 2.364 | 0.018 | 31 |
| ENSG00000205084 | transmembrane protein 231 [Source:HGNC Symbol;Acc:HGNC:37234] | TMEM231 | protein_coding | -0.074 | -2.893 | 0.004 | 0.062 | 3.333 | 0.001 | 29 |
| ENSG00000134490 | transmembrane protein 241 [Source:HGNC Symbol;Acc:HGNC:31723] | TMEM241 | protein_coding | 0.110 | 2.376 | 0.018 | -0.095 | -2.965 | 0.003 | 28 |
| ENSG00000182087 | transmembrane protein 259 [Source:HGNC Symbol;Acc:HGNC:17039] | TMEM259 | protein_coding | 0.034 | 3.097 | 0.002 | -0.012 | -2.021 | 0.043 | 25 |
| ENSG00000121858 | TNF superfamily member 10 [Source:HGNC Symbol;Acc:HGNC:11925] | TNFSF10 | protein_coding | 0.047 | 2.603 | 0.009 | -0.038 | -2.377 | 0.017 | 28 |
| ENSG00000118194 | troponin T2, cardiac type [Source:HGNC Symbol;Acc:HGNC:11949] | TNNT2 | protein_coding | -0.047 | -2.064 | 0.039 | 0.027 | 2.082 | 0.037 | 25 |
| ENSG00000111077 | tensin 2 [Source:HGNC Symbol;Acc:HGNC:19737] | TNS2 | protein_coding | 0.069 | 2.118 | 0.034 | -0.027 | -1.978 | 0.048 | 26 |
| ENSG00000177302 | DNA topoisomerase III alpha [Source:HGNC Symbol;Acc:HGNC:11992] | TOP3A | protein_coding | -0.073 | -2.714 | 0.007 | 0.072 | 3.268 | 0.001 | 28 |
| ENSG00000166166 | tRNA methyltransferase 61A [Source:HGNC Symbol;Acc:HGNC:23790] | TRMT61A | protein_coding | -0.047 | -2.395 | 0.017 | 0.039 | 2.291 | 0.022 | 28 |
| ENSG00000179981 | teashirt zinc finger homeobox 1 [Source:HGNC Symbol;Acc:HGNC:10669] | TSHZ1 | protein_coding | 0.029 | 2.041 | 0.041 | -0.027 | -1.985 | 0.047 | 29 |

| | | | | | | | | | | |
|---|---|---|---|---|---|---|---|---|---|---|
| ENSG00000160803 | ubiquilin 4 [Source:HGNC Symbol;Acc:HGNC:1237] | UBQLN4 | protein_coding | 0.023 | 2.140 | 0.032 | -0.053 | -3.550 | 0.000 | 32 |
| ENSG00000175970 | unc-119 lipid binding chaperone B [Source:HGNC Symbol;Acc:HGNC:16488] | UNC119B | protein_coding | 0.046 | 2.336 | 0.019 | -0.021 | -1.986 | 0.047 | 26 |
| ENSG00000132478 | unk zinc finger [Source:HGNC Symbol;Acc:HGNC:29369] | UNK | protein_coding | 0.048 | 2.024 | 0.043 | -0.027 | -2.183 | 0.029 | 27 |
| ENSG00000169062 | UPF3A regulator of nonsense mediated mRNA decay [Source:HGNC Symbol;Acc:HGNC:20332] | UPF3A | protein_coding | 0.029 | 1.983 | 0.047 | -0.040 | -1.987 | 0.047 | 32 |
| ENSG00000136878 | ubiquitin specific peptidase 20 [Source:HGNC Symbol;Acc:HGNC:12619] | USP20 | protein_coding | 0.060 | 2.276 | 0.023 | -0.027 | -2.252 | 0.024 | 26 |
| ENSG00000234769 | WASP family homolog 4, pseudogene [Source:HGNC Symbol;Acc:HGNC:14126] | WASH4P | unprocessed_pseudogene | -0.326 | -3.616 | 0.000 | 0.163 | 2.890 | 0.004 | 27 |
| ENSG00000164961 | WASH complex subunit 5 [Source:HGNC Symbol;Acc:HGNC:28984] | WASHC5 | protein_coding | 0.071 | 2.519 | 0.012 | -0.042 | -2.585 | 0.010 | 28 |
| ENSG00000248334 | WAS protein homolog associated with actin, golgi membranes and microtubules pseudogene 2 [Source:HGNC Symbol;Acc:HGNC:32360] | WHAMMP2 | transcribed_unprocessed_pseudogene | 0.074 | 2.566 | 0.010 | -0.062 | -2.701 | 0.007 | 28 |
| ENSG00000152422 | X-ray repair cross complementing 4 [Source:HGNC Symbol;Acc:HGNC:12831] | XRCC4 | protein_coding | 0.051 | 2.884 | 0.004 | -0.037 | -2.044 | 0.041 | 29 |
| ENSG00000239407 | novel transcript | Z68871.1 | lncRNA | -0.132 | -3.035 | 0.002 | 0.034 | 1.971 | 0.049 | 25 |
| ENSG00000166707 | zinc finger CCHC-type containing 18 [Source:HGNC Symbol;Acc:HGNC:32459] | ZCCHC18 | protein_coding | -0.166 | -1.998 | 0.046 | 0.153 | 2.229 | 0.026 | 29 |
| ENSG00000160445 | zyg-11 related cell cycle regulator [Source:HGNC Symbol;Acc:HGNC:30960] | ZER1 | protein_coding | 0.037 | 2.250 | 0.024 | -0.018 | -2.405 | 0.016 | 26 |
| ENSG00000066827 | zinc finger and AT-hook domain containing [Source:HGNC Symbol;Acc:HGNC:19899] | ZFAT | protein_coding | 0.057 | 2.029 | 0.042 | -0.033 | -2.076 | 0.038 | 27 |
| ENSG00000275111 | zinc finger protein 2 [Source:HGNC Symbol;Acc:HGNC:12991] | ZNF2 | protein_coding | -0.075 | -2.204 | 0.028 | 0.046 | 2.863 | 0.004 | 27 |

| | | | | | | | | | | |
|---|---|---|---|---|---|---|---|---|---|---|
| ENSG00000166261 | zinc finger protein 202 [Source:HGNC Symbol;Acc:HGNC:12994] | ZNF202 | protein_coding | -0.058 | -1.971 | 0.049 | 0.090 | 2.635 | 0.008 | 30 |
| ENSG00000198026 | zinc finger protein 335 [Source:HGNC Symbol;Acc:HGNC:15807] | ZNF335 | protein_coding | 0.030 | 2.648 | 0.008 | -0.049 | -2.581 | 0.010 | 32 |
| ENSG00000198597 | zinc finger protein 536 [Source:HGNC Symbol;Acc:HGNC:29025] | ZNF536 | protein_coding | 0.060 | 2.325 | 0.020 | -0.094 | -2.165 | 0.030 | 30 |
| ENSG00000178229 | zinc finger protein 543 [Source:HGNC Symbol;Acc:HGNC:25281] | ZNF543 | protein_coding | -0.029 | -2.066 | 0.039 | 0.026 | 2.346 | 0.019 | 28 |
| ENSG00000230844 | ZNF674 antisense RNA 1 (head to head) [Source:HGNC Symbol;Acc:HGNC:44266] | ZNF674-AS1 | lncRNA | -0.095 | -2.285 | 0.022 | 0.044 | 2.225 | 0.026 | 26 |
| Metabolites | | a-ketoglutarate | metabolite | -0.049 | -0.787 | 0.431 | 0.045 | 1.408 | 0.159 | 29 |
| | | Carnitine C12 | metabolite | 0.018 | 0.131 | 0.896 | 0.077 | 2.195 | **0.028** | 25 |
| | | Carnitine C14:1 | metabolite | 0.020 | 0.233 | 0.816 | 0.089 | 2.791 | **0.005** | 25 |
| | | Carnitine C16 | metabolite | 0.041 | 0.933 | 0.351 | 0.177 | 4.782 | **0.000** | 32 |
| | | Carnitine C16-OH | metabolite | 0.042 | 1.102 | 0.270 | 0.169 | 1.693 | 0.090 | 32 |
| | | Carnitine C18 | metabolite | 0.076 | 0.747 | 0.455 | 0.067 | 2.672 | **0.008** | 25 |
| | | Carnitine C18-OH | metabolite | -0.055 | -0.575 | 0.565 | 0.086 | 2.467 | **0.014** | 24 |
| | | Carnitine C18:1 | metabolite | 0.043 | 0.451 | 0.652 | 0.053 | 1.591 | 0.112 | 26 |
| | | Carnitine C18:1-OH | metabolite | -0.008 | -0.080 | 0.936 | 0.094 | 3.605 | **0.000** | 24 |
| | | Carnitine C18:2 | metabolite | 0.059 | 0.625 | 0.532 | 0.040 | 1.042 | 0.297 | 26 |
| | | Carnitine C18:2-OH | metabolite | 0.047 | 1.104 | 0.270 | 0.224 | 3.257 | **0.001** | 32 |
| | | Glutarylcarnitine (C5DC) | metabolite | -0.024 | -0.163 | 0.871 | 0.062 | 1.484 | 0.138 | 25 |
| | | Carnitine C8:1 | metabolite | 0.063 | 1.095 | 0.273 | 0.027 | 0.359 | 0.719 | 30 |
| | | Ribose-5-phosphate | metabolite | 0.124 | 2.138 | **0.033** | 0.218 | 2.416 | **0.016** | 32 |

Table S3 - Pre- and Post-operative characteristics in samples with transcriptomics and metabolomics analyses. (*) - Tests among BMI groups were conducted by exact test for categorical variables, and ANOVA or non-parametric Kruskal-Wallis test for continuous variables. Data are presented as n (%) for categorical variables and mean (SD) or median (IQR) for continuous variables. Abbreviations: ACE – Angiotensin Converting Enzyme; AKI – Acute Kidney Injury; BMI – Body Mass Index; CABG – Coronary artery Bypass Grafting; CCS – Canadian Cardiovascular Society; Hct – Haematocrit; FiO2 – Fraction of Inspired Oxygen; KDIGO - The Kidney Disease Improving Global Outcomes; MABP – Mean Arterial Blood Pressure; MODS – Multiorgan Dysfunction Syndrome; NYHA – New York Heart Association; PO2 – Partial Pressure of Oxygen; RBC – Red Blood Cells; VD – Vessel Disease.

| Subset (n=60) | BMI<25 (n=16) | 25≥BMI≤32 (n=33) | BMI>32 (n=11) | p-value | Missing data (n) |
|---|---|---|---|---|---|
| Age (years) - Median (1st - 3rd quartile) | 67 (62 - 75) | 67 (61.8 - 74) | 60 (54 - 67.5) | 0.415 | 0 |
| Sex (male) - n (%) | 14 (88%) | 28 (85%) | 10 (91%) | 1.000 | 0 |
| Ethnic (White) - n (%) | 14 (88%) | 28 (85%) | 10 (91%) | 0.336 | 0 |
| BMI | 67 (62 - 75) | 67 (61.8 - 74) | 60 (54 - 67.5) | <0.001 | 0 |
| Smoking History | | | | | |
| Never smoker - n (%) | 6 (38%) | 14 (42%) | 4 (36%) | 0.944 | 0 |
| Ex-smoker - n (%) | 8 (50%) | 16 (48%) | 5 (45%) | | |
| Current smoker - n (%) | 2 (12%) | 3 (9%) | 2 (18%) | | |
| **Medical history** | | | | | |
| Diabetes - n (%) | 2 (12%) | 9 (27%) | 4 (36%) | 0.342 | 0 |
| Permanent Pacemaker - n (%) | 1 (6%) | 1 (3%) | 0 (0%) | 1 | 0 |
| Stroke/Transient Ischaemic Attack - n (%) | 2 (12%) | 3 (9%) | 1 (9%) | 1 | 0 |
| Chronic pulmonary disease - n (%) | 3 (19%) | 3 (9%) | 2 (18%) | 0.516 | 0 |
| Neurological disease - n (%) | 0 (0%) | 0 (0%) | 0 (0%) | -- | 0 |
| Renal disease - n (%) | 0 (0%) | 1 (3%) | 2 (18%) | 0.139 | 0 |
| Myocardial infarction - n (%) | 5 (31%) | 8 (24%) | 1 (9%) | 0.464 | 0 |
| Extracardiac arteriopathy - n (%) | 2 (12%) | 4 (12%) | 1 (9%) | 1 | 0 |
| Liver disease - n (%) | 0 (0%) | 0 (0%) | 0 (0%) | -- | 0 |
| Pulmonary hypertension - n (%) | 0 (0%) | 1 (3%) | 0 (0%) | 1 | 0 |
| **Pre-operative Medication Histroy** | | | | | |
| Statin - n (%) | 12 (75%) | 23 (70%) | 10 (91%) | 0.476 | 0 |
| Anti-platelet agents - n (%) | 11 (69%) | 28 (85%) | 10 (91%) | 0.354 | 0 |
| ACE inhibitors - n (%) | 9 (56%) | 14 (42%) | 5 (45%) | 0.691 | 0 |
| **Clinical characteristics** | | | | | |
| Surgery type | | | | | |
| CABG only - n (%) | 13 (81%) | 29 (88%) | 10 (91%) | 0.772 | 0 |
| CABG & Valve - n (%) | 3 (19%) | 4 (12%) | 1 (9%) | | |
| NYHA | | | | | |
| Class I - n (%) | 4 (25%) | 10 (30%) | 4 (36%) | 0.773 | 0 |
| Class II - n (%) | 11 (69%) | 20 (61%) | 5 (45%) | | |
| Class III, IV - n (%) | 1 (6%) | 3 (9%) | 2 (18%) | | |
| CCS | | | | | |
| Asymptomatic - n (%) | 5 (31%) | 2 (6%) | 2 (18%) | 0.341 | 0 |
| Class I - n (%) | 6 (38%) | 11 (33%) | 4 (36%) | | |

| | | | | | |
|---|---|---|---|---|---|
| Class II - n (%) | 4 (25%) | 16 (48%) | 4 (36%) | | |
| Class III, IV - n (%) | 1 (6%) | 4 (12%) | 1 (9%) | | |
| Left Ventricular Ejection Fraction | | | | | |
| Good (>49%) - n (%) | 13 (81%) | 27 (82%) | 8 (73%) | 0.827 | 0 |
| Fair (30-49%) - n (%) | 3 (19%) | 6 (18%) | 3 (27%) | | |
| Left main stem disease - n (%) | 2 (12%) | 9 (27%) | 2 (18%) | 0.535 | 0 |
| Extent of coronary disease | | | | | |
| Normal/ 1VD - n (%) | 1 (6%) | 1 (3%) | 3 (27%) | 0.04 | 0 |
| 2VD - n (%) | 7 (44%) | 6 (18%) | 2 (18%) | 0.04 | 0 |
| 3VD - n (%) | 8 (50%) | 26 (79%) | 6 (55%) | 0.04 | 0 |
| Pre-operative PaO2/FiO2 ratio - Median (1st - 3rd quartile) | 533 (445.2 - 690.5) | 495 (409.5 - 533.3) | 457 (409.5 - 457.1) | 0.280 | 8 |
| Pre-operative Platelets count (x109/L) - Mean (STD) | 222.2 (55.7) | 232.6 (64.9) | 231.5 (63.3) | 0.858 | 1 |
| Pre-operative Serum Creatinine (umol/L) - Median (1st - 3rd quartile) | 78 (70.8 - 102.5) | 77 (68 - 87) | 84 (77 - 92) | 0.491 | 0 |
| Pre-operative Bilirubin (umol/L) - Median (1st - 3rd quartile) | 10 (7.5 - 12.5) | 11 (8.5 - 12.5) | 8 (8 - 12) | 0.801 | 5 |
| Postoperative | | | | | |
| Hct (%) - Mean (STD) | 31.9 (3.9) | 34.6 (3.9) | 35 (4.9) | 0.076 | 0 |
| MABP (mm Hg) - Median (1st - 3rd quartile) | 76 (68.8 - 85) | 73 (64 - 76) | 65 (61.5 - 72) | 0.672 | 0 |
| Lactate (mmol/L) - Median (1st - 3rd quartile) | 2 (1.1 - 2) | 2 (1.4 - 2.2) | 2 (1.3 - 2) | 0.970 | 1 |
| Inotropic score at 24h - Median (1st - 3rd quartile) | 0 (0 - 2) | 1 (0 - 3) | 0 (0 - 3) | 0.198 | 7 |
| Vasoactive score at 24h - Median (1st - 3rd quartile) | 5 (2.5 - 9) | 5 (2.5 - 7) | 4 (2 - 7.5) | 0.198 | 4 |
| MODS ICU - Median (1st - 3rd quartile) | 1 (1 - 3) | 2 (2 - 3) | 3 (2 - 3) | 0.044 | 2 |
| Worst postoperative MODS score - Median (1st - 3rd quartile) | 2 (1 - 5) | 3 (2 - 4) | 3 (3 - 4) | 0.023 | 1 |
| PaO2/FiO2 ratio at 48h - Median (1st - 3rd quartile) | 410 (342.1 - 572.6) | 395 (307.1 - 409.5) | 410 (342.3 - 457.1) | 0.127 | 1 |
| Serum creatinine 48h (umol/L) - Median (1st - 3rd quartile) | 73 (67.5 - 90) | 71 (60 - 82) | 74 (72 - 76) | 0.383 | 2 |
| RBC transfused postoperative - n (%) | 8 (50%) | 12 (36%) | 2 (18%) | 0.274 | 0 |
| nonRBC transfusion at more than 48h - n (%) | 2 (12%) | 4 (12%) | 1 (9%) | 1 | 0 |
| nonRBC transfusion within 48h - n (%) | 6 (38%) | 6 (18%) | 0 (0%) | 0.046 | 1 |
| PaO2/FiO2 ratio at 48hr <=300 - n (%) | 3 (19%) | 4 (12%) | 2 (18%) | 0.7 | 1 |
| AKI according to kdigo criteria - n (%) | 1 (6%) | 1 (3%) | 0 (0%) | 1 | 1 |

Table S4 – Details of the pathway analysis. Number of genes indicates number of transcripts in the analyzed dataset. Direction indicates gene set’s up or downregulation.

| Pathway | 25≥BMI≤32 vs BMI<25 | | | | BMI>32 vs BMI<25 | | | | BMI>32 vs25≥BMI≤32 | | | |
|---|---|---|---|---|---|---|---|---|---|---|---|---|
| | Number of Genes | Direction | p-value | Adjusted p-value | Number of Genes | Direction | p-value | Adjusted p-value | Number of Genes | Direction | p-value | Adjusted p-value |
| Metabolism of amino acids and derivatives | 327 | Down | 0.001 | 0.034 | NA | NA | NA | NA | NA | NA | NA | NA |
| Translation | 292 | Down | 0.000 | 0.004 | NA | NA | NA | NA | NA | NA | NA | NA |
| Signaling by ROBO receptors | 207 | Down | 0.000 | 0.004 | 207 | Down | 5.8E-04 | 0.034 | NA | NA | NA | NA |
| rRNA processing | 203 | Down | 0.000 | 0.000 | 203 | Down | 7.2E-05 | 0.006 | NA | NA | NA | NA |
| rRNA processing in the nucleus and cytosol | 191 | Down | 0.000 | 0.000 | 191 | Down | 3.1E-05 | 0.003 | NA | NA | NA | NA |
| Major pathway of rRNA processing in the nucleolus and cytosol | 181 | Down | 0.000 | 0.000 | 181 | Down | 1.5E-05 | 0.002 | NA | NA | NA | NA |
| Regulation of expression of SLITs and ROBOs | 163 | Down | 0.000 | 0.000 | 163 | Down | 7.7E-05 | 0.006 | NA | NA | NA | NA |
| Influenza Infection | 154 | Down | 0.000 | 0.000 | 154 | Down | 2.4E-06 | 0.000 | NA | NA | NA | NA |
| Influenza Viral RNA Transcription and Replication | 135 | Down | 0.000 | 0.000 | 135 | Down | 1.5E-07 | 0.000 | NA | NA | NA | NA |
| Cap-dependent Translation Initiation | 118 | Down | 0.000 | 0.000 | 118 | Down | 9.4E-10 | 0.000 | NA | NA | NA | NA |
| Eukaryotic Translation Initiation | 118 | Down | 0.000 | 0.000 | 118 | Down | 9.4E-10 | 0.000 | NA | NA | NA | NA |
| Nonsense Mediated Decay (NMD) enhanced by the Exon Junction Complex (EJC) | 114 | Down | 0.000 | 0.000 | 114 | Down | 1.1E-09 | 0.000 | NA | NA | NA | NA |
| Nonsense-Mediated Decay (NMD) | 114 | Down | 0.000 | 0.000 | 114 | Down | 1.1E-09 | 0.000 | NA | NA | NA | NA |
| Selenoamino acid metabolism | 114 | Down | 0.000 | 0.000 | 114 | Down | 1.2E-11 | 0.000 | NA | NA | NA | NA |
| GTP hydrolysis and joining of the 60S ribosomal subunit | 111 | Down | 0.000 | 0.000 | 111 | Down | 6.2E-11 | 0.000 | NA | NA | NA | NA |
| SRP-dependent cotranslational protein targeting to membrane | 111 | Down | 0.000 | 0.000 | 111 | Down | 2.2E-12 | 0.000 | NA | NA | NA | NA |
| L13a-mediated translational silencing of Ceruloplasmin expression | 110 | Down | 0.000 | 0.000 | 110 | Down | 1.1E-11 | 0.000 | NA | NA | NA | NA |
| Formation of a pool of free 40S subunits | 100 | Down | 0.000 | 0.000 | 100 | Down | 2.8E-13 | 0.000 | NA | NA | NA | NA |
| Response of EIF2AK4 (GCN2) to amino acid deficiency | 100 | Down | 0.000 | 0.000 | 100 | Down | 4.2E-13 | 0.000 | NA | NA | NA | NA |
| Nonsense Mediated Decay (NMD) independent of the Exon Junction Complex (EJC) | 94 | Down | 0.000 | 0.000 | 94 | Down | 3.1E-14 | 0.000 | NA | NA | NA | NA |

| | | | | | | | | | | | | |
|---|---|---|---|---|---|---|---|---|---|---|---|---|
| Eukaryotic Translation Elongation | 92 | Down | 0.000 | 0.000 | 92 | Down | 1.1E-14 | 0.000 | NA | NA | NA | NA |
| Eukaryotic Translation Termination | 92 | Down | 0.000 | 0.000 | 92 | Down | 2.3E-13 | 0.000 | NA | NA | NA | NA |
| Selenocysteine synthesis | 92 | Down | 0.000 | 0.000 | 92 | Down | 3.1E-14 | 0.000 | NA | NA | NA | NA |
| Peptide chain elongation | 88 | Down | 0.000 | 0.000 | 88 | Down | 6.9E-15 | 0.000 | NA | NA | NA | NA |
| Viral mRNA Translation | 88 | Down | 0.000 | 0.000 | 88 | Down | 5.7E-15 | 0.000 | NA | NA | NA | NA |
| Activation of the mRNA upon binding of the cap-binding complex and eIFs, and subsequent binding to 43S | 59 | Down | 0.000 | 0.000 | 59 | Down | 1.3E-05 | 0.001 | NA | NA | NA | NA |
| Ribosomal scanning and start codon recognition | 58 | Down | 0.000 | 0.000 | 58 | Down | 5.3E-05 | 0.005 | NA | NA | NA | NA |
| Translation initiation complex formation | 58 | Down | 0.000 | 0.000 | 58 | Down | 1.1E-05 | 0.001 | NA | NA | NA | NA |
| Interferon alpha/beta signaling | 56 | Up | 0.000 | 0.008 | NA | NA | NA | NA | NA | NA | NA | NA |
| Formation of the ternary complex, and subsequently, the 43S complex | 51 | Down | 0.000 | 0.000 | 51 | Down | 1.3E-06 | 0.000 | NA | NA | NA | NA |
| Iron uptake and transport | 50 | Down | 0.000 | 0.017 | NA | NA | NA | NA | NA | NA | NA | NA |
| Striated Muscle Contraction | 29 | Up | 0.000 | 0.004 | 29 | Up | 1.5E-04 | 0.012 | NA | NA | NA | NA |
| Synthesis, secretion, and deacylation of Ghrelin | 12 | Down | 0.000 | 0.011 | NA | NA | NA | NA | NA | NA | NA | NA |
| mitochondrial fatty acid beta-oxidation of saturated fatty acids | 10 | Up | 0.000 | 0.026 | NA | NA | NA | NA | NA | NA | NA | NA |
| Defective F8 binding to von Willebrand factor | 2 | Up | 0.000 | 0.017 | NA | NA | NA | NA | NA | NA | NA | NA |
| Defective F8 cleavage by thrombin | 2 | Up | 0.000 | 0.017 | NA | NA | NA | NA | NA | NA | NA | NA |
| HSF1-dependent transactivation | NA | NA | NA | NA | 35 | Up | 2.2E-04 | 0.014 | NA | NA | NA | NA |
| Keratan sulfate degradation | NA | NA | NA | NA | 12 | Down | 3.3E-04 | 0.021 | NA | NA | NA | NA |
| Erythrocytes take up oxygen and release carbon dioxide | NA | NA | NA | NA | 8 | Up | 6.8E-05 | 0.006 | NA | NA | NA | NA |
| TNFR1-mediated ceramide production | NA | NA | NA | NA | 6 | Down | 8.2E-04 | 0.046 | NA | NA | NA | NA |
| Alternative complement activation | NA | NA | NA | NA | 4 | Down | 4.6E-04 | 0.028 | NA | NA | NA | NA |
| Ribonuclease P activity (lncRNA, Gene Ontology) | NA | NA | NA | NA | 2 | Down | 2.1E-04 | 0.014 | NA | NA | NA | NA |
| tRNA processing (lncRNA, Gene Ontology) | NA | NA | NA | NA | 2 | Down | 2.1E-04 | 0.014 | NA | NA | NA | NA |
| ribonuclease P complex (lncRNA, Gene Ontology) | NA | NA | NA | NA | 2 | Down | 2.1E-04 | 0.014 | NA | NA | NA | NA |

| | | | | | | | | | | | | |
|---|---|---|---|---|---|---|---|---|---|---|---|---|
| RNA phosphodiester bond hydrolysis, endonucleolytic (lncRNA, Gene Ontology) | NA | NA | NA | NA | 2 | Down | 2.1E-04 | 0.014 | NA | NA | NA | NA |
| Defective ACTH causes Obesity and Pro-opiomelanocortinin deficiency (POMCD) | NA | NA | NA | NA | 1 | Up | 7.3E-04 | 0.042 | NA | NA | NA | NA |
| Binding and Uptake of Ligands by Scavenger Receptors | NA | NA | NA | NA | NA | NA | NA | NA | 83 | Up | 2.3E-05 | 5.6E-03 |
| Nucleolus (lncRNA, Gene Ontology) | NA | NA | NA | NA | 83 | Down | 1.9E-09 | 0.000 | 83 | Down | 4.0E-06 | 1.6E-03 |
| FCGR3A-mediated IL10 synthesis | NA | NA | NA | NA | NA | NA | NA | NA | 82 | Up | 8.8E-05 | 1.5E-02 |
| RNA processing (lncRNA, Gene Ontology) | NA | NA | NA | NA | 81 | Down | 1.1E-09 | 0.000 | 81 | Down | 2.8E-06 | 1.4E-03 |
| Antigen activates B Cell Receptor (BCR) leading to generation of second messengers | NA | NA | NA | NA | NA | NA | NA | NA | 74 | Up | 9.8E-06 | 3.1E-03 |
| FCERI mediated Ca+2 mobilization | NA | NA | NA | NA | NA | NA | NA | NA | 74 | Up | 1.6E-04 | 2.4E-02 |
| Role of phospholipids in phagocytosis | NA | NA | NA | NA | NA | NA | NA | NA | 70 | Up | 1.3E-04 | 2.0E-02 |
| Role of LAT2/NTAL/LAB on calcium mobilization | NA | NA | NA | NA | NA | NA | NA | NA | 59 | Up | 2.6E-05 | 5.8E-03 |
| FCGR activation | NA | NA | NA | NA | 58 | Up | 4.8E-04 | 0.029 | 58 | Up | 3.6E-07 | 2.4E-04 |
| Creation of C4 and C2 activators | NA | NA | NA | NA | NA | NA | NA | NA | 56 | Up | 1.0E-05 | 3.1E-03 |
| Scavenging of heme from plasma | NA | NA | NA | NA | 55 | Up | 2.0E-04 | 0.014 | 55 | Up | 3.9E-07 | 2.4E-04 |
| Classical antibody-mediated complement activation | NA | NA | NA | NA | NA | NA | NA | NA | 51 | Up | 1.4E-05 | 3.8E-03 |
| CD22 mediated BCR regulation | NA | NA | NA | NA | 50 | Up | 9.8E-05 | 0.008 | 50 | Up | 8.4E-08 | 1.7E-04 |
| Cholesterol biosynthesis | NA | NA | NA | NA | NA | NA | NA | NA | 24 | Up | 1.4E-07 | 1.7E-04 |
| Acetylcholine regulates insulin secretion | NA | NA | NA | NA | NA | NA | NA | NA | 10 | Down | 2.8E-04 | 4.0E-02 |
| Metallothioneins bind metals | NA | NA | NA | NA | NA | NA | NA | NA | 6 | Up | 5.7E-05 | 1.1E-02 |
| Peptide hormone biosynthesis | NA | NA | NA | NA | NA | NA | NA | NA | 5 | Up | 2.9E-05 | 5.9E-03 |

Table S5 – Details of genes that were highly variable between the BMI groups (p-value < 0.05) with weighted gene correlation networks membership.

| Ensembl ID | Gene Symbol | Description | Biotype | 25≥BMI≤32 vs BMI<25 | BMI>32 vs BMI<25 | BMI>32 vs25≥BMI≤32 | AveExpr | F | P.Value | Network |
|---|---|---|---|---|---|---|---|---|---|---|
| ENSG00000008516 | MMP25 | matrix metallopeptidase 25 [Source:HGNC Symbol;Acc:HGNC:14246] | protein_coding | 0.660 | 1.483 | 0.823 | -0.840 | 3.588 | 0.034 | black |
| ENSG00000044524 | EPHA3 | EPH receptor A3 [Source:HGNC Symbol;Acc:HGNC:3387] | protein_coding | 0.332 | 0.303 | -0.029 | 4.173 | 3.281 | 0.045 | black |
| ENSG00000081059 | TCF7 | transcription factor 7 [Source:HGNC Symbol;Acc:HGNC:11639] | protein_coding | 0.692 | 0.382 | -0.310 | 0.683 | 4.538 | 0.015 | black |
| ENSG00000103855 | CD276 | CD276 molecule [Source:HGNC Symbol;Acc:HGNC:19137] | protein_coding | -0.160 | -0.335 | -0.175 | 3.597 | 4.537 | 0.015 | black |
| ENSG00000112195 | TREML2 | triggering receptor expressed on myeloid cells like 2 [Source:HGNC Symbol;Acc:HGNC:21092] | protein_coding | 0.561 | 1.663 | 1.102 | -2.836 | 6.854 | 0.002 | black |
| ENSG00000115607 | IL18RAP | interleukin 18 receptor accessory protein [Source:HGNC Symbol;Acc:HGNC:5989] | protein_coding | 1.019 | 1.721 | 0.702 | -1.977 | 6.032 | 0.004 | black |
| ENSG00000128383 | APOBEC3A | apolipoprotein B mRNA editing enzyme catalytic subunit 3A [Source:HGNC Symbol;Acc:HGNC:17343] | protein_coding | 0.719 | 1.443 | 0.724 | -2.212 | 3.213 | 0.048 | black |
| ENSG00000134909 | ARHGAP32 | Rho GTPase activating protein 32 [Source:HGNC Symbol;Acc:HGNC:17399] | protein_coding | -0.330 | -0.274 | 0.055 | 2.964 | 3.300 | 0.044 | black |
| ENSG00000151789 | ZNF385D | zinc finger protein 385D [Source:HGNC Symbol;Acc:HGNC:26191] | protein_coding | -0.849 | -0.948 | -0.099 | 1.222 | 5.439 | 0.007 | black |
| ENSG00000151948 | GLT1D1 | glycosyltransferase 1 domain containing 1 [Source:HGNC Symbol;Acc:HGNC:26483] | protein_coding | 0.500 | 1.431 | 0.931 | -2.593 | 3.845 | 0.027 | black |
| ENSG00000157551 | KCNJ15 | potassium inwardly rectifying channel subfamily J member 15 [Source:HGNC Symbol;Acc:HGNC:6261] | protein_coding | 0.200 | 1.144 | 0.945 | -0.392 | 3.709 | 0.031 | black |
| ENSG00000158517 | NCF1 | neutrophil cytosolic factor 1 [Source:HGNC Symbol;Acc:HGNC:7660] | protein_coding | 0.444 | 0.984 | 0.540 | 0.283 | 3.298 | 0.045 | black |
| ENSG00000162676 | GFI1 | growth factor independent 1 transcriptional repressor [Source:HGNC Symbol;Acc:HGNC:4237] | protein_coding | 1.015 | 0.148 | -0.866 | -1.992 | 3.888 | 0.026 | black |
| ENSG00000186529 | CYP4F3 | cytochrome P450 family 4 subfamily F member 3 [Source:HGNC Symbol;Acc:HGNC:2646] | protein_coding | 0.466 | 1.604 | 1.137 | -1.468 | 3.947 | 0.025 | black |
| ENSG00000231259 | ANAPC1P2 | ANAPC1 pseudogene 2 [Source:HGNC Symbol;Acc:HGNC:54708] | unprocessed_pseudogene | 0.406 | 2.080 | 1.674 | -3.395 | 3.707 | 0.031 | black |
| ENSG00000280132 | AC026471.6 | novel transcript | TEC | -0.819 | -0.471 | 0.347 | -2.296 | 3.240 | 0.047 | black |
| ENSG00000005187 | ACSM3 | acyl-CoA synthetase medium chain family member 3 [Source:HGNC Symbol;Acc:HGNC:10522] | protein_coding | 0.390 | 0.550 | 0.159 | 2.505 | 3.933 | 0.025 | blue |
| ENSG00000007908 | SELE | selectin E [Source:HGNC Symbol;Acc:HGNC:10718] | protein_coding | -0.227 | 1.951 | 2.178 | 0.162 | 4.623 | 0.014 | blue |
| ENSG00000008300 | CELSR3 | cadherin EGF LAG seven-pass G-type receptor 3 [Source:HGNC Symbol;Acc:HGNC:3230] | protein_coding | 0.549 | 1.329 | 0.781 | -1.310 | 3.517 | 0.037 | blue |
| ENSG00000011304 | PTBP1 | polypyrimidine tract binding protein 1 [Source:HGNC Symbol;Acc:HGNC:9583] | protein_coding | 0.003 | 0.332 | 0.330 | 4.042 | 3.188 | 0.049 | blue |

| | | | | | | | | | | |
|---|---|---|---|---|---|---|---|---|---|---|
| ENSG00000050327 | ARHGEF5 | Rho guanine nucleotide exchange factor 5 [Source:HGNC Symbol;Acc:HGNC:13209] | protein_coding | 0.132 | -0.309 | -0.441 | 1.745 | 3.362 | 0.042 | blue |
| ENSG00000054282 | SDCCAG8 | serologically defined colon cancer antigen 8 [Source:HGNC Symbol;Acc:HGNC:10671] | protein_coding | -0.209 | -0.057 | 0.151 | 3.703 | 3.171 | 0.050 | blue |
| ENSG00000069399 | BCL3 | BCL3 transcription coactivator [Source:HGNC Symbol;Acc:HGNC:998] | protein_coding | -0.387 | 0.194 | 0.581 | 1.358 | 3.928 | 0.026 | blue |
| ENSG00000072182 | ASIC4 | acid sensing ion channel subunit family member 4 [Source:HGNC Symbol;Acc:HGNC:21263] | protein_coding | 1.217 | 0.334 | -0.883 | -2.353 | 3.435 | 0.039 | blue |
| ENSG00000081148 | IMPG2 | interphotoreceptor matrix proteoglycan 2 [Source:HGNC Symbol;Acc:HGNC:18362] | protein_coding | -0.299 | 0.132 | 0.431 | -0.159 | 3.237 | 0.047 | blue |
| ENSG00000087074 | PPP1R15A | protein phosphatase 1 regulatory subunit 15A [Source:HGNC Symbol;Acc:HGNC:14375] | protein_coding | -0.183 | 0.259 | 0.442 | 5.265 | 4.776 | 0.012 | blue |
| ENSG00000099860 | GADD45B | growth arrest and DNA damage inducible beta [Source:HGNC Symbol;Acc:HGNC:4096] | protein_coding | -0.279 | 0.422 | 0.701 | 4.460 | 3.543 | 0.036 | blue |
| ENSG00000100450 | GZMH | granzyme H [Source:HGNC Symbol;Acc:HGNC:4710] | protein_coding | 0.795 | 0.374 | -0.421 | -1.574 | 3.418 | 0.040 | blue |
| ENSG00000102390 | PBDC1 | polysaccharide biosynthesis domain containing 1 [Source:HGNC Symbol;Acc:HGNC:28790] | protein_coding | -0.313 | -0.327 | -0.014 | 0.752 | 3.293 | 0.045 | blue |
| ENSG00000104368 | PLAT | plasminogen activator, tissue type [Source:HGNC Symbol;Acc:HGNC:9051] | protein_coding | -0.341 | 0.103 | 0.444 | 3.573 | 3.830 | 0.028 | blue |
| ENSG00000104490 | NCALD | neurocalcin delta [Source:HGNC Symbol;Acc:HGNC:7655] | protein_coding | -0.247 | 0.101 | 0.349 | 3.042 | 3.462 | 0.039 | blue |
| ENSG00000105835 | NAMPT | nicotinamide phosphoribosyltransferase [Source:HGNC Symbol;Acc:HGNC:30092] | protein_coding | -0.232 | 0.135 | 0.366 | 6.281 | 3.584 | 0.035 | blue |
| ENSG00000108342 | CSF3 | colony stimulating factor 3 [Source:HGNC Symbol;Acc:HGNC:2438] | protein_coding | 0.090 | 1.546 | 1.457 | -3.534 | 3.477 | 0.038 | blue |
| ENSG00000108691 | CCL2 | C-C motif chemokine ligand 2 [Source:HGNC Symbol;Acc:HGNC:10618] | protein_coding | -0.557 | 0.424 | 0.981 | 4.125 | 3.413 | 0.040 | blue |
| ENSG00000109610 | SOD3 | superoxide dismutase 3 [Source:HGNC Symbol;Acc:HGNC:11181] | protein_coding | 0.242 | 0.033 | -0.210 | 3.881 | 4.076 | 0.022 | blue |
| ENSG00000111424 | VDR | vitamin D receptor [Source:HGNC Symbol;Acc:HGNC:12679] | protein_coding | -0.886 | -0.412 | 0.474 | -1.088 | 3.682 | 0.032 | blue |
| ENSG00000111912 | NCOA7 | nuclear receptor coactivator 7 [Source:HGNC Symbol;Acc:HGNC:21081] | protein_coding | -0.350 | 0.208 | 0.558 | 3.768 | 4.516 | 0.015 | blue |
| ENSG00000113356 | POLR3G | RNA polymerase III subunit G [Source:HGNC Symbol;Acc:HGNC:30075] | protein_coding | -0.699 | 0.107 | 0.806 | -0.682 | 4.681 | 0.013 | blue |
| ENSG00000115665 | SLC5A7 | solute carrier family 5 member 7 [Source:HGNC Symbol;Acc:HGNC:14025] | protein_coding | 0.759 | 0.382 | -0.377 | -0.730 | 3.882 | 0.027 | blue |
| ENSG00000117036 | ETV3 | ETS variant transcription factor 3 [Source:HGNC Symbol;Acc:HGNC:3492] | protein_coding | -0.171 | 0.017 | 0.188 | 3.442 | 3.613 | 0.034 | blue |
| ENSG00000117479 | SLC19A2 | solute carrier family 19 member 2 [Source:HGNC Symbol;Acc:HGNC:10938] | protein_coding | -0.328 | 0.112 | 0.441 | 3.129 | 3.694 | 0.031 | blue |
| ENSG00000119231 | SENP5 | SUMO specific peptidase 5 [Source:HGNC Symbol;Acc:HGNC:28407] | protein_coding | -0.236 | -0.054 | 0.182 | 4.128 | 4.827 | 0.012 | blue |

| | | | | | | | | | | |
|---|---|---|---|---|---|---|---|---|---|---|
| ENSG00000121101 | TEX14 | testis expressed 14, intercellular bridge forming factor [Source:HGNC Symbol;Acc:HGNC:11737] | protein_coding | -0.623 | 0.621 | 1.244 | -2.762 | 3.489 | 0.038 | blue |
| ENSG00000124831 | LRRFIP1 | LRR binding FLII interacting protein 1 [Source:HGNC Symbol;Acc:HGNC:6702] | protein_coding | 0.121 | 0.282 | 0.161 | 5.483 | 5.830 | 0.005 | blue |
| ENSG00000125148 | MT2A | metallothionein 2A [Source:HGNC Symbol;Acc:HGNC:7406] | protein_coding | -0.267 | 0.262 | 0.529 | 3.039 | 3.349 | 0.043 | blue |
| ENSG00000131873 | CHSY1 | chondroitin sulfate synthase 1 [Source:HGNC Symbol;Acc:HGNC:17198] | protein_coding | -0.113 | 0.222 | 0.335 | 4.054 | 3.919 | 0.026 | blue |
| ENSG00000132002 | DNAJB1 | DnaJ heat shock protein family (Hsp40) member B1 [Source:HGNC Symbol;Acc:HGNC:5270] | protein_coding | -0.152 | 0.106 | 0.258 | 4.141 | 3.444 | 0.039 | blue |
| ENSG00000132680 | KHDC4 | KH domain containing 4, pre-mRNA splicing factor [Source:HGNC Symbol;Acc:HGNC:29145] | protein_coding | 0.187 | 0.072 | -0.115 | 3.430 | 3.224 | 0.048 | blue |
| ENSG00000133874 | RNF122 | ring finger protein 122 [Source:HGNC Symbol;Acc:HGNC:21147] | protein_coding | -0.335 | 0.163 | 0.498 | 1.842 | 4.665 | 0.014 | blue |
| ENSG00000136011 | STAB2 | stabilin 2 [Source:HGNC Symbol;Acc:HGNC:18629] | protein_coding | 0.746 | 1.809 | 1.063 | 0.113 | 5.282 | 0.008 | blue |
| ENSG00000138166 | DUSP5 | dual specificity phosphatase 5 [Source:HGNC Symbol;Acc:HGNC:3071] | protein_coding | -0.192 | 0.635 | 0.827 | 1.719 | 4.758 | 0.013 | blue |
| ENSG00000140379 | BCL2A1 | BCL2 related protein A1 [Source:HGNC Symbol;Acc:HGNC:991] | protein_coding | -0.450 | 0.792 | 1.243 | -1.444 | 4.978 | 0.010 | blue |
| ENSG00000140406 | TLNRD1 | talin rod domain containing 1 [Source:HGNC Symbol;Acc:HGNC:13519] | protein_coding | -0.320 | -0.037 | 0.283 | 2.440 | 3.270 | 0.046 | blue |
| ENSG00000141698 | NT5C3B | 5'-nucleotidase, cytosolic IIIB [Source:HGNC Symbol;Acc:HGNC:28300] | protein_coding | -0.205 | -0.260 | -0.054 | 2.528 | 3.594 | 0.034 | blue |
| ENSG00000143320 | CRABP2 | cellular retinoic acid binding protein 2 [Source:HGNC Symbol;Acc:HGNC:2339] | protein_coding | -0.678 | 0.122 | 0.801 | -1.537 | 3.293 | 0.045 | blue |
| ENSG00000157557 | ETS2 | ETS proto-oncogene 2, transcription factor [Source:HGNC Symbol;Acc:HGNC:3489] | protein_coding | -0.146 | 0.181 | 0.327 | 4.453 | 4.183 | 0.020 | blue |
| ENSG00000159167 | STC1 | stanniocalcin 1 [Source:HGNC Symbol;Acc:HGNC:11373] | protein_coding | 0.045 | 0.783 | 0.737 | 1.819 | 5.021 | 0.010 | blue |
| ENSG00000159339 | PADI4 | peptidyl arginine deiminase 4 [Source:HGNC Symbol;Acc:HGNC:18368] | protein_coding | 0.310 | 1.329 | 1.019 | -2.660 | 3.572 | 0.035 | blue |
| ENSG00000160785 | SLC25A44 | solute carrier family 25 member 44 [Source:HGNC Symbol;Acc:HGNC:29036] | protein_coding | -0.184 | 0.056 | 0.240 | 3.205 | 4.932 | 0.011 | blue |
| ENSG00000163159 | VPS72 | vacuolar protein sorting 72 homolog [Source:HGNC Symbol;Acc:HGNC:11644] | protein_coding | 0.102 | -0.107 | -0.209 | 2.964 | 3.568 | 0.035 | blue |
| ENSG00000163638 | ADAMTS9 | ADAM metallopeptidase with thrombospondin type 1 motif 9 [Source:HGNC Symbol;Acc:HGNC:13202] | protein_coding | 0.077 | 0.746 | 0.669 | 2.790 | 6.067 | 0.004 | blue |
| ENSG00000163659 | TIPARP | TCDD inducible poly(ADP-ribose) polymerase [Source:HGNC Symbol;Acc:HGNC:23696] | protein_coding | -0.233 | 0.131 | 0.364 | 4.737 | 5.080 | 0.010 | blue |
| ENSG00000163909 | HEYL | hes related family bHLH transcription factor with YRPW motif like [Source:HGNC Symbol;Acc:HGNC:4882] | protein_coding | 0.090 | 0.279 | 0.188 | 4.254 | 3.662 | 0.032 | blue |
| ENSG00000164120 | HPGD | 15-hydroxyprostaglandin dehydrogenase [Source:HGNC Symbol;Acc:HGNC:5154] | protein_coding | -0.789 | -0.153 | 0.636 | -0.951 | 6.485 | 0.003 | blue |

| | | | | | | | | | | |
|---|---|---|---|---|---|---|---|---|---|---|
| ENSG00000164949 | GEM | GTP binding protein overexpressed in skeletal muscle [Source:HGNC Symbol;Acc:HGNC:4234] | protein_coding | -0.325 | 0.352 | 0.678 | 3.652 | 3.528 | 0.036 | blue |
| ENSG00000165030 | NFIL3 | nuclear factor, interleukin 3 regulated [Source:HGNC Symbol;Acc:HGNC:7787] | protein_coding | -0.308 | 0.196 | 0.504 | 3.114 | 4.548 | 0.015 | blue |
| ENSG00000171236 | LRG1 | leucine rich alpha-2-glycoprotein 1 [Source:HGNC Symbol;Acc:HGNC:29480] | protein_coding | -0.327 | 1.237 | 1.564 | -2.058 | 6.716 | 0.002 | blue |
| ENSG00000171617 | ENC1 | ectodermal-neural cortex 1 [Source:HGNC Symbol;Acc:HGNC:3345] | protein_coding | -0.311 | 0.080 | 0.391 | 2.751 | 3.936 | 0.025 | blue |
| ENSG00000172530 | BANP | BTG3 associated nuclear protein [Source:HGNC Symbol;Acc:HGNC:13450] | protein_coding | -0.187 | 0.272 | 0.459 | 2.138 | 3.810 | 0.028 | blue |
| ENSG00000172602 | RND1 | Rho family GTPase 1 [Source:HGNC Symbol;Acc:HGNC:18314] | protein_coding | -0.032 | 1.742 | 1.774 | -1.368 | 6.852 | 0.002 | blue |
| ENSG00000172831 | CES2 | carboxylesterase 2 [Source:HGNC Symbol;Acc:HGNC:1864] | protein_coding | 0.136 | -0.023 | -0.159 | 4.210 | 3.635 | 0.033 | blue |
| ENSG00000173530 | TNFRSF10D | TNF receptor superfamily member 10d [Source:HGNC Symbol;Acc:HGNC:11907] | protein_coding | -0.141 | 0.364 | 0.505 | 1.411 | 3.238 | 0.047 | blue |
| ENSG00000178726 | THBD | thrombomodulin [Source:HGNC Symbol;Acc:HGNC:11784] | protein_coding | -0.259 | 0.304 | 0.563 | 3.371 | 4.473 | 0.016 | blue |
| ENSG00000179082 | C9orf106 | chromosome 9 putative open reading frame 106 [Source:HGNC Symbol;Acc:HGNC:31370] | lncRNA | 0.727 | -0.628 | -1.355 | -1.877 | 7.572 | 0.001 | blue |
| ENSG00000180596 | H2BC4 | H2B clustered histone 4 [Source:HGNC Symbol;Acc:HGNC:4757] | protein_coding | 0.177 | -0.028 | -0.205 | 3.297 | 3.405 | 0.041 | blue |
| ENSG00000184588 | PDE4B | phosphodiesterase 4B [Source:HGNC Symbol;Acc:HGNC:8781] | protein_coding | -0.363 | 0.044 | 0.407 | 4.575 | 7.698 | 0.001 | blue |
| ENSG00000185022 | MAFF | MAF bZIP transcription factor F [Source:HGNC Symbol;Acc:HGNC:6780] | protein_coding | -0.378 | 0.535 | 0.913 | 2.439 | 3.421 | 0.040 | blue |
| ENSG00000186056 | MATN1-AS1 | MATN1 antisense RNA 1 [Source:HGNC Symbol;Acc:HGNC:40364] | lncRNA | 0.519 | -0.749 | -1.269 | -1.509 | 4.168 | 0.021 | blue |
| ENSG00000186407 | CD300E | CD300e molecule [Source:HGNC Symbol;Acc:HGNC:28874] | protein_coding | 0.095 | 1.070 | 0.975 | -0.810 | 5.256 | 0.008 | blue |
| ENSG00000187140 | FOXD3 | forkhead box D3 [Source:HGNC Symbol;Acc:HGNC:3804] | protein_coding | 0.345 | -0.251 | -0.597 | -0.484 | 3.331 | 0.043 | blue |
| ENSG00000187624 | C17orf97 | chromosome 17 open reading frame 97 [Source:HGNC Symbol;Acc:HGNC:33800] | protein_coding | 0.659 | -0.078 | -0.737 | 0.250 | 3.295 | 0.045 | blue |
| ENSG00000196189 | SEMA4A | semaphorin 4A [Source:HGNC Symbol;Acc:HGNC:10729] | protein_coding | -0.780 | 0.596 | 1.377 | 1.266 | 4.446 | 0.016 | blue |
| ENSG00000196843 | ARID5A | AT-rich interaction domain 5A [Source:HGNC Symbol;Acc:HGNC:17361] | protein_coding | -0.103 | 0.435 | 0.537 | 2.619 | 4.311 | 0.018 | blue |
| ENSG00000197063 | MAFG | MAF bZIP transcription factor G [Source:HGNC Symbol;Acc:HGNC:6781] | protein_coding | 0.006 | 0.262 | 0.256 | 3.370 | 3.276 | 0.045 | blue |
| ENSG00000203392 | AC105020.1 | novel transcript, antisense to CSPG4 | lncRNA | -0.667 | -0.224 | 0.443 | -2.638 | 3.552 | 0.036 | blue |
| ENSG00000205362 | MT1A | metallothionein 1A [Source:HGNC Symbol;Acc:HGNC:7393] | protein_coding | -0.536 | 1.389 | 1.925 | -1.896 | 7.349 | 0.002 | blue |

| | | | | | | | | | | |
|---|---|---|---|---|---|---|---|---|---|---|
| ENSG00000205502 | C2CD4B | C2 calcium dependent domain containing 4B [Source:HGNC Symbol;Acc:HGNC:33628] | protein_coding | -0.145 | 1.343 | 1.488 | -1.386 | 4.024 | 0.024 | blue |
| ENSG00000205710 | C17orf107 | chromosome 17 open reading frame 107 [Source:HGNC Symbol;Acc:HGNC:37238] | protein_coding | -0.507 | 0.031 | 0.539 | 0.243 | 3.571 | 0.035 | blue |
| ENSG00000213753 | CENPBD1P1 | CENPB DNA-binding domains containing 1 pseudogene 1 [Source:HGNC Symbol;Acc:HGNC:28421] | transcribed_processed_pseudogene | -0.063 | 0.476 | 0.539 | 3.360 | 4.367 | 0.017 | blue |
| ENSG00000215022 | AL008729.1 | novel transcript, antisense to PHACTR1 | lncRNA | -1.104 | -0.510 | 0.594 | -2.011 | 4.738 | 0.013 | blue |
| ENSG00000218336 | TENM3 | teneurin transmembrane protein 3 [Source:HGNC Symbol;Acc:HGNC:29944] | protein_coding | 0.206 | -0.310 | -0.516 | 3.341 | 4.249 | 0.019 | blue |
| ENSG00000229828 | PDE4DIPP1 | phosphodiesterase 4D interacting protein pseudogene 1 [Source:HGNC Symbol;Acc:HGNC:50867] | unprocessed_pseudogene | 1.137 | -1.068 | -2.205 | -3.373 | 3.685 | 0.032 | blue |
| ENSG00000229927 | RHEBP1 | RHEB pseudogene 1 [Source:HGNC Symbol;Acc:HGNC:10010] | processed_pseudogene | 1.055 | -0.912 | -1.967 | -2.919 | 3.470 | 0.038 | blue |
| ENSG00000230658 | KLHL7-DT | KLHL7 divergent transcript [Source:HGNC Symbol;Acc:HGNC:43431] | lncRNA | -1.168 | -0.020 | 1.148 | -1.819 | 6.573 | 0.003 | blue |
| ENSG00000237513 | AC007384.1 | novel transcript | lncRNA | -0.207 | 0.783 | 0.990 | -1.425 | 3.677 | 0.032 | blue |
| ENSG00000237928 | NFIA-AS2 | NFIA antisense RNA 2 [Source:HGNC Symbol;Acc:HGNC:40401] | lncRNA | -0.712 | -0.599 | 0.114 | -0.666 | 3.829 | 0.028 | blue |
| ENSG00000239713 | APOBEC3G | apolipoprotein B mRNA editing enzyme catalytic subunit 3G [Source:HGNC Symbol;Acc:HGNC:17357] | protein_coding | 0.310 | 0.117 | -0.192 | 1.534 | 4.143 | 0.021 | blue |
| ENSG00000249839 | AC011330.1 | histidine acid phosphatase domain containing 2A (HISPPD2A) pseudogene | unprocessed_pseudogene | 2.365 | 0.372 | -1.993 | -2.799 | 5.169 | 0.009 | blue |
| ENSG00000250312 | ZNF718 | zinc finger protein 718 [Source:HGNC Symbol;Acc:HGNC:26889] | protein_coding | 0.171 | -0.399 | -0.571 | 1.365 | 3.867 | 0.027 | blue |
| ENSG00000250506 | CDK3 | cyclin dependent kinase 3 [Source:HGNC Symbol;Acc:HGNC:1772] | protein_coding | -0.909 | 0.250 | 1.160 | -1.755 | 3.429 | 0.040 | blue |
| ENSG00000251441 | RTEL1P1 | regulator of telomere elongation helicase 1 pseudogene 1 [Source:HGNC Symbol;Acc:HGNC:44213] | transcribed_processed_pseudogene | 0.660 | -0.368 | -1.028 | -2.497 | 3.763 | 0.030 | blue |
| ENSG00000255717 | SNHG1 | small nucleolar RNA host gene 1 [Source:HGNC Symbol;Acc:HGNC:32688] | lncRNA | 0.076 | -0.339 | -0.415 | 1.948 | 4.936 | 0.011 | blue |
| ENSG00000255819 | KLRC4-KLRK1 | KLRC4-KLRK1 readthrough [Source:HGNC Symbol;Acc:HGNC:48357] | protein_coding | 1.177 | -0.077 | -1.253 | -2.951 | 3.482 | 0.038 | blue |
| ENSG00000256463 | SALL3 | spalt like transcription factor 3 [Source:HGNC Symbol;Acc:HGNC:10527] | protein_coding | 0.102 | 0.880 | 0.778 | -0.631 | 3.320 | 0.044 | blue |
| ENSG00000272446 | AL158850.1 | novel transcript | lncRNA | 0.476 | -0.085 | -0.561 | -0.609 | 4.162 | 0.021 | blue |
| ENSG00000272617 | AC026464.6 | novel protein, COG8-PDF readthrough | protein_coding | 1.371 | -0.809 | -2.180 | -3.284 | 6.696 | 0.003 | blue |
| ENSG00000277283 | AC004812.2 | novel transcript, antisense to RAB35 | lncRNA | 0.078 | -0.631 | -0.709 | -0.690 | 4.947 | 0.011 | blue |
| ENSG00000277363 | SRCIN1 | SRC kinase signaling inhibitor 1 [Source:HGNC Symbol;Acc:HGNC:29506] | protein_coding | -0.279 | 0.435 | 0.714 | 0.321 | 3.882 | 0.027 | blue |

| | | | | | | | | | | |
|---|---|---|---|---|---|---|---|---|---|---|
| ENSG00000278989 | AP001148.1 | novel transcript | TEC | 0.314 | -0.506 | -0.820 | -1.567 | 3.457 | 0.039 | blue |
| ENSG00000280077 | AL353763.2 | TEC | TEC | -0.094 | -0.527 | -0.433 | -0.016 | 3.364 | 0.042 | blue |
| ENSG00000280594 | BTG3-AS1 | BTG3 antisense RNA 1 [Source:HGNC Symbol;Acc:HGNC:53145] | lncRNA | -0.354 | 0.847 | 1.201 | -2.447 | 6.519 | 0.003 | blue |
| ENSG00000285679 | AC079142.1 | novel transcript | lncRNA | -0.088 | -0.559 | -0.470 | -1.239 | 3.201 | 0.049 | blue |
| ENSG00000286235 | AL035461.4 | novel protein | protein_coding | 1.801 | -0.874 | -2.676 | -3.893 | 4.982 | 0.010 | blue |
| ENSG00000288253 | AC010332.3 | novel transcript | lncRNA | 0.691 | -0.918 | -1.610 | -2.907 | 3.933 | 0.025 | blue |
| ENSG00000288534 | AP001931.2 | TMX2-CTNND1 readthrough (NMD candidate) | protein_coding | -1.012 | -0.084 | 0.929 | 3.431 | 3.623 | 0.033 | blue |
| ENSG00000019549 | SNAI2 | snail family transcriptional repressor 2 [Source:HGNC Symbol;Acc:HGNC:11094] | protein_coding | -0.149 | -0.417 | -0.268 | 2.793 | 3.550 | 0.036 | brown |
| ENSG00000036257 | CUL3 | cullin 3 [Source:HGNC Symbol;Acc:HGNC:2553] | protein_coding | 0.237 | 0.158 | -0.079 | 4.597 | 5.665 | 0.006 | brown |
| ENSG00000049449 | RCN1 | reticulocalbin 1 [Source:HGNC Symbol;Acc:HGNC:9934] | protein_coding | -0.268 | -0.486 | -0.218 | 3.853 | 5.442 | 0.007 | brown |
| ENSG00000070526 | ST6GALNAC1 | ST6 N-acetylgalactosaminide alpha-2,6-sialyltransferase 1 [Source:HGNC Symbol;Acc:HGNC:23614] | protein_coding | 0.355 | 0.755 | 0.399 | 0.172 | 4.040 | 0.023 | brown |
| ENSG00000100664 | EIF5 | eukaryotic translation initiation factor 5 [Source:HGNC Symbol;Acc:HGNC:3299] | protein_coding | 0.202 | 0.157 | -0.045 | 6.467 | 6.477 | 0.003 | brown |
| ENSG00000101191 | DIDO1 | death inducer-obliterator 1 [Source:HGNC Symbol;Acc:HGNC:2680] | protein_coding | 0.095 | 0.136 | 0.042 | 4.853 | 4.001 | 0.024 | brown |
| ENSG00000101413 | RPRD1B | regulation of nuclear pre-mRNA domain containing 1B [Source:HGNC Symbol;Acc:HGNC:16209] | protein_coding | 0.210 | 0.374 | 0.164 | 4.496 | 4.164 | 0.021 | brown |
| ENSG00000101938 | CHRDL1 | chordin like 1 [Source:HGNC Symbol;Acc:HGNC:29861] | protein_coding | -0.271 | -0.525 | -0.255 | 4.534 | 3.324 | 0.044 | brown |
| ENSG00000104313 | EYA1 | EYA transcriptional coactivator and phosphatase 1 [Source:HGNC Symbol;Acc:HGNC:3519] | protein_coding | 0.467 | 0.743 | 0.276 | 2.425 | 5.518 | 0.007 | brown |
| ENSG00000105499 | PLA2G4C | phospholipase A2 group IVC [Source:HGNC Symbol;Acc:HGNC:9037] | protein_coding | 0.413 | 0.493 | 0.080 | 3.074 | 4.491 | 0.016 | brown |
| ENSG00000108239 | TBC1D12 | TBC1 domain family member 12 [Source:HGNC Symbol;Acc:HGNC:29082] | protein_coding | -0.220 | -0.204 | 0.017 | 3.026 | 3.612 | 0.034 | brown |
| ENSG00000110881 | ASIC1 | acid sensing ion channel subunit 1 [Source:HGNC Symbol;Acc:HGNC:100] | protein_coding | 0.008 | -0.582 | -0.589 | -0.140 | 3.196 | 0.049 | brown |
| ENSG00000111224 | PARP11 | poly(ADP-ribose) polymerase family member 11 [Source:HGNC Symbol;Acc:HGNC:1186] | protein_coding | -0.503 | -0.422 | 0.080 | 1.232 | 3.292 | 0.045 | brown |
| ENSG00000116774 | OLFML3 | olfactomedin like 3 [Source:HGNC Symbol;Acc:HGNC:24956] | protein_coding | -0.343 | -0.463 | -0.121 | 4.225 | 3.705 | 0.031 | brown |
| ENSG00000117020 | AKT3 | AKT serine/threonine kinase 3 [Source:HGNC Symbol;Acc:HGNC:393] | protein_coding | -0.222 | -0.167 | 0.055 | 5.009 | 4.142 | 0.021 | brown |

| | | | | | | | | | | |
|---|---|---|---|---|---|---|---|---|---|---|
| ENSG00000117868 | ESYT2 | extended synaptotagmin 2 [Source:HGNC Symbol;Acc:HGNC:22211] | protein_coding | -0.178 | -0.130 | 0.048 | 5.883 | 4.313 | 0.018 | brown |
| ENSG00000118194 | TNNT2 | troponin T2, cardiac type [Source:HGNC Symbol;Acc:HGNC:11949] | protein_coding | 0.126 | 0.378 | 0.253 | 10.245 | 4.575 | 0.015 | brown |
| ENSG00000119682 | AREL1 | apoptosis resistant E3 ubiquitin protein ligase 1 [Source:HGNC Symbol;Acc:HGNC:20363] | protein_coding | 0.267 | 0.462 | 0.195 | 3.489 | 4.362 | 0.018 | brown |
| ENSG00000120820 | GLT8D2 | glycosyltransferase 8 domain containing 2 [Source:HGNC Symbol;Acc:HGNC:24890] | protein_coding | -0.397 | -0.417 | -0.020 | 2.425 | 4.313 | 0.018 | brown |
| ENSG00000120910 | PPP3CC | protein phosphatase 3 catalytic subunit gamma [Source:HGNC Symbol;Acc:HGNC:9316] | protein_coding | -0.086 | 0.223 | 0.309 | 3.423 | 3.409 | 0.040 | brown |
| ENSG00000122707 | RECK | reversion inducing cysteine rich protein with kazal motifs [Source:HGNC Symbol;Acc:HGNC:11345] | protein_coding | -0.258 | -0.252 | 0.006 | 3.082 | 3.271 | 0.046 | brown |
| ENSG00000122870 | BICC1 | BicC family RNA binding protein 1 [Source:HGNC Symbol;Acc:HGNC:19351] | protein_coding | -0.286 | -0.414 | -0.128 | 4.523 | 4.033 | 0.023 | brown |
| ENSG00000129226 | CD68 | CD68 molecule [Source:HGNC Symbol;Acc:HGNC:1693] | protein_coding | -0.299 | -0.586 | -0.287 | 4.540 | 4.210 | 0.020 | brown |
| ENSG00000132780 | NASP | nuclear autoantigenic sperm protein [Source:HGNC Symbol;Acc:HGNC:7644] | protein_coding | 0.167 | 0.251 | 0.083 | 3.843 | 3.803 | 0.029 | brown |
| ENSG00000134917 | ADAMTS8 | ADAM metallopeptidase with thrombospondin type 1 motif 8 [Source:HGNC Symbol;Acc:HGNC:224] | protein_coding | -0.660 | -0.890 | -0.230 | -0.940 | 5.345 | 0.008 | brown |
| ENSG00000136999 | CCN3 | cellular communication network factor 3 [Source:HGNC Symbol;Acc:HGNC:7885] | protein_coding | -0.421 | -0.178 | 0.243 | 2.767 | 3.434 | 0.039 | brown |
| ENSG00000140092 | FBLN5 | fibulin 5 [Source:HGNC Symbol;Acc:HGNC:3602] | protein_coding | -0.261 | -0.393 | -0.132 | 5.737 | 3.873 | 0.027 | brown |
| ENSG00000140254 | DUOXA1 | dual oxidase maturation factor 1 [Source:HGNC Symbol;Acc:HGNC:26507] | protein_coding | -0.532 | -1.044 | -0.512 | -1.056 | 3.827 | 0.028 | brown |
| ENSG00000141232 | TOB1 | transducer of ERBB2, 1 [Source:HGNC Symbol;Acc:HGNC:11979] | protein_coding | -0.228 | -0.251 | -0.023 | 3.660 | 3.573 | 0.035 | brown |
| ENSG00000141252 | VPS53 | VPS53 subunit of GARP complex [Source:HGNC Symbol;Acc:HGNC:25608] | protein_coding | 0.012 | 0.353 | 0.341 | 3.760 | 4.482 | 0.016 | brown |
| ENSG00000143344 | RGL1 | ral guanine nucleotide dissociation stimulator like 1 [Source:HGNC Symbol;Acc:HGNC:30281] | protein_coding | -0.096 | -0.256 | -0.160 | 4.624 | 3.322 | 0.044 | brown |
| ENSG00000145147 | SLIT2 | slit guidance ligand 2 [Source:HGNC Symbol;Acc:HGNC:11086] | protein_coding | 0.072 | -0.267 | -0.339 | 3.931 | 5.314 | 0.008 | brown |
| ENSG00000147255 | IGSF1 | immunoglobulin superfamily member 1 [Source:HGNC Symbol;Acc:HGNC:5948] | protein_coding | 0.710 | 0.785 | 0.075 | 3.638 | 3.518 | 0.037 | brown |
| ENSG00000147394 | ZNF185 | zinc finger protein 185 with LIM domain [Source:HGNC Symbol;Acc:HGNC:12976] | protein_coding | -0.482 | -0.556 | -0.073 | 0.709 | 3.963 | 0.025 | brown |
| ENSG00000148082 | SHC3 | SHC adaptor protein 3 [Source:HGNC Symbol;Acc:HGNC:18181] | protein_coding | -0.794 | -0.579 | 0.215 | 1.601 | 5.491 | 0.007 | brown |
| ENSG00000148344 | PTGES | prostaglandin E synthase [Source:HGNC Symbol;Acc:HGNC:9599] | protein_coding | -0.578 | -0.782 | -0.204 | 0.217 | 3.931 | 0.025 | brown |
| ENSG00000148700 | ADD3 | adducin 3 [Source:HGNC Symbol;Acc:HGNC:245] | protein_coding | -0.160 | -0.228 | -0.069 | 6.483 | 3.947 | 0.025 | brown |

| | | | | | | | | | | |
|---|---|---|---|---|---|---|---|---|---|---|
| ENSG00000149090 | PAMR1 | peptidase domain containing associated with muscle regeneration 1 [Source:HGNC Symbol;Acc:HGNC:24554] | protein_coding | -0.812 | -0.817 | -0.005 | 3.052 | 6.267 | 0.004 | brown |
| ENSG00000153707 | PTPRD | protein tyrosine phosphatase receptor type D [Source:HGNC Symbol;Acc:HGNC:9668] | protein_coding | -0.710 | -0.574 | 0.136 | 2.598 | 3.432 | 0.040 | brown |
| ENSG00000154263 | ABCA10 | ATP binding cassette subfamily A member 10 [Source:HGNC Symbol;Acc:HGNC:30] | protein_coding | 0.080 | -0.466 | -0.545 | 2.739 | 3.272 | 0.046 | brown |
| ENSG00000156097 | GPR61 | G protein-coupled receptor 61 [Source:HGNC Symbol;Acc:HGNC:13300] | protein_coding | 1.073 | 0.014 | -1.059 | -2.462 | 3.250 | 0.046 | brown |
| ENSG00000156466 | GDF6 | growth differentiation factor 6 [Source:HGNC Symbol;Acc:HGNC:4221] | protein_coding | -0.449 | -0.703 | -0.254 | 1.077 | 4.000 | 0.024 | brown |
| ENSG00000162407 | PLPP3 | phospholipid phosphatase 3 [Source:HGNC Symbol;Acc:HGNC:9229] | protein_coding | -0.227 | -0.257 | -0.029 | 5.571 | 3.784 | 0.029 | brown |
| ENSG00000162616 | DNAJB4 | DnaJ heat shock protein family (Hsp40) member B4 [Source:HGNC Symbol;Acc:HGNC:14886] | protein_coding | 0.045 | 0.224 | 0.179 | 5.020 | 3.171 | 0.050 | brown |
| ENSG00000162733 | DDR2 | discoidin domain receptor tyrosine kinase 2 [Source:HGNC Symbol;Acc:HGNC:2731] | protein_coding | -0.091 | -0.209 | -0.118 | 6.606 | 4.449 | 0.016 | brown |
| ENSG00000164591 | MYOZ3 | myozenin 3 [Source:HGNC Symbol;Acc:HGNC:18565] | protein_coding | 0.472 | 0.616 | 0.144 | 1.180 | 3.789 | 0.029 | brown |
| ENSG00000165072 | MAMDC2 | MAM domain containing 2 [Source:HGNC Symbol;Acc:HGNC:23673] | protein_coding | -0.310 | -0.152 | 0.158 | 3.638 | 3.734 | 0.030 | brown |
| ENSG00000166145 | SPINT1 | serine peptidase inhibitor, Kunitz type 1 [Source:HGNC Symbol;Acc:HGNC:11246] | protein_coding | -0.856 | -1.288 | -0.432 | -0.811 | 3.971 | 0.025 | brown |
| ENSG00000166507 | NDST2 | N-deacetylase and N-sulfotransferase 2 [Source:HGNC Symbol;Acc:HGNC:7681] | protein_coding | -0.144 | -0.462 | -0.318 | 2.006 | 4.399 | 0.017 | brown |
| ENSG00000166925 | TSC22D4 | TSC22 domain family member 4 [Source:HGNC Symbol;Acc:HGNC:21696] | protein_coding | 0.243 | 0.118 | -0.125 | 3.740 | 4.501 | 0.016 | brown |
| ENSG00000166949 | SMAD3 | SMAD family member 3 [Source:HGNC Symbol;Acc:HGNC:6769] | protein_coding | -0.166 | -0.377 | -0.211 | 4.335 | 4.658 | 0.014 | brown |
| ENSG00000169604 | ANTXR1 | ANTXR cell adhesion molecule 1 [Source:HGNC Symbol;Acc:HGNC:21014] | protein_coding | -0.352 | -0.408 | -0.057 | 5.886 | 6.827 | 0.002 | brown |
| ENSG00000174348 | PODN | podocan [Source:HGNC Symbol;Acc:HGNC:23174] | protein_coding | -0.205 | -0.460 | -0.255 | 4.579 | 6.754 | 0.002 | brown |
| ENSG00000175198 | PCCA | propionyl-CoA carboxylase subunit alpha [Source:HGNC Symbol;Acc:HGNC:8653] | protein_coding | -0.298 | -0.226 | 0.072 | 3.637 | 3.840 | 0.028 | brown |
| ENSG00000176209 | SMIM19 | small integral membrane protein 19 [Source:HGNC Symbol;Acc:HGNC:25166] | protein_coding | -0.067 | 0.198 | 0.265 | 3.241 | 3.990 | 0.024 | brown |
| ENSG00000178028 | DMAP1 | DNA methyltransferase 1 associated protein 1 [Source:HGNC Symbol;Acc:HGNC:18291] | protein_coding | 0.323 | 0.182 | -0.141 | 2.342 | 4.047 | 0.023 | brown |
| ENSG00000180530 | NRIP1 | nuclear receptor interacting protein 1 [Source:HGNC Symbol;Acc:HGNC:8001] | protein_coding | -0.162 | -0.290 | -0.128 | 4.880 | 4.431 | 0.017 | brown |
| ENSG00000183117 | CSMD1 | CUB and Sushi multiple domains 1 [Source:HGNC Symbol;Acc:HGNC:14026] | protein_coding | 1.358 | 3.326 | 1.968 | -2.131 | 6.799 | 0.002 | brown |
| ENSG00000184060 | ADAP2 | ArfGAP with dual PH domains 2 [Source:HGNC Symbol;Acc:HGNC:16487] | protein_coding | -0.273 | -0.534 | -0.262 | 2.402 | 3.471 | 0.038 | brown |

| | | | | | | | | | | |
|---|---|---|---|---|---|---|---|---|---|---|
| ENSG00000186818 | LILRB4 | leukocyte immunoglobulin like receptor B4 [Source:HGNC Symbol;Acc:HGNC:6608] | protein_coding | -0.387 | -0.579 | -0.192 | 0.953 | 3.549 | 0.036 | brown |
| ENSG00000187210 | GCNT1 | glucosaminyl (N-acetyl) transferase 1 [Source:HGNC Symbol;Acc:HGNC:4203] | protein_coding | 0.500 | -0.476 | -0.976 | 1.811 | 3.623 | 0.033 | brown |
| ENSG00000189046 | ALKBH2 | alkB homolog 2, alpha-ketoglutarate dependent dioxygenase [Source:HGNC Symbol;Acc:HGNC:32487] | protein_coding | 0.176 | 0.427 | 0.250 | 0.965 | 3.254 | 0.046 | brown |
| ENSG00000196368 | NUDT11 | nudix hydrolase 11 [Source:HGNC Symbol;Acc:HGNC:18011] | protein_coding | -0.758 | -0.844 | -0.085 | -2.243 | 3.170 | 0.050 | brown |
| ENSG00000197021 | EOLA2 | endothelium and lymphocyte associated ASCH domain 2 [Source:HGNC Symbol;Acc:HGNC:17402] | protein_coding | 0.163 | 0.403 | 0.240 | 1.762 | 3.938 | 0.025 | brown |
| ENSG00000197766 | CFD | complement factor D [Source:HGNC Symbol;Acc:HGNC:2771] | protein_coding | -0.182 | -0.679 | -0.497 | 5.791 | 4.183 | 0.020 | brown |
| ENSG00000197863 | ZNF790 | zinc finger protein 790 [Source:HGNC Symbol;Acc:HGNC:33114] | protein_coding | 0.179 | 0.472 | 0.293 | 2.315 | 4.148 | 0.021 | brown |
| ENSG00000198121 | LPAR1 | lysophosphatidic acid receptor 1 [Source:HGNC Symbol;Acc:HGNC:3166] | protein_coding | -0.278 | -0.328 | -0.050 | 4.301 | 3.732 | 0.030 | brown |
| ENSG00000198744 | MTCO3P12 | MT-CO3 pseudogene 12 [Source:HGNC Symbol;Acc:HGNC:52042] | unprocessed_pseudogene | 0.451 | 2.302 | 1.850 | 1.088 | 6.646 | 0.003 | brown |
| ENSG00000198863 | RUNDC1 | RUN domain containing 1 [Source:HGNC Symbol;Acc:HGNC:25418] | protein_coding | 0.174 | 0.305 | 0.130 | 2.210 | 3.819 | 0.028 | brown |
| ENSG00000205037 | AC134312.1 | novel transcript | lncRNA | 0.355 | 1.566 | 1.211 | -2.261 | 4.727 | 0.013 | brown |
| ENSG00000205336 | ADGRG1 | adhesion G protein-coupled receptor G1 [Source:HGNC Symbol;Acc:HGNC:4512] | protein_coding | 0.720 | 0.657 | -0.063 | 1.657 | 5.916 | 0.005 | brown |
| ENSG00000224043 | CCNT2-AS1 | CCNT2 antisense RNA 1 [Source:HGNC Symbol;Acc:HGNC:40130] | lncRNA | 0.739 | 0.782 | 0.042 | 0.912 | 7.934 | 0.001 | brown |
| ENSG00000235655 | H3P6 | H3 histone pseudogene 6 [Source:HGNC Symbol;Acc:HGNC:42980] | processed_pseudogene | 0.112 | 0.591 | 0.480 | -1.586 | 5.247 | 0.008 | brown |
| ENSG00000235823 | OLMALINC | oligodendrocyte maturation-associated long intergenic non-coding RNA [Source:HGNC Symbol;Acc:HGNC:28060] | lncRNA | 0.323 | 0.159 | -0.164 | 1.695 | 3.276 | 0.045 | brown |
| ENSG00000236824 | BCYRN1 | brain cytoplasmic RNA 1 [Source:HGNC Symbol;Acc:HGNC:1022] | scRNA | 0.427 | 0.303 | -0.124 | 3.654 | 3.308 | 0.044 | brown |
| ENSG00000237596 | AL138828.1 | novel transcript | lncRNA | 0.320 | 0.949 | 0.629 | 1.742 | 4.358 | 0.018 | brown |
| ENSG00000245812 | LINC02202 | long intergenic non-protein coding RNA 2202 [Source:HGNC Symbol;Acc:HGNC:53068] | lncRNA | 0.367 | -0.608 | -0.975 | -0.475 | 4.117 | 0.022 | brown |
| ENSG00000248333 | CDK11B | cyclin dependent kinase 11B [Source:HGNC Symbol;Acc:HGNC:1729] | protein_coding | 0.268 | 0.292 | 0.023 | 2.522 | 5.361 | 0.008 | brown |
| ENSG00000253710 | ALG11 | ALG11 alpha-1,2-mannosyltransferase [Source:HGNC Symbol;Acc:HGNC:32456] | protein_coding | 0.243 | 0.146 | -0.097 | 4.173 | 4.942 | 0.011 | brown |
| ENSG00000258636 | AL121821.2 | novel transcript | lncRNA | -0.696 | -0.409 | 0.288 | -0.518 | 3.281 | 0.045 | brown |
| ENSG00000258818 | RNASE4 | ribonuclease A family member 4 [Source:HGNC Symbol;Acc:HGNC:10047] | protein_coding | -0.281 | -0.446 | -0.165 | 3.339 | 3.580 | 0.035 | brown |

| | | | | | | | | | | |
|---|---|---|---|---|---|---|---|---|---|---|
| ENSG00000267069 | AP005264.1 | novel transcript | lncRNA | 0.646 | 0.898 | 0.252 | -1.013 | 4.787 | 0.012 | brown |
| ENSG00000267530 | LINC01836 | long intergenic non-protein coding RNA 1836 [Source:HGNC Symbol;Acc:HGNC:52652] | lncRNA | 0.831 | 1.092 | 0.261 | -2.776 | 3.318 | 0.044 | brown |
| ENSG00000269896 | AL513477.1 | small nuclear ribonucleoprotein N (SNRPN) pseudogene | transcribed_processed_pseudogene | 0.114 | -1.026 | -1.140 | -2.327 | 4.656 | 0.014 | brown |
| ENSG00000275835 | TUBGCP5 | tubulin gamma complex associated protein 5 [Source:HGNC Symbol;Acc:HGNC:18600] | protein_coding | 0.353 | 0.178 | -0.176 | 2.950 | 4.410 | 0.017 | brown |
| ENSG00000279030 | AC007336.3 | novel transcript | TEC | -0.764 | -0.685 | 0.079 | -2.255 | 3.764 | 0.029 | brown |
| ENSG00000284649 | AC009093.8 | BTG3 associated nuclear protein (BANP) pseudogene | transcribed_unprocessed_pseudogene | 0.237 | 1.730 | 1.494 | -2.906 | 5.166 | 0.009 | brown |
| ENSG00000010319 | SEMA3G | semaphorin 3G [Source:HGNC Symbol;Acc:HGNC:30400] | protein_coding | 0.510 | 0.578 | 0.067 | 2.984 | 3.657 | 0.032 | green |
| ENSG00000067208 | EVI5 | ecotropic viral integration site 5 [Source:HGNC Symbol;Acc:HGNC:3501] | protein_coding | -0.251 | -0.104 | 0.148 | 4.886 | 3.865 | 0.027 | green |
| ENSG00000074582 | BCS1L | BCS1 homolog, ubiquinol-cytochrome c reductase complex chaperone [Source:HGNC Symbol;Acc:HGNC:1020] | protein_coding | 0.281 | 0.165 | -0.116 | 2.151 | 3.212 | 0.048 | green |
| ENSG00000102924 | CBLN1 | cerebellin 1 precursor [Source:HGNC Symbol;Acc:HGNC:1543] | protein_coding | 0.939 | 0.796 | -0.143 | -0.031 | 5.556 | 0.006 | green |
| ENSG00000113361 | CDH6 | cadherin 6 [Source:HGNC Symbol;Acc:HGNC:1765] | protein_coding | 0.563 | 0.792 | 0.229 | 2.100 | 4.809 | 0.012 | green |
| ENSG00000124785 | NRN1 | neuritin 1 [Source:HGNC Symbol;Acc:HGNC:17972] | protein_coding | 0.392 | 0.308 | -0.084 | 2.151 | 3.896 | 0.026 | green |
| ENSG00000125851 | PCSK2 | proprotein convertase subtilisin/kexin type 2 [Source:HGNC Symbol;Acc:HGNC:8744] | protein_coding | 0.720 | 0.163 | -0.558 | 1.669 | 6.609 | 0.003 | green |
| ENSG00000125871 | MGME1 | mitochondrial genome maintenance exonuclease 1 [Source:HGNC Symbol;Acc:HGNC:16205] | protein_coding | 0.309 | 0.399 | 0.090 | 1.779 | 3.214 | 0.048 | green |
| ENSG00000134460 | IL2RA | interleukin 2 receptor subunit alpha [Source:HGNC Symbol;Acc:HGNC:6008] | protein_coding | -1.035 | -0.571 | 0.464 | -0.255 | 4.196 | 0.020 | green |
| ENSG00000142748 | FCN3 | ficolin 3 [Source:HGNC Symbol;Acc:HGNC:3625] | protein_coding | 1.025 | 0.696 | -0.329 | -1.667 | 4.038 | 0.023 | green |
| ENSG00000143248 | RGS5 | regulator of G protein signaling 5 [Source:HGNC Symbol;Acc:HGNC:10001] | protein_coding | 0.467 | 0.313 | -0.154 | 7.130 | 5.585 | 0.006 | green |
| ENSG00000144057 | ST6GAL2 | ST6 beta-galactoside alpha-2,6-sialyltransferase 2 [Source:HGNC Symbol;Acc:HGNC:10861] | protein_coding | 0.759 | -0.586 | -1.346 | 0.271 | 6.993 | 0.002 | green |
| ENSG00000165810 | BTNL9 | butyrophilin like 9 [Source:HGNC Symbol;Acc:HGNC:24176] | protein_coding | 0.654 | 0.657 | 0.003 | 2.582 | 3.258 | 0.046 | green |
| ENSG00000168874 | ATOH8 | atonal bHLH transcription factor 8 [Source:HGNC Symbol;Acc:HGNC:24126] | protein_coding | 0.171 | -0.024 | -0.196 | 4.236 | 3.422 | 0.040 | green |
| ENSG00000177042 | TMEM80 | transmembrane protein 80 [Source:HGNC Symbol;Acc:HGNC:27453] | protein_coding | 0.374 | 0.666 | 0.292 | 1.412 | 4.596 | 0.014 | green |
| ENSG00000183153 | GJD3 | gap junction protein delta 3 [Source:HGNC Symbol;Acc:HGNC:19147] | protein_coding | 0.517 | 0.313 | -0.204 | 0.332 | 3.644 | 0.033 | green |

| | | | | | | | | | | |
|---|---|---|---|---|---|---|---|---|---|---|
| ENSG00000198597 | ZNF536 | zinc finger protein 536 [Source:HGNC Symbol;Acc:HGNC:29025] | protein_coding | 0.450 | 0.045 | -0.405 | 1.545 | 3.998 | 0.024 | green |
| ENSG00000198712 | MT-CO2 | mitochondrially encoded cytochrome c oxidase II [Source:HGNC Symbol;Acc:HGNC:7421] | protein_coding | 0.208 | 0.309 | 0.101 | 10.360 | 3.193 | 0.049 | green |
| ENSG00000205863 | C1QTNF9B | C1q and TNF related 9B [Source:HGNC Symbol;Acc:HGNC:34072] | protein_coding | 0.112 | 1.365 | 1.253 | -3.033 | 3.837 | 0.028 | green |
| ENSG00000258819 | LINC02289 | long intergenic non-protein coding RNA 2289 [Source:HGNC Symbol;Acc:HGNC:53205] | lncRNA | 0.735 | 0.637 | -0.098 | -1.247 | 3.813 | 0.028 | green |
| ENSG00000275385 | CCL18 | C-C motif chemokine ligand 18 [Source:HGNC Symbol;Acc:HGNC:10616] | protein_coding | -1.550 | -0.627 | 0.923 | -1.864 | 3.781 | 0.029 | green |
| ENSG00000279249 | AC007614.1 | novel transcript, antisense to CBLN1 | lncRNA | 0.822 | -0.104 | -0.927 | -2.156 | 4.502 | 0.016 | green |
| ENSG00000158578 | ALAS2 | 5'-aminolevulinate synthase 2 [Source:HGNC Symbol;Acc:HGNC:397] | protein_coding | 1.281 | 1.069 | -0.212 | -0.792 | 3.639 | 0.033 | greenyellow |
| ENSG00000188536 | HBA2 | hemoglobin subunit alpha 2 [Source:HGNC Symbol;Acc:HGNC:4824] | protein_coding | 0.663 | 1.364 | 0.700 | 5.305 | 4.672 | 0.013 | greenyellow |
| ENSG00000206172 | HBA1 | hemoglobin subunit alpha 1 [Source:HGNC Symbol;Acc:HGNC:4823] | protein_coding | 0.594 | 1.228 | 0.634 | 5.314 | 3.665 | 0.032 | greenyellow |
| ENSG00000215559 | ANKRD20A11P | ankyrin repeat domain 20 family member A11, pseudogene [Source:HGNC Symbol;Acc:HGNC:42024] | transcribed_unprocessed_pseudogene | -1.534 | -0.955 | 0.578 | -2.853 | 4.308 | 0.018 | greenyellow |
| ENSG00000232573 | RPL3P4 | ribosomal protein L3 pseudogene 4 [Source:HGNC Symbol;Acc:HGNC:19805] | processed_pseudogene | 1.997 | 0.091 | -1.906 | -2.570 | 5.055 | 0.010 | greenyellow |
| ENSG00000254692 | AL136295.1 | novel protein | protein_coding | 2.016 | 2.837 | 0.821 | -2.450 | 4.636 | 0.014 | greenyellow |
| ENSG00000280571 | AC006059.2 | novel protein | protein_coding | 0.218 | -3.698 | -3.916 | 0.132 | 18.007 | 0.000 | greenyellow |
| ENSG00000054179 | ENTPD2 | ectonucleoside triphosphate diphosphohydrolase 2 [Source:HGNC Symbol;Acc:HGNC:3364] | protein_coding | 0.994 | 0.405 | -0.590 | -2.448 | 4.967 | 0.010 | grey |
| ENSG00000121060 | TRIM25 | tripartite motif containing 25 [Source:HGNC Symbol;Acc:HGNC:12932] | protein_coding | 0.306 | 0.434 | 0.128 | 3.401 | 3.250 | 0.047 | grey |
| ENSG00000154040 | CABYR | calcium binding tyrosine phosphorylation regulated [Source:HGNC Symbol;Acc:HGNC:15569] | protein_coding | -1.001 | -0.324 | 0.677 | -2.742 | 4.271 | 0.019 | grey |
| ENSG00000164308 | ERAP2 | endoplasmic reticulum aminopeptidase 2 [Source:HGNC Symbol;Acc:HGNC:29499] | protein_coding | 0.682 | -0.547 | -1.229 | 3.103 | 3.533 | 0.036 | grey |
| ENSG00000166140 | ZFYVE19 | zinc finger FYVE-type containing 19 [Source:HGNC Symbol;Acc:HGNC:20758] | protein_coding | -0.197 | 0.118 | 0.316 | 1.987 | 3.646 | 0.033 | grey |
| ENSG00000228252 | COL6A4P2 | collagen type VI alpha 4 pseudogene 2 [Source:HGNC Symbol;Acc:HGNC:38501] | transcribed_unitary_pseudogene | 0.843 | -0.214 | -1.056 | -2.876 | 3.181 | 0.049 | grey |

| | | | | | | | | | | |
|---|---|---|---|---|---|---|---|---|---|---|
| ENSG00000235535 | TRDN-AS1 | TRDN antisense RNA 1 [Source:HGNC Symbol;Acc:HGNC:40592] | lncRNA | -0.590 | 0.023 | 0.612 | 2.529 | 4.122 | 0.022 | grey |
| ENSG00000261175 | LINC02188 | long intergenic non-protein coding RNA 2188 [Source:HGNC Symbol;Acc:HGNC:53050] | lncRNA | -0.645 | 0.615 | 1.260 | -2.862 | 4.171 | 0.021 | grey |
| ENSG00000266993 | AL050343.2 | novel transcript, antisense to NRD1 | lncRNA | -0.143 | -1.296 | -1.153 | -2.139 | 4.207 | 0.020 | grey |
| ENSG00000283809 | AC007326.4 | novel protein | protein_coding | -1.155 | -0.231 | 0.924 | -1.985 | 3.292 | 0.045 | grey |
| ENSG00000110880 | CORO1C | coronin 1C [Source:HGNC Symbol;Acc:HGNC:2254] | protein_coding | -0.206 | -0.193 | 0.012 | 4.413 | 3.885 | 0.027 | magenta |
| ENSG00000113966 | ARL6 | ADP ribosylation factor like GTPase 6 [Source:HGNC Symbol;Acc:HGNC:13210] | protein_coding | -0.282 | -0.463 | -0.182 | 1.603 | 3.169 | 0.050 | magenta |
| ENSG00000142856 | ITGB3BP | integrin subunit beta 3 binding protein [Source:HGNC Symbol;Acc:HGNC:6157] | protein_coding | -0.698 | -0.577 | 0.121 | 1.007 | 6.449 | 0.003 | magenta |
| ENSG00000151322 | NPAS3 | neuronal PAS domain protein 3 [Source:HGNC Symbol;Acc:HGNC:19311] | protein_coding | -0.765 | -0.575 | 0.190 | 0.469 | 3.618 | 0.034 | magenta |
| ENSG00000159712 | ANKRD18CP | ankyrin repeat domain 18C, pseudogene [Source:HGNC Symbol;Acc:HGNC:43601] | unprocessed_pseudogene | -1.036 | -0.445 | 0.591 | -2.379 | 3.475 | 0.038 | magenta |
| ENSG00000196504 | PRPF40A | pre-mRNA processing factor 40 homolog A [Source:HGNC Symbol;Acc:HGNC:16463] | protein_coding | 0.262 | 0.143 | -0.119 | 5.011 | 5.077 | 0.010 | magenta |
| ENSG00000259001 | AL355075.4 | ribonuclease P RNA component H1 | lncRNA | -0.425 | -0.696 | -0.272 | 3.465 | 3.434 | 0.039 | magenta |
| ENSG00000268433 | MTDHP3 | metadherin pseudogene 3 [Source:HGNC Symbol;Acc:HGNC:52359] | processed_pseudogene | -1.089 | 0.149 | 1.238 | -2.379 | 6.090 | 0.004 | magenta |
| ENSG00000270757 | HSPE1-MOB4 | HSPE1-MOB4 readthrough [Source:HGNC Symbol;Acc:HGNC:49184] | protein_coding | 0.999 | 2.525 | 1.525 | -3.277 | 3.399 | 0.041 | magenta |
| ENSG00000277209 | RPPH1 | ribonuclease P RNA component H1 [Source:HGNC Symbol;Acc:HGNC:19273] | ribozyme | -0.323 | -0.626 | -0.303 | 9.457 | 4.387 | 0.017 | magenta |
| ENSG00000075213 | SEMA3A | semaphorin 3A [Source:HGNC Symbol;Acc:HGNC:10723] | protein_coding | 0.074 | 0.540 | 0.466 | 2.826 | 4.255 | 0.019 | pink |
| ENSG00000112782 | CLIC5 | chloride intracellular channel 5 [Source:HGNC Symbol;Acc:HGNC:13517] | protein_coding | 0.224 | -0.007 | -0.231 | 6.753 | 5.095 | 0.009 | pink |
| ENSG00000123405 | NFE2 | nuclear factor, erythroid 2 [Source:HGNC Symbol;Acc:HGNC:7780] | protein_coding | 0.820 | 1.177 | 0.356 | -1.642 | 3.686 | 0.032 | pink |
| ENSG00000153443 | UBALD1 | UBA like domain containing 1 [Source:HGNC Symbol;Acc:HGNC:29576] | protein_coding | -0.281 | -0.307 | -0.027 | 1.191 | 3.216 | 0.048 | pink |
| ENSG00000164116 | GUCY1A1 | guanylate cyclase 1 soluble subunit alpha 1 [Source:HGNC Symbol;Acc:HGNC:4685] | protein_coding | 0.147 | 0.331 | 0.184 | 5.868 | 3.419 | 0.040 | pink |
| ENSG00000183828 | NUDT14 | nudix hydrolase 14 [Source:HGNC Symbol;Acc:HGNC:20141] | protein_coding | 0.195 | -0.163 | -0.359 | 1.578 | 3.477 | 0.038 | pink |
| ENSG00000196781 | TLE1 | TLE family member 1, transcriptional corepressor [Source:HGNC Symbol;Acc:HGNC:11837] | protein_coding | -0.240 | 0.093 | 0.334 | 3.139 | 3.421 | 0.040 | pink |
| ENSG00000197249 | SERPINA1 | serpin family A member 1 [Source:HGNC Symbol;Acc:HGNC:8941] | protein_coding | 0.291 | 0.769 | 0.477 | 1.284 | 3.742 | 0.030 | pink |

| | | | | | | | | | | |
|---|---|---|---|---|---|---|---|---|---|---|
| ENSG00000206337 | HCP5 | HLA complex P5 [Source:HGNC Symbol;Acc:HGNC:21659] | lncRNA | 0.358 | 0.227 | -0.131 | 2.918 | 4.207 | 0.020 | pink |
| ENSG00000206503 | HLA-A | major histocompatibility complex, class I, A [Source:HGNC Symbol;Acc:HGNC:4931] | protein_coding | 0.318 | 0.180 | -0.138 | 7.089 | 4.012 | 0.024 | pink |
| ENSG00000211598 | IGKV4-1 | immunoglobulin kappa variable 4-1 [Source:HGNC Symbol;Acc:HGNC:5834] | IG_V_gene | -2.074 | -1.365 | 0.709 | -0.574 | 3.245 | 0.047 | pink |
| ENSG00000211938 | IGHV3-7 | immunoglobulin heavy variable 3-7 [Source:HGNC Symbol;Acc:HGNC:5620] | IG_V_gene | 1.385 | 2.422 | 1.037 | -1.426 | 3.262 | 0.046 | pink |
| ENSG00000254838 | GVINP1 | GTPase, very large interferon inducible pseudogene 1 [Source:HGNC Symbol;Acc:HGNC:25813] | transcribed_unprocessed_pseudogene | 0.423 | 0.246 | -0.177 | 2.093 | 4.444 | 0.016 | pink |
| ENSG00000287129 | AC097500.1 | novel transcript | lncRNA | -0.719 | 0.140 | 0.859 | -2.475 | 3.958 | 0.025 | pink |
| ENSG00000100031 | GGT1 | gamma-glutamyltransferase 1 [Source:HGNC Symbol;Acc:HGNC:4250] | protein_coding | -0.739 | -1.651 | -0.912 | -0.933 | 3.449 | 0.039 | purple |
| ENSG00000112972 | HMGCS1 | 3-hydroxy-3-methylglutaryl-CoA synthase 1 [Source:HGNC Symbol;Acc:HGNC:5007] | protein_coding | -0.034 | 0.411 | 0.445 | 3.535 | 3.296 | 0.045 | purple |
| ENSG00000121858 | TNFSF10 | TNF superfamily member 10 [Source:HGNC Symbol;Acc:HGNC:11925] | protein_coding | 0.264 | -0.060 | -0.324 | 4.139 | 4.796 | 0.012 | purple |
| ENSG00000130164 | LDLR | low density lipoprotein receptor [Source:HGNC Symbol;Acc:HGNC:6547] | protein_coding | -0.051 | 0.914 | 0.965 | 3.595 | 5.668 | 0.006 | purple |
| ENSG00000165186 | PTCHD1 | patched domain containing 1 [Source:HGNC Symbol;Acc:HGNC:26392] | protein_coding | 0.744 | 0.113 | -0.631 | -0.229 | 3.271 | 0.046 | purple |
| ENSG00000171657 | GPR82 | G protein-coupled receptor 82 [Source:HGNC Symbol;Acc:HGNC:4533] | protein_coding | 0.312 | -0.740 | -1.052 | -1.881 | 3.665 | 0.032 | purple |
| ENSG00000186480 | INSIG1 | insulin induced gene 1 [Source:HGNC Symbol;Acc:HGNC:6083] | protein_coding | -0.160 | 0.383 | 0.544 | 3.830 | 3.718 | 0.031 | purple |
| ENSG00000213523 | SRA1 | steroid receptor RNA activator 1 [Source:HGNC Symbol;Acc:HGNC:11281] | protein_coding | 0.014 | 0.254 | 0.240 | 2.751 | 3.229 | 0.047 | purple |
| ENSG00000243364 | EFNA4 | ephrin A4 [Source:HGNC Symbol;Acc:HGNC:3224] | protein_coding | 0.218 | -0.360 | -0.578 | -0.823 | 3.544 | 0.036 | purple |
| ENSG00000005801 | ZNF195 | zinc finger protein 195 [Source:HGNC Symbol;Acc:HGNC:12986] | protein_coding | -0.233 | -0.006 | 0.227 | 2.624 | 3.900 | 0.026 | red |
| ENSG00000009307 | CSDE1 | cold shock domain containing E1 [Source:HGNC Symbol;Acc:HGNC:29905] | protein_coding | -0.043 | 0.060 | 0.103 | 8.481 | 3.570 | 0.035 | red |
| ENSG00000073464 | CLCN4 | chloride voltage-gated channel 4 [Source:HGNC Symbol;Acc:HGNC:2022] | protein_coding | 0.224 | 0.293 | 0.068 | 4.630 | 4.355 | 0.018 | red |
| ENSG00000077157 | PPP1R12B | protein phosphatase 1 regulatory subunit 12B [Source:HGNC Symbol;Acc:HGNC:7619] | protein_coding | 0.062 | 0.282 | 0.220 | 9.773 | 4.332 | 0.018 | red |
| ENSG00000080166 | DCT | dopachrome tautomerase [Source:HGNC Symbol;Acc:HGNC:2709] | protein_coding | -0.202 | 1.034 | 1.236 | -1.950 | 4.340 | 0.018 | red |
| ENSG00000115138 | POMC | proopiomelanocortin [Source:HGNC Symbol;Acc:HGNC:9201] | protein_coding | 0.250 | 0.960 | 0.710 | -0.547 | 5.196 | 0.009 | red |
| ENSG00000128482 | RNF112 | ring finger protein 112 [Source:HGNC Symbol;Acc:HGNC:12968] | protein_coding | 0.337 | -0.295 | -0.633 | -0.290 | 3.368 | 0.042 | red |

| | | | | | | | | | | |
|---|---|---|---|---|---|---|---|---|---|---|
| ENSG00000132274 | TRIM22 | tripartite motif containing 22 [Source:HGNC Symbol;Acc:HGNC:16379] | protein_coding | 0.042 | -0.315 | -0.357 | 4.319 | 3.177 | 0.050 | red |
| ENSG00000135930 | EIF4E2 | eukaryotic translation initiation factor 4E family member 2 [Source:HGNC Symbol;Acc:HGNC:3293] | protein_coding | -0.204 | -0.371 | -0.166 | 3.815 | 3.348 | 0.043 | red |
| ENSG00000135931 | ARMC9 | armadillo repeat containing 9 [Source:HGNC Symbol;Acc:HGNC:20730] | protein_coding | -0.431 | -0.804 | -0.372 | 1.067 | 4.524 | 0.015 | red |
| ENSG00000174437 | ATP2A2 | ATPase sarcoplasmic/endoplasmic reticulum Ca2+ transporting 2 [Source:HGNC Symbol;Acc:HGNC:812] | protein_coding | 0.056 | 0.275 | 0.219 | 9.765 | 3.624 | 0.033 | red |
| ENSG00000183508 | TENT5C | terminal nucleotidyltransferase 5C [Source:HGNC Symbol;Acc:HGNC:24712] | protein_coding | 0.209 | 0.367 | 0.158 | 4.191 | 5.090 | 0.009 | red |
| ENSG00000189283 | FHIT | fragile histidine triad diadenosine triphosphatase [Source:HGNC Symbol;Acc:HGNC:3701] | protein_coding | 0.301 | 0.773 | 0.473 | 1.148 | 3.877 | 0.027 | red |
| ENSG00000189419 | SPATA41 | spermatogenesis associated 41 [Source:HGNC Symbol;Acc:HGNC:48613] | lncRNA | 0.132 | -1.343 | -1.475 | -1.872 | 7.503 | 0.001 | red |
| ENSG00000196196 | HRCT1 | histidine rich carboxyl terminus 1 [Source:HGNC Symbol;Acc:HGNC:33872] | protein_coding | -0.402 | -1.096 | -0.694 | -1.425 | 3.717 | 0.031 | red |
| ENSG00000213928 | IRF9 | interferon regulatory factor 9 [Source:HGNC Symbol;Acc:HGNC:6131] | protein_coding | 0.259 | -0.045 | -0.304 | 3.190 | 3.321 | 0.044 | red |
| ENSG00000226816 | AC005082.1 | novel transcript | lncRNA | -0.414 | 0.645 | 1.059 | -1.461 | 4.575 | 0.015 | red |
| ENSG00000249378 | LINC01060 | long intergenic non-protein coding RNA 1060 [Source:HGNC Symbol;Acc:HGNC:49081] | lncRNA | 0.410 | -1.306 | -1.716 | -3.382 | 3.473 | 0.038 | red |
| ENSG00000277639 | AC007906.2 | novel protein | protein_coding | 0.056 | -1.114 | -1.170 | -1.771 | 3.722 | 0.031 | red |
| ENSG00000286039 | AC093849.2 | novel transcript | lncRNA | -0.034 | 0.324 | 0.358 | 2.186 | 4.220 | 0.020 | red |
| ENSG00000003402 | CFLAR | CASP8 and FADD like apoptosis regulator [Source:HGNC Symbol;Acc:HGNC:1876] | protein_coding | 0.191 | 0.220 | 0.029 | 6.684 | 3.404 | 0.041 | turquoise |
| ENSG00000008988 | RPS20 | ribosomal protein S20 [Source:HGNC Symbol;Acc:HGNC:10405] | protein_coding | -0.159 | -0.097 | 0.062 | 7.193 | 3.215 | 0.048 | turquoise |
| ENSG00000047849 | MAP4 | microtubule associated protein 4 [Source:HGNC Symbol;Acc:HGNC:6862] | protein_coding | 0.211 | 0.098 | -0.113 | 8.418 | 3.812 | 0.028 | turquoise |
| ENSG00000068781 | STON1-GTF2A1L | STON1-GTF2A1L readthrough [Source:HGNC Symbol;Acc:HGNC:30651] | protein_coding | 0.795 | 1.183 | 0.388 | 1.178 | 3.587 | 0.034 | turquoise |
| ENSG00000076706 | MCAM | melanoma cell adhesion molecule [Source:HGNC Symbol;Acc:HGNC:6934] | protein_coding | 0.240 | 0.122 | -0.118 | 6.150 | 6.109 | 0.004 | turquoise |
| ENSG00000082684 | SEMA5B | semaphorin 5B [Source:HGNC Symbol;Acc:HGNC:10737] | protein_coding | 0.773 | 0.902 | 0.129 | 1.011 | 5.696 | 0.006 | turquoise |
| ENSG00000089157 | RPLP0 | ribosomal protein lateral stalk subunit P0 [Source:HGNC Symbol;Acc:HGNC:10371] | protein_coding | -0.183 | -0.182 | 0.001 | 7.379 | 3.232 | 0.047 | turquoise |
| ENSG00000091622 | PITPNM3 | PITPNM family member 3 [Source:HGNC Symbol;Acc:HGNC:21043] | protein_coding | 0.602 | 0.919 | 0.317 | 1.025 | 4.251 | 0.019 | turquoise |
| ENSG00000092529 | CAPN3 | calpain 3 [Source:HGNC Symbol;Acc:HGNC:1480] | protein_coding | 0.484 | 0.235 | -0.249 | 1.996 | 3.428 | 0.040 | turquoise |

| | | | | | | | | | | |
|---|---|---|---|---|---|---|---|---|---|---|
| ENSG00000095637 | SORBS1 | sorbin and SH3 domain containing 1 [Source:HGNC Symbol;Acc:HGNC:14565] | protein_coding | 0.173 | 0.075 | -0.098 | 7.955 | 3.361 | 0.042 | turquoise |
| ENSG00000099282 | TSPAN15 | tetraspanin 15 [Source:HGNC Symbol;Acc:HGNC:23298] | protein_coding | 0.448 | 0.384 | -0.064 | 1.459 | 3.883 | 0.027 | turquoise |
| ENSG00000100316 | RPL3 | ribosomal protein L3 [Source:HGNC Symbol;Acc:HGNC:10332] | protein_coding | -0.190 | -0.120 | 0.071 | 7.099 | 3.183 | 0.049 | turquoise |
| ENSG00000100836 | PABPN1 | poly(A) binding protein nuclear 1 [Source:HGNC Symbol;Acc:HGNC:8565] | protein_coding | 0.266 | 0.137 | -0.129 | 4.163 | 4.963 | 0.011 | turquoise |
| ENSG00000103245 | CIAO3 | cytosolic iron-sulfur assembly component 3 [Source:HGNC Symbol;Acc:HGNC:14179] | protein_coding | 0.393 | 0.237 | -0.156 | 2.397 | 5.280 | 0.008 | turquoise |
| ENSG00000105723 | GSK3A | glycogen synthase kinase 3 alpha [Source:HGNC Symbol;Acc:HGNC:4616] | protein_coding | 0.228 | 0.216 | -0.013 | 2.872 | 3.593 | 0.034 | turquoise |
| ENSG00000107263 | RAPGEF1 | Rap guanine nucleotide exchange factor 1 [Source:HGNC Symbol;Acc:HGNC:4568] | protein_coding | 0.204 | 0.176 | -0.028 | 4.387 | 3.348 | 0.043 | turquoise |
| ENSG00000107798 | LIPA | lipase A, lysosomal acid type [Source:HGNC Symbol;Acc:HGNC:6617] | protein_coding | -0.296 | -0.310 | -0.013 | 4.136 | 3.301 | 0.044 | turquoise |
| ENSG00000109099 | PMP22 | peripheral myelin protein 22 [Source:HGNC Symbol;Acc:HGNC:9118] | protein_coding | -0.184 | -0.196 | -0.012 | 5.509 | 4.483 | 0.016 | turquoise |
| ENSG00000110799 | VWF | von Willebrand factor [Source:HGNC Symbol;Acc:HGNC:12726] | protein_coding | 0.381 | 0.270 | -0.110 | 7.554 | 5.778 | 0.005 | turquoise |
| ENSG00000112210 | RAB23 | RAB23, member RAS oncogene family [Source:HGNC Symbol;Acc:HGNC:14263] | protein_coding | -0.206 | -0.055 | 0.151 | 3.798 | 3.472 | 0.038 | turquoise |
| ENSG00000112306 | RPS12 | ribosomal protein S12 [Source:HGNC Symbol;Acc:HGNC:10385] | protein_coding | -0.175 | -0.175 | 0.000 | 6.094 | 3.407 | 0.040 | turquoise |
| ENSG00000115145 | STAM2 | signal transducing adaptor molecule 2 [Source:HGNC Symbol;Acc:HGNC:11358] | protein_coding | -0.126 | -0.206 | -0.080 | 4.280 | 3.791 | 0.029 | turquoise |
| ENSG00000115904 | SOS1 | SOS Ras/Rac guanine nucleotide exchange factor 1 [Source:HGNC Symbol;Acc:HGNC:11187] | protein_coding | 0.304 | 0.087 | -0.217 | 4.676 | 3.315 | 0.044 | turquoise |
| ENSG00000116251 | RPL22 | ribosomal protein L22 [Source:HGNC Symbol;Acc:HGNC:10315] | protein_coding | -0.197 | -0.135 | 0.061 | 5.952 | 4.906 | 0.011 | turquoise |
| ENSG00000118454 | ANKRD13C | ankyrin repeat domain 13C [Source:HGNC Symbol;Acc:HGNC:25374] | protein_coding | -0.320 | -0.208 | 0.112 | 3.447 | 3.928 | 0.026 | turquoise |
| ENSG00000119655 | NPC2 | NPC intracellular cholesterol transporter 2 [Source:HGNC Symbol;Acc:HGNC:14537] | protein_coding | -0.220 | -0.240 | -0.020 | 4.374 | 3.235 | 0.047 | turquoise |
| ENSG00000120451 | SNX19 | sorting nexin 19 [Source:HGNC Symbol;Acc:HGNC:21532] | protein_coding | 0.243 | -0.017 | -0.260 | 5.310 | 5.058 | 0.010 | turquoise |
| ENSG00000123427 | EEF1AKMT3 | EEF1A lysine methyltransferase 3 [Source:HGNC Symbol;Acc:HGNC:24936] | protein_coding | -0.248 | -0.546 | -0.298 | 0.563 | 3.489 | 0.038 | turquoise |
| ENSG00000123636 | BAZ2B | bromodomain adjacent to zinc finger domain 2B [Source:HGNC Symbol;Acc:HGNC:963] | protein_coding | 0.203 | 0.225 | 0.022 | 5.083 | 3.306 | 0.044 | turquoise |
| ENSG00000128591 | FLNC | filamin C [Source:HGNC Symbol;Acc:HGNC:3756] | protein_coding | 0.243 | 0.147 | -0.095 | 7.905 | 3.422 | 0.040 | turquoise |
| ENSG00000128891 | CCDC32 | coiled-coil domain containing 32 [Source:HGNC Symbol;Acc:HGNC:28295] | protein_coding | 0.291 | 0.019 | -0.272 | 2.514 | 5.944 | 0.005 | turquoise |

| | | | | | | | | | | |
|---|---|---|---|---|---|---|---|---|---|---|
| ENSG00000130208 | APOC1 | apolipoprotein C1 [Source:HGNC Symbol;Acc:HGNC:607] | protein_coding | -0.929 | -0.920 | 0.009 | -1.933 | 4.031 | 0.023 | turquoise |
| ENSG00000130338 | TULP4 | TUB like protein 4 [Source:HGNC Symbol;Acc:HGNC:15530] | protein_coding | -0.135 | 0.019 | 0.154 | 6.330 | 3.307 | 0.044 | turquoise |
| ENSG00000130816 | DNMT1 | DNA methyltransferase 1 [Source:HGNC Symbol;Acc:HGNC:2976] | protein_coding | 0.540 | 0.392 | -0.148 | 3.285 | 8.849 | 0.000 | turquoise |
| ENSG00000130818 | ZNF426 | zinc finger protein 426 [Source:HGNC Symbol;Acc:HGNC:20725] | protein_coding | -0.227 | -0.085 | 0.142 | 3.917 | 3.508 | 0.037 | turquoise |
| ENSG00000134030 | CTIF | cap binding complex dependent translation initiation factor [Source:HGNC Symbol;Acc:HGNC:23925] | protein_coding | 0.303 | 0.197 | -0.106 | 4.061 | 3.707 | 0.031 | turquoise |
| ENSG00000134419 | RPS15A | ribosomal protein S15a [Source:HGNC Symbol;Acc:HGNC:10389] | protein_coding | -0.199 | -0.202 | -0.003 | 6.226 | 3.430 | 0.040 | turquoise |
| ENSG00000135837 | CEP350 | centrosomal protein 350 [Source:HGNC Symbol;Acc:HGNC:24238] | protein_coding | 0.210 | 0.190 | -0.021 | 5.693 | 3.326 | 0.043 | turquoise |
| ENSG00000137876 | RSL24D1 | ribosomal L24 domain containing 1 [Source:HGNC Symbol;Acc:HGNC:18479] | protein_coding | -0.241 | -0.165 | 0.076 | 4.318 | 3.848 | 0.027 | turquoise |
| ENSG00000138363 | ATIC | 5-aminoimidazole-4-carboxamide ribonucleotide formyltransferase/IMP cyclohydrolase [Source:HGNC Symbol;Acc:HGNC:794] | protein_coding | 0.530 | 0.241 | -0.288 | 1.500 | 3.254 | 0.046 | turquoise |
| ENSG00000138623 | SEMA7A | semaphorin 7A (John Milton Hagen blood group) [Source:HGNC Symbol;Acc:HGNC:10741] | protein_coding | 0.638 | 0.356 | -0.282 | -0.097 | 3.672 | 0.032 | turquoise |
| ENSG00000138738 | PRDM5 | PR/SET domain 5 [Source:HGNC Symbol;Acc:HGNC:9349] | protein_coding | -0.204 | 0.110 | 0.314 | 3.657 | 3.211 | 0.048 | turquoise |
| ENSG00000139714 | MORN3 | MORN repeat containing 3 [Source:HGNC Symbol;Acc:HGNC:29807] | protein_coding | -0.831 | -0.024 | 0.807 | -1.867 | 4.480 | 0.016 | turquoise |
| ENSG00000140983 | RHOT2 | ras homolog family member T2 [Source:HGNC Symbol;Acc:HGNC:21169] | protein_coding | 0.266 | 0.022 | -0.244 | 3.270 | 3.734 | 0.030 | turquoise |
| ENSG00000141668 | CBLN2 | cerebellin 2 precursor [Source:HGNC Symbol;Acc:HGNC:1544] | protein_coding | -1.858 | -1.538 | 0.320 | -2.318 | 5.888 | 0.005 | turquoise |
| ENSG00000143033 | MTF2 | metal response element binding transcription factor 2 [Source:HGNC Symbol;Acc:HGNC:29535] | protein_coding | 0.304 | 0.295 | -0.009 | 2.380 | 3.279 | 0.045 | turquoise |
| ENSG00000143162 | CREG1 | cellular repressor of E1A stimulated genes 1 [Source:HGNC Symbol;Acc:HGNC:2351] | protein_coding | -0.171 | -0.226 | -0.054 | 5.171 | 4.137 | 0.021 | turquoise |
| ENSG00000143970 | ASXL2 | ASXL transcriptional regulator 2 [Source:HGNC Symbol;Acc:HGNC:23805] | protein_coding | 0.251 | 0.096 | -0.155 | 4.723 | 4.088 | 0.022 | turquoise |
| ENSG00000146006 | LRRTM2 | leucine rich repeat transmembrane neuronal 2 [Source:HGNC Symbol;Acc:HGNC:19409] | protein_coding | 1.167 | 0.938 | -0.229 | -1.976 | 4.536 | 0.015 | turquoise |
| ENSG00000146278 | PNRC1 | proline rich nuclear receptor coactivator 1 [Source:HGNC Symbol;Acc:HGNC:17278] | protein_coding | -0.208 | -0.101 | 0.107 | 6.229 | 3.899 | 0.026 | turquoise |
| ENSG00000147419 | CCDC25 | coiled-coil domain containing 25 [Source:HGNC Symbol;Acc:HGNC:25591] | protein_coding | -0.215 | -0.146 | 0.069 | 3.726 | 3.286 | 0.045 | turquoise |
| ENSG00000148803 | FUOM | fucose mutarotase [Source:HGNC Symbol;Acc:HGNC:24733] | protein_coding | -0.359 | -0.272 | 0.087 | 0.764 | 3.396 | 0.041 | turquoise |
| ENSG00000149532 | CPSF7 | cleavage and polyadenylation specific factor 7 [Source:HGNC Symbol;Acc:HGNC:30098] | protein_coding | 0.220 | 0.102 | -0.118 | 4.057 | 3.730 | 0.030 | turquoise |

| | | | | | | | | | | |
|---|---|---|---|---|---|---|---|---|---|---|
| ENSG00000156875 | MFSD14A | major facilitator superfamily domain containing 14A [Source:HGNC Symbol;Acc:HGNC:23363] | protein_coding | -0.159 | -0.014 | 0.145 | 3.701 | 3.246 | 0.047 | turquoise |
| ENSG00000159023 | EPB41 | erythrocyte membrane protein band 4.1 [Source:HGNC Symbol;Acc:HGNC:3377] | protein_coding | 0.312 | 0.283 | -0.029 | 4.131 | 4.494 | 0.016 | turquoise |
| ENSG00000159788 | RGS12 | regulator of G protein signaling 12 [Source:HGNC Symbol;Acc:HGNC:9994] | protein_coding | 0.416 | 0.122 | -0.294 | 2.446 | 5.176 | 0.009 | turquoise |
| ENSG00000160007 | ARHGAP35 | Rho GTPase activating protein 35 [Source:HGNC Symbol;Acc:HGNC:4591] | protein_coding | 0.177 | 0.188 | 0.011 | 6.083 | 3.666 | 0.032 | turquoise |
| ENSG00000160321 | ZNF208 | zinc finger protein 208 [Source:HGNC Symbol;Acc:HGNC:12999] | protein_coding | -0.871 | -0.070 | 0.801 | 2.777 | 8.398 | 0.001 | turquoise |
| ENSG00000160408 | ST6GALNAC6 | ST6 N-acetylgalactosaminide alpha-2,6-sialyltransferase 6 [Source:HGNC Symbol;Acc:HGNC:23364] | protein_coding | -0.231 | -0.211 | 0.020 | 4.699 | 4.203 | 0.020 | turquoise |
| ENSG00000160803 | UBQLN4 | ubiquilin 4 [Source:HGNC Symbol;Acc:HGNC:1237] | protein_coding | 0.209 | 0.014 | -0.195 | 3.223 | 4.732 | 0.013 | turquoise |
| ENSG00000160908 | ZNF394 | zinc finger protein 394 [Source:HGNC Symbol;Acc:HGNC:18832] | protein_coding | -0.197 | -0.114 | 0.083 | 2.996 | 3.632 | 0.033 | turquoise |
| ENSG00000161970 | RPL26 | ribosomal protein L26 [Source:HGNC Symbol;Acc:HGNC:10327] | protein_coding | -0.153 | -0.141 | 0.013 | 7.280 | 3.221 | 0.048 | turquoise |
| ENSG00000163291 | PAQR3 | progestin and adipoQ receptor family member 3 [Source:HGNC Symbol;Acc:HGNC:30130] | protein_coding | -0.291 | -0.240 | 0.051 | 2.042 | 3.452 | 0.039 | turquoise |
| ENSG00000163655 | GMPS | guanine monophosphate synthase [Source:HGNC Symbol;Acc:HGNC:4378] | protein_coding | -0.181 | -0.049 | 0.132 | 4.148 | 3.642 | 0.033 | turquoise |
| ENSG00000164944 | VIRMA | vir like m6A methyltransferase associated [Source:HGNC Symbol;Acc:HGNC:24500] | protein_coding | 0.228 | 0.286 | 0.058 | 4.445 | 3.774 | 0.029 | turquoise |
| ENSG00000166562 | SEC11C | SEC11 homolog C, signal peptidase complex subunit [Source:HGNC Symbol;Acc:HGNC:23400] | protein_coding | -0.203 | -0.029 | 0.174 | 2.184 | 3.284 | 0.045 | turquoise |
| ENSG00000167524 | RSKR | ribosomal protein S6 kinase related [Source:HGNC Symbol;Acc:HGNC:26314] | protein_coding | 0.590 | 0.393 | -0.197 | -0.179 | 3.238 | 0.047 | turquoise |
| ENSG00000169379 | ARL13B | ADP ribosylation factor like GTPase 13B [Source:HGNC Symbol;Acc:HGNC:25419] | protein_coding | -0.317 | -0.153 | 0.164 | 2.154 | 3.302 | 0.044 | turquoise |
| ENSG00000170296 | GABARAP | GABA type A receptor-associated protein [Source:HGNC Symbol;Acc:HGNC:4067] | protein_coding | -0.121 | -0.203 | -0.082 | 5.172 | 3.280 | 0.045 | turquoise |
| ENSG00000170484 | KRT74 | keratin 74 [Source:HGNC Symbol;Acc:HGNC:28929] | protein_coding | -1.460 | -0.210 | 1.251 | -3.427 | 4.834 | 0.012 | turquoise |
| ENSG00000171132 | PRKCE | protein kinase C epsilon [Source:HGNC Symbol;Acc:HGNC:9401] | protein_coding | 0.326 | 0.332 | 0.006 | 2.746 | 3.177 | 0.050 | turquoise |
| ENSG00000171943 | SRGAP2C | SLIT-ROBO Rho GTPase activating protein 2C [Source:HGNC Symbol;Acc:HGNC:30584] | protein_coding | -0.481 | -0.436 | 0.044 | 4.035 | 4.918 | 0.011 | turquoise |
| ENSG00000172007 | RAB33B | RAB33B, member RAS oncogene family [Source:HGNC Symbol;Acc:HGNC:16075] | protein_coding | -0.167 | 0.021 | 0.188 | 3.208 | 3.567 | 0.035 | turquoise |
| ENSG00000172594 | SMPDL3A | sphingomyelin phosphodiesterase acid like 3A [Source:HGNC Symbol;Acc:HGNC:17389] | protein_coding | -0.379 | -0.377 | 0.002 | 1.412 | 6.345 | 0.003 | turquoise |
| ENSG00000174444 | RPL4 | ribosomal protein L4 [Source:HGNC Symbol;Acc:HGNC:10353] | protein_coding | -0.193 | -0.152 | 0.041 | 7.818 | 3.688 | 0.032 | turquoise |

| | | | | | | | | | | |
|---|---|---|---|---|---|---|---|---|---|---|
| ENSG00000175137 | SH3BP5L | SH3 binding domain protein 5 like [Source:HGNC Symbol;Acc:HGNC:29360] | protein_coding | 0.204 | 0.161 | -0.043 | 3.050 | 4.098 | 0.022 | turquoise |
| ENSG00000177707 | NECTIN3 | nectin cell adhesion molecule 3 [Source:HGNC Symbol;Acc:HGNC:17664] | protein_coding | -0.296 | -0.477 | -0.181 | 2.999 | 4.526 | 0.015 | turquoise |
| ENSG00000178104 | PDE4DIP | phosphodiesterase 4D interacting protein [Source:HGNC Symbol;Acc:HGNC:15580] | protein_coding | 0.276 | 0.260 | -0.016 | 8.902 | 7.032 | 0.002 | turquoise |
| ENSG00000179583 | CIITA | class II major histocompatibility complex transactivator [Source:HGNC Symbol;Acc:HGNC:7067] | protein_coding | 0.504 | 0.298 | -0.205 | 2.714 | 3.908 | 0.026 | turquoise |
| ENSG00000179776 | CDH5 | cadherin 5 [Source:HGNC Symbol;Acc:HGNC:1764] | protein_coding | 0.308 | 0.277 | -0.031 | 4.854 | 3.335 | 0.043 | turquoise |
| ENSG00000181126 | HLA-V | major histocompatibility complex, class I, V (pseudogene) [Source:HGNC Symbol;Acc:HGNC:23482] | transcribed_unprocessed_pseudogene | -1.238 | -0.060 | 1.177 | -2.874 | 3.899 | 0.026 | turquoise |
| ENSG00000185222 | TCEAL9 | transcription elongation factor A like 9 [Source:HGNC Symbol;Acc:HGNC:30084] | protein_coding | -0.222 | -0.171 | 0.051 | 3.550 | 4.252 | 0.019 | turquoise |
| ENSG00000187720 | THSD4 | thrombospondin type 1 domain containing 4 [Source:HGNC Symbol;Acc:HGNC:25835] | protein_coding | 0.313 | 0.215 | -0.097 | 5.703 | 4.725 | 0.013 | turquoise |
| ENSG00000188322 | SBK1 | SH3 domain binding kinase 1 [Source:HGNC Symbol;Acc:HGNC:17699] | protein_coding | 1.058 | 0.764 | -0.294 | -2.553 | 3.464 | 0.038 | turquoise |
| ENSG00000188846 | RPL14 | ribosomal protein L14 [Source:HGNC Symbol;Acc:HGNC:10305] | protein_coding | -0.162 | -0.160 | 0.002 | 6.353 | 3.993 | 0.024 | turquoise |
| ENSG00000189266 | PNRC2 | proline rich nuclear receptor coactivator 2 [Source:HGNC Symbol;Acc:HGNC:23158] | protein_coding | -0.147 | -0.016 | 0.131 | 5.390 | 4.693 | 0.013 | turquoise |
| ENSG00000197140 | ADAM32 | ADAM metallopeptidase domain 32 [Source:HGNC Symbol;Acc:HGNC:15479] | protein_coding | -0.573 | -0.384 | 0.189 | 1.613 | 3.865 | 0.027 | turquoise |
| ENSG00000215458 | AATBC | apoptosis associated transcript in bladder cancer [Source:HGNC Symbol;Acc:HGNC:51526] | lncRNA | 0.773 | 1.149 | 0.376 | -2.811 | 3.221 | 0.048 | turquoise |
| ENSG00000215912 | TTC34 | tetratricopeptide repeat domain 34 [Source:HGNC Symbol;Acc:HGNC:34297] | protein_coding | 0.856 | 0.295 | -0.561 | -1.270 | 4.869 | 0.011 | turquoise |
| ENSG00000225684 | FAM225B | family with sequence similarity 225 member B [Source:HGNC Symbol;Acc:HGNC:21865] | lncRNA | 0.183 | 1.725 | 1.542 | -1.866 | 4.692 | 0.013 | turquoise |
| ENSG00000225791 | TRAM2-AS1 | TRAM2 antisense RNA 1 (head to head) [Source:HGNC Symbol;Acc:HGNC:48663] | lncRNA | -0.341 | -0.239 | 0.103 | 1.938 | 4.942 | 0.011 | turquoise |
| ENSG00000229657 | AL391822.1 | ribosomal protein L13a (RPL13A) pseudogene | processed_pseudogene | 0.992 | 1.247 | 0.255 | -2.058 | 5.390 | 0.007 | turquoise |
| ENSG00000230373 | GOLGA6L5P | golgin A6 family like 5, pseudogene [Source:HGNC Symbol;Acc:HGNC:30472] | transcribed_unprocessed_pseudogene | 0.104 | -0.802 | -0.906 | -0.071 | 3.945 | 0.025 | turquoise |
| ENSG00000241790 | ENO1P4 | enolase 1 pseudogene 4 [Source:HGNC Symbol;Acc:HGNC:37945] | processed_pseudogene | 0.603 | 1.971 | 1.369 | -4.090 | 4.862 | 0.011 | turquoise |
| ENSG00000256977 | LIMS3 | LIM zinc finger domain containing 3 [Source:HGNC Symbol;Acc:HGNC:30047] | protein_coding | 0.710 | 0.401 | -0.309 | 0.728 | 3.222 | 0.048 | turquoise |
| ENSG00000257379 | AC023509.1 | novel transcript | lncRNA | 1.922 | 1.952 | 0.030 | -1.213 | 3.360 | 0.042 | turquoise |
| ENSG00000258484 | SPESP1 | sperm equatorial segment protein 1 [Source:HGNC Symbol;Acc:HGNC:15570] | protein_coding | -1.085 | -0.084 | 1.001 | 0.451 | 6.878 | 0.002 | turquoise |

| | | | | | | | | | | |
|---|---|---|---|---|---|---|---|---|---|---|
| ENSG00000265681 | RPL17 | ribosomal protein L17 [Source:HGNC Symbol;Acc:HGNC:10307] | protein_coding | -0.164 | -0.110 | 0.054 | 6.684 | 3.588 | 0.034 | turquoise |
| ENSG00000266086 | AC015813.2 | novel transcript | protein_coding | 0.796 | -0.206 | -1.003 | 0.548 | 5.413 | 0.007 | turquoise |
| ENSG00000268555 | AC123912.4 | novel transcript | lncRNA | -0.088 | 0.509 | 0.597 | 0.314 | 3.445 | 0.039 | turquoise |
| ENSG00000274272 | AC069281.2 | novel transcript | lncRNA | 1.326 | 1.305 | -0.021 | -1.874 | 3.944 | 0.025 | turquoise |
| ENSG00000280351 | AC127496.7 | TEC | TEC | 0.843 | 0.990 | 0.147 | -2.446 | 3.776 | 0.029 | turquoise |
| ENSG00000284691 | AC073111.4 | novel zinc finger protein | protein_coding | 0.334 | -0.138 | -0.471 | 0.670 | 5.207 | 0.009 | turquoise |
| ENSG00000287839 | AL353807.5 | novel transcript | lncRNA | 0.661 | 0.185 | -0.477 | -1.399 | 3.480 | 0.038 | turquoise |
| ENSG00000057294 | PKP2 | plakophilin 2 [Source:HGNC Symbol;Acc:HGNC:9024] | protein_coding | 0.256 | 0.132 | -0.124 | 7.028 | 3.897 | 0.026 | yellow |
| ENSG00000073910 | FRY | FRY microtubule binding protein [Source:HGNC Symbol;Acc:HGNC:20367] | protein_coding | 0.190 | 0.312 | 0.122 | 6.255 | 5.019 | 0.010 | yellow |
| ENSG00000101608 | MYL12A | myosin light chain 12A [Source:HGNC Symbol;Acc:HGNC:16701] | protein_coding | 0.454 | 0.080 | -0.374 | 7.847 | 5.757 | 0.005 | yellow |
| ENSG00000101871 | MID1 | midline 1 [Source:HGNC Symbol;Acc:HGNC:7095] | protein_coding | -0.291 | -0.147 | 0.144 | 4.153 | 3.352 | 0.042 | yellow |
| ENSG00000103710 | RASL12 | RAS like family 12 [Source:HGNC Symbol;Acc:HGNC:30289] | protein_coding | 0.258 | 0.124 | -0.134 | 4.180 | 3.637 | 0.033 | yellow |
| ENSG00000107165 | TYRP1 | tyrosinase related protein 1 [Source:HGNC Symbol;Acc:HGNC:12450] | protein_coding | 0.361 | 0.041 | -0.321 | 3.461 | 3.754 | 0.030 | yellow |
| ENSG00000116977 | LGALS8 | galectin 8 [Source:HGNC Symbol;Acc:HGNC:6569] | protein_coding | 0.232 | 0.058 | -0.174 | 5.499 | 4.494 | 0.016 | yellow |
| ENSG00000123901 | GPR83 | G protein-coupled receptor 83 [Source:HGNC Symbol;Acc:HGNC:4523] | protein_coding | 0.585 | -0.050 | -0.635 | -0.119 | 3.548 | 0.036 | yellow |
| ENSG00000133169 | BEX1 | brain expressed X-linked 1 [Source:HGNC Symbol;Acc:HGNC:1036] | protein_coding | -0.842 | -0.386 | 0.456 | 1.433 | 3.878 | 0.027 | yellow |
| ENSG00000134571 | MYBPC3 | myosin binding protein C3 [Source:HGNC Symbol;Acc:HGNC:7551] | protein_coding | 0.233 | 0.156 | -0.077 | 8.845 | 3.908 | 0.026 | yellow |
| ENSG00000136040 | PLXNC1 | plexin C1 [Source:HGNC Symbol;Acc:HGNC:9106] | protein_coding | -0.339 | 0.023 | 0.362 | 2.522 | 3.345 | 0.043 | yellow |
| ENSG00000136144 | RCBTB1 | RCC1 and BTB domain containing protein 1 [Source:HGNC Symbol;Acc:HGNC:18243] | protein_coding | 0.222 | 0.135 | -0.087 | 3.780 | 4.322 | 0.018 | yellow |
| ENSG00000136932 | TRMO | tRNA methyltransferase O [Source:HGNC Symbol;Acc:HGNC:30967] | protein_coding | -0.243 | -0.202 | 0.041 | 1.848 | 3.464 | 0.038 | yellow |
| ENSG00000141664 | ZCCHC2 | zinc finger CCHC-type containing 2 [Source:HGNC Symbol;Acc:HGNC:22916] | protein_coding | 0.170 | -0.115 | -0.284 | 3.247 | 3.764 | 0.030 | yellow |
| ENSG00000146197 | SCUBE3 | signal peptide, CUB domain and EGF like domain containing 3 [Source:HGNC Symbol;Acc:HGNC:13655] | protein_coding | 0.685 | 0.080 | -0.605 | 0.040 | 3.592 | 0.034 | yellow |

| | | | | | | | | | | |
|---|---|---|---|---|---|---|---|---|---|---|
| ENSG00000148660 | CAMK2G | calcium/calmodulin dependent protein kinase II gamma [Source:HGNC Symbol;Acc:HGNC:1463] | protein_coding | 0.295 | 0.242 | -0.053 | 2.768 | 4.071 | 0.023 | yellow |
| ENSG00000148925 | BTBD10 | BTB domain containing 10 [Source:HGNC Symbol;Acc:HGNC:21445] | protein_coding | -0.219 | 0.021 | 0.240 | 3.754 | 5.681 | 0.006 | yellow |
| ENSG00000163145 | C1QTNF7 | C1q and TNF related 7 [Source:HGNC Symbol;Acc:HGNC:14342] | protein_coding | -0.494 | -0.335 | 0.159 | 2.377 | 4.599 | 0.014 | yellow |
| ENSG00000163681 | SLMAP | sarcolemma associated protein [Source:HGNC Symbol;Acc:HGNC:16643] | protein_coding | 0.367 | 0.186 | -0.181 | 7.039 | 3.787 | 0.029 | yellow |
| ENSG00000166974 | MAPRE2 | microtubule associated protein RP/EB family member 2 [Source:HGNC Symbol;Acc:HGNC:6891] | protein_coding | 0.200 | 0.076 | -0.124 | 6.062 | 3.448 | 0.039 | yellow |
| ENSG00000175084 | DES | desmin [Source:HGNC Symbol;Acc:HGNC:2770] | protein_coding | 0.238 | 0.191 | -0.047 | 10.761 | 7.549 | 0.001 | yellow |
| ENSG00000175182 | FAM131A | family with sequence similarity 131 member A [Source:HGNC Symbol;Acc:HGNC:28308] | protein_coding | 0.271 | 0.294 | 0.022 | 2.613 | 3.290 | 0.045 | yellow |
| ENSG00000196109 | ZNF676 | zinc finger protein 676 [Source:HGNC Symbol;Acc:HGNC:20429] | protein_coding | -0.418 | -0.103 | 0.315 | 1.288 | 3.306 | 0.044 | yellow |
| ENSG00000197977 | ELOVL2 | ELOVL fatty acid elongase 2 [Source:HGNC Symbol;Acc:HGNC:14416] | protein_coding | 0.501 | 0.026 | -0.475 | 0.202 | 3.724 | 0.031 | yellow |
| ENSG00000198467 | TPM2 | tropomyosin 2 [Source:HGNC Symbol;Acc:HGNC:12011] | protein_coding | 0.213 | 0.130 | -0.082 | 7.110 | 3.314 | 0.044 | yellow |
| ENSG00000198952 | SMG5 | SMG5 nonsense mediated mRNA decay factor [Source:HGNC Symbol;Acc:HGNC:24644] | protein_coding | 0.207 | 0.103 | -0.104 | 5.130 | 4.797 | 0.012 | yellow |
| ENSG00000205084 | TMEM231 | transmembrane protein 231 [Source:HGNC Symbol;Acc:HGNC:37234] | protein_coding | -0.394 | -0.108 | 0.286 | 2.287 | 4.462 | 0.016 | yellow |
| ENSG00000244306 | AL589743.1 | double homeobox A pseudogene 10 | transcribed_processed_pseudogene | -1.053 | -0.920 | 0.133 | 0.051 | 4.192 | 0.020 | yellow |
| ENSG00000260596 | DUX4 | double homeobox 4 [Source:HGNC Symbol;Acc:HGNC:50800] | protein_coding | -1.036 | 0.593 | 1.629 | -2.604 | 3.444 | 0.039 | yellow |
| ENSG00000261485 | PAN3-AS1 | PAN3 antisense RNA 1 [Source:HGNC Symbol;Acc:HGNC:39932] | lncRNA | 0.771 | 0.511 | -0.260 | -1.666 | 5.096 | 0.009 | yellow |
| ENSG00000285238 | AC006064.6 | novel transcript | protein_coding | 1.987 | 2.646 | 0.659 | -2.431 | 4.253 | 0.019 | yellow |

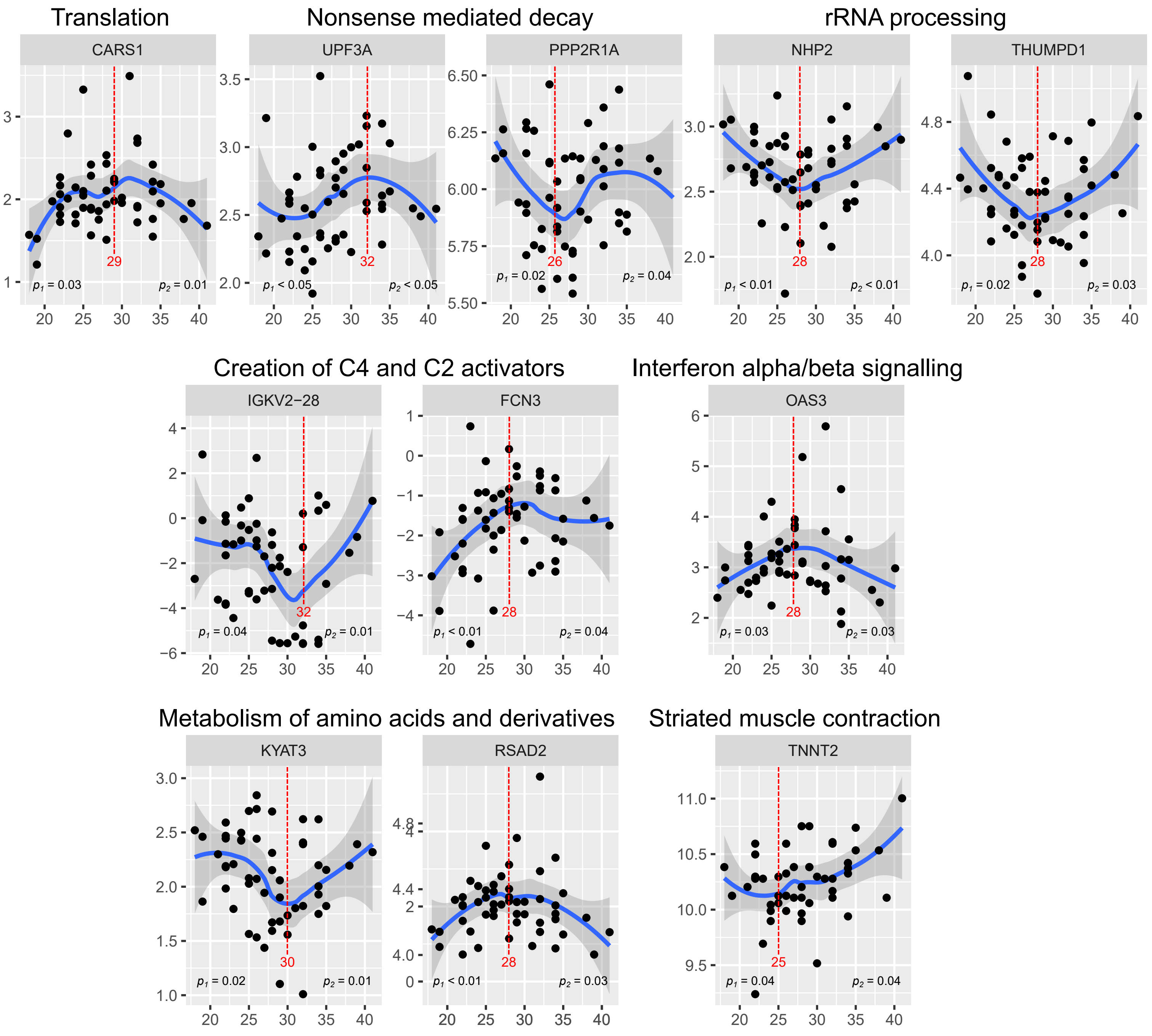

**Figure S1** – Plots of transcripts with the membership in significant pathways and showing biphasic BMI relationship. The blue line shows loess regression trend. Red line indicates a breakpoint identified by the Two Line method; p1 and p2 -values indicate significant regression before or after the breakpoint.

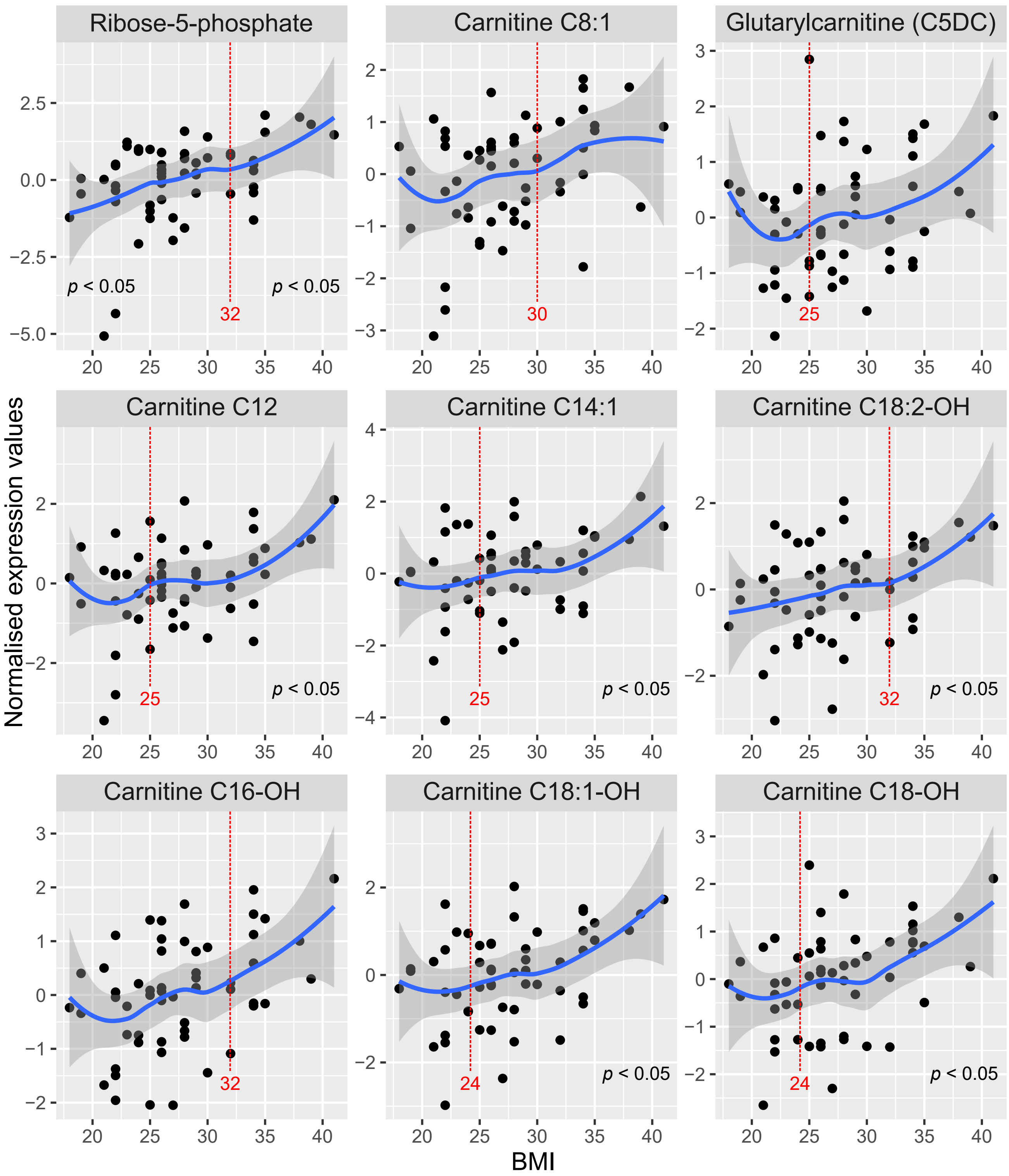

**Figure S1** – Plots of transcripts with the membership in significant pathways and showing biphasic BMI relationship. The blue line shows loess regression trend. Red line indicates a breakpoint identified by the Two Line method; p1 and p2 -values indicate significant regression before or after the breakpoint.
